# Drivers of Mortality in COVID ARDS Depend on Patient Sub-Type

**DOI:** 10.1101/2022.07.04.22277239

**Authors:** Helen Cheyne, Amir Gandomi, Shahrzad Hosseini Vajargah, Victoria M. Catterson, Travis Mackoy, Lauren Mccullagh, Gabriel Musso, Negin Hajizadeh

## Abstract

**Background:** The most common cause of death in people with COVID-19 is acute respiratory distress syndrome (ARDS). ARDS is a heterogeneous syndrome, however, subgroups that have been identified among non-COVID-19 ARDS patients do not clearly apply to COVID-19 ARDS patients. Additionally, studies of COVID-19 ARDS have been limited by sample size.

**Methods:** We applied an iterative clustering and machine learning framework to electronic health record data from thousands of hospitalized COVID-19 ARDS patients with the goal of defining and characterizing clinically-relevant COVID-19 ARDS subgroups (phenoclusters). We then applied a supervised model to identify risk factors for hospital mortality for each phenocluster and compared these between phenoclusters and the entire cohort.

**Findings:** Risk factors that predict mortality in the overall cohort of COVID-19 ARDS patients do not necessarily predict mortality in phenoclusters. In fact, some risk factors increase the risk of hospital mortality in some phenoclusters, but decrease mortality in others.

**Interpretation:** These phenocluster-specific risk factors would not have been observed with a single predictive model. Heterogeneity in phenoclusters of COVID-19 ARDS as well as drivers of mortality may partially explain challenges in finding effective treatments when applied to all patients with ARDS.

**Funding:** This work was supported by philanthropic funds to the Feinstein Institutes for Medical Research. The funding source did not control any aspect of the study and did not review the results. All authors had full access to the full data in the study and accept responsibility to submit for publication.

## Introduction

The acute respiratory distress syndrome (ARDS) is a leading cause of intensive care unit (ICU) admissions and mortality, and the leading cause of death in people infected with SARS-CoV2, carrying a 50-80% mortality rate^1^. Recognizing the heterogeneity in this disease, investigators have attempted to sub-group ARDS patients, with hopes that this can lead to tailored treatments and decreased mortality^2^. Whether these ARDS sub-groups are similar to those found in COVID-19 associated ARDS is unclear.

Machine learning (ML) based frameworks present substantial potential for the improvement of clinical care^3^. Specifically, clustering is a powerful unsupervised ML tool for discovering patterns and structure in datasets, while supervised predictive frameworks can be leveraged to identify features associated with outcomes that are not apparent through classic statistical approaches. When applied in combination with traditional approaches, ML models have shown effectiveness in predicting patient outcomes across disease states including Sepsis, ICU admission, and Asthma/COPD^4–6^. Unlike supervised ML methods, that in this context would use known associations between patient data and outcomes to train predictive models, clustering is unsupervised, and instead looks to categorize patients into phenoclusters where patients are more like each other than patients in other phenoclusters.

Only two studies have looked specifically at sub-phenotypes of COVID-19 ARDS. Ranjeva *et al* studied ICU patients with COVID-19 ARDS and found two distinct phenoclusters^7^. The phenocluster that had double the mortality (40% vs. 23% odds of 28 day-mortality) had higher coagulation factors and more end-organ dysfunction, but only mildly increased relative hyperinflammation. In contrast, Sinha *et al*. (2020) identified two phenoclusters that correlated well with the previously identified hyper-and hypo-inflammatory groups among ARDS patients^8^. Both studies included fewer than 250 patients.

We sought to further examine COVID-19 ARDS phenoclusters and risk factors for in-hospital mortality using a large dataset of COVID-19 ARDS patients. In contrast to other studies, onset of ARDS was labeled at the time of identification of severe hypoxemia during hospitalization, which may have been before intubation. This approach would allow for earlier determination of phenoclusters and was less susceptible to bias/error based on variability of decisions about timing of intubation.

## Methods

### Study Design, Participants and Study Size

This was a retrospective cohort study and was conducted in multiple phases (**Figure 1**). A detailed explanation of all methods is provided in the **Supplemental Methods**. Supervised (S), semi-supervised (SS), and unsupervised U) methods were used for feature selection (S), model selection (SS), participant cluster modeling (U), and mortality risk regression (S).

**Figure 1:**
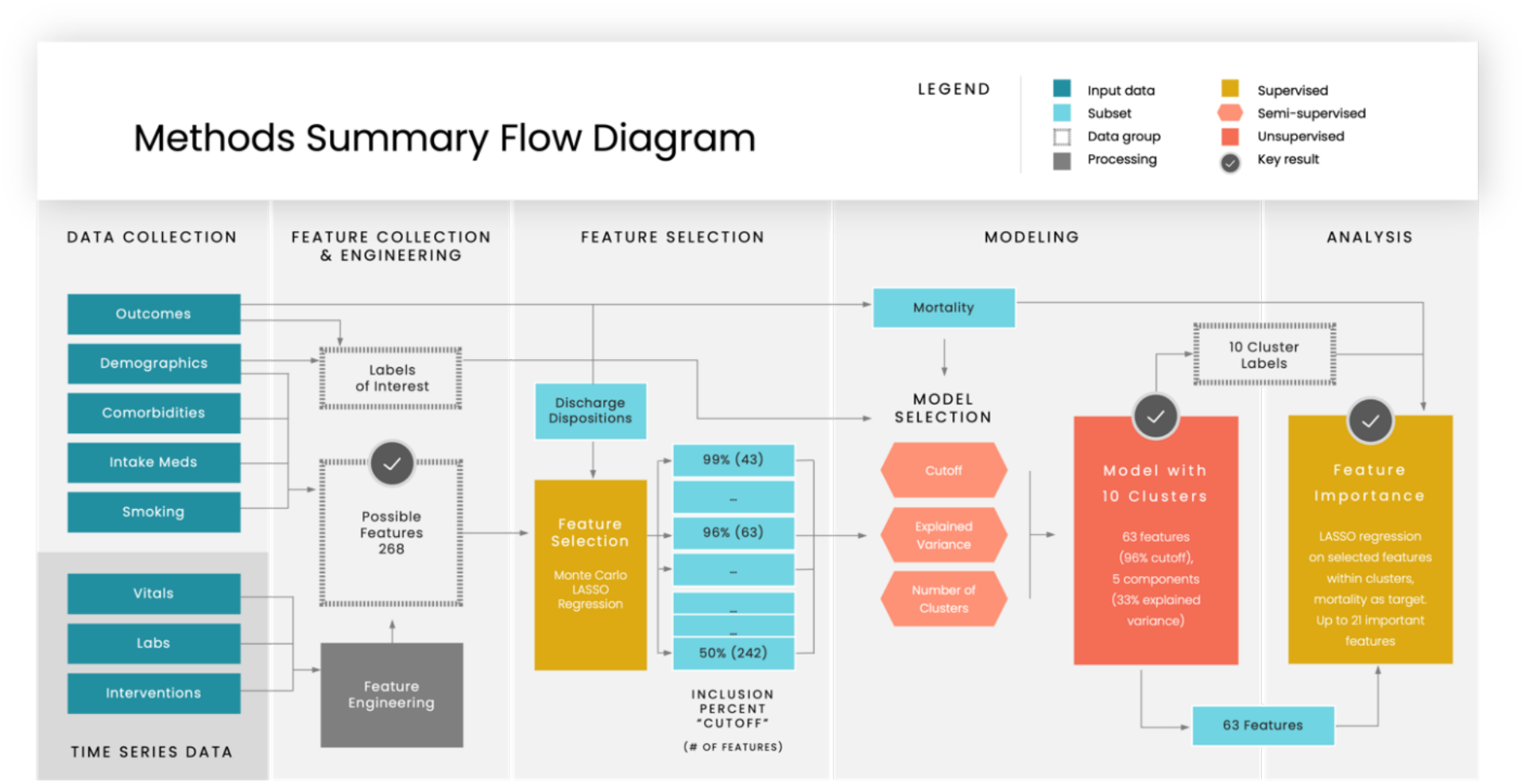
Methods summary flow diagram. Our process began by collecting both intake and time series data regarding patients who developed COVID-19 ARDS. The time series data were aggregated in to up to 5 descriptive metrics through feature engineering. Only variables that were known at or before 24 hours after the onset of hypoxemia were considered as possible features. Variables that were of interest were grouped together and referred to as labels. The possible features underwent feature selection based on association with discharge dispositions, and resulted in subsets of features with various inclusion percent “cut-off” levels. The lower the “cut-off” the more features were included. Using 5-fold cross-validation the ideal set of parameters (“cut-off”, explained variance included from principal component analysis, and the number of phenoclusters) for the model was chosen. This selection was based on enrichment of mortality, of other labels of interest, and of the selected features. The parameters chosen were a 96% feature selection inclusion criteria “cut-off” which included 63 features, 33% explained variance which reduced the selected features into 5 orthogonal components, and 10 phenoclusters. Finally, the feature importance relative to mortality was analysed in each cluster for all 63 selected features using LASSO regression.

This study used the Northwell COVID-19 ARDS dataset (NorthCARDS) including patients from 12 hospitals across the Northwell Health system in the New York City area^9^. Our study sample consisted of all patients who fit the Berlin criteria for ARDS^10^. Although unsupervised data models do not require a specific sample size, larger datasets allow for more granular differences to be identified. The dataset is segmented into two waves. Wave 1 included 1,901 patients admitted between February 16th to April 30th, 2020 and Wave 2 included 963 patients admitted between November 1st to February 28th, 2021. All hospitalizations have a final discharge disposition (i.e., their hospital outcomes are known). We looked for variations between the waves using descriptive analyses (see **Tables 1** and **2**).

**Table 11:**
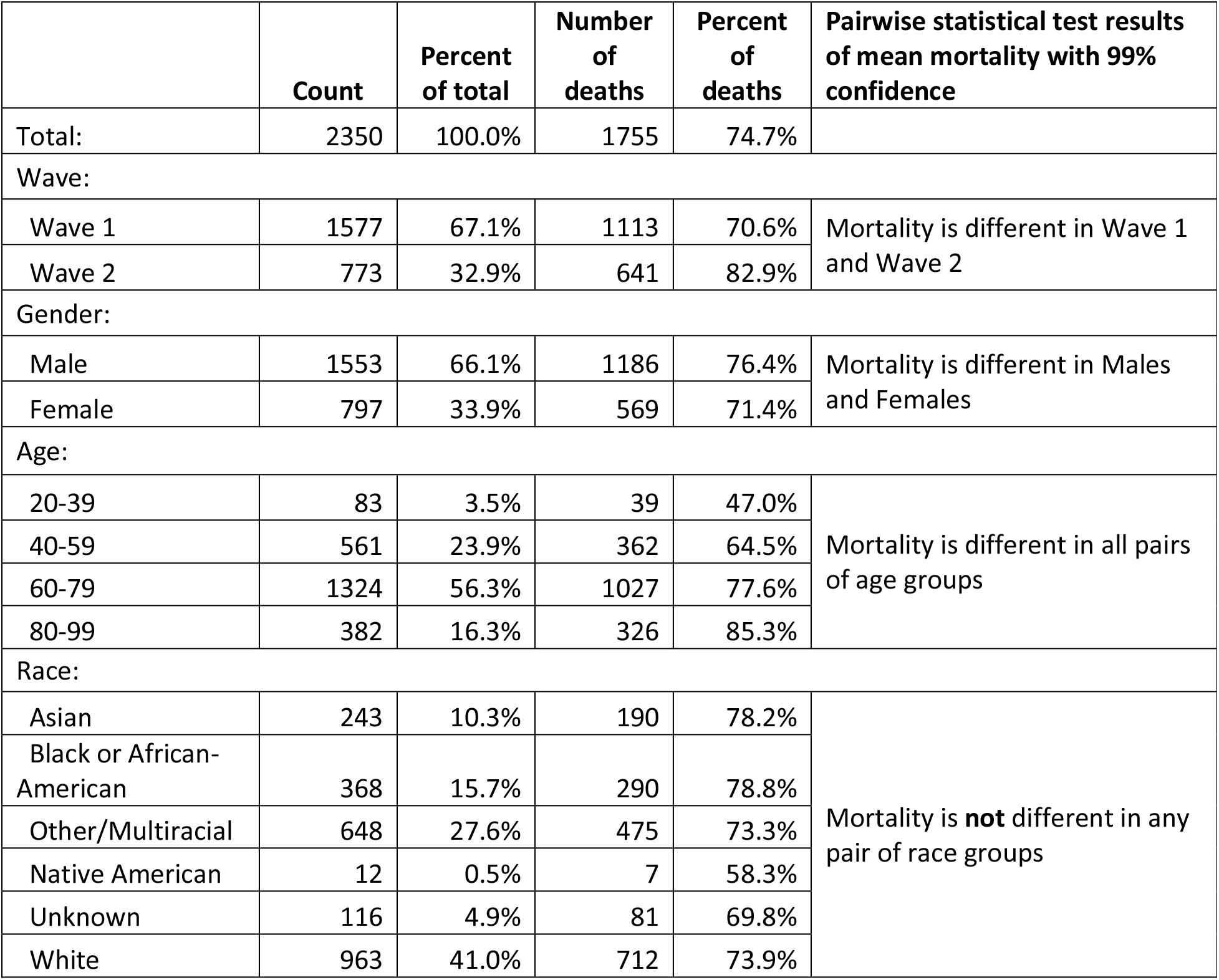
Mortality by wave, age, gender, and race in training cohort

**Table 2:** 2 Distributions of discharge dispositions in the full training set.

|  | Count (percent) |  |  |
| --- | --- | --- | --- |
|  | All | Wave 1 | Wave 2 |
| Discharge Disposition: |  |  |  |
| Expired* | 1749 (74.4%) | 1113 (70.6%) | 636 (82.3%) |
| Facility Extubated | 219 (9.3%) | 156 (9.9%) | 63 (8.2%) |
| Facility Intubated | 84 (3.6%) | 66 (4.2%) | 18 (2.3%) |
| Home | 298 (12.7%) | 242 (15.3%) | 56 (7.2%) |
| *Expired number slightly higher than in-hospital mortality due to 6 people who had reports of death within 1 month of discharge in the EMR |  |  |  |

### Variables: outcomes, predictors, confounders, and modifiers

Outcome variables included discharge disposition and mortality. Categorical descriptive data on each patient included comorbidities, intake medications, smoking status, demographics, and responsiveness to steroids. Longitudinal descriptive data on each patient included labs and vitals, and was aggregated into median, range, slope, inter-quartile range (IQR), and fluctuations from hospital admission to 24 hours after ARDS onset (T0+24), as applicable. For details on aggregations and engineered features see **Supplemental Methods**.

### Addressing Bias in the Patient Dataset

We mitigated selection bias by including all patients who were identified as having ARDS in the NorthCARDS database, which includes patients from the socioeconomically and racially diverse demographic of the Northwell Health system. However, there are known racial disparities in hospitalizations and deaths across US health systems which our study could not have overcome^11^. Our inclusion of patients at the time of onset of severe hypoxemia in the hospital regardless of intubation status allowed for identification of people who had ARDS independent of patient/clinician decisions to intubate. We attempted to mitigate information bias through manual investigation of outliers or unusual findings. For example, the large number of ‘unknown’ values for smoking status likely represented people who were incapacitated on admission and who could not complete this field in the setting of a large volume of patient admissions. Similarly, we binned data into 8-hour time frames reflecting the real-world delays in nursing measurement recording during the COVID-19 pandemic. Exposures and outcomes were both known on an individual level thus minimizing ecological fallacy.

### Data Cleaning

Categorical variables were encoded, and continuous variables were standardized to have a mean of 0 and a standard deviation of 1 to ensure consistent contribution to models.

### Segmentation of the Dataset into Train and Test Sets

Data were divided into six equal folds, stratified based on discharge disposition and demographics. Five of the folds were leveraged for training and cross validation, and the sixth was withheld as a final test data set. All 33 censored patients (those that were still hospitalized when the data was initially collected) were placed in the final test data set.

### Separation of Model Targets from Possible Features

Model targets or outcomes included discharge disposition and mortality. Variables considered as features for the model included demographics, comorbidities, home medications, responsiveness to steroids, labs, and vitals up to 24 hours after the onset of hypoxemia (see **Table S4**).

### Feature Selection Through Supervised Machine Learning

Monte Carlo logistic regression models with L1 regularization (LASSO) were used for feature selection. They were trained using each discharge disposition as the target and a subset of the possible features as the predictors. The Monte Carlo strategy bookends these models, on one side by randomly selecting a subset of the possible 268 predictors 1000 times, and on the other end by aggregating the results into a ranked list of selection-worthy features. How many of these features were used, was determined in the next step in conjunction with other model parameters, based on the aggregated proportion of models in which that feature was a contributing factor (cut-off).

### Clustering Framework and Selected Model Parameters

K-means clustering was used to group patients based on principal components calculated from a cut-off defined feature set with a fixed explained variance level. Models were fit for a variety of feature selection thresholds, PCA explained variance levels, and number of K-means clusters and then examined. The selected model parameters were chosen simultaneously because they are interdependent. A three-step process was implemented to select the final model. It primarily prioritized models that have clusters with distinct mortality characteristics as well as clusters that are distinguished by a variety of other characteristics of interest features (meaning the model which identified the most differences in mean values across all clusters among the most variables). This process is detailed in the **Supplemental Methods**.

### Analysis of Factors Driving Mortality

Once the phenoclusters were identified an L1 regularized (LASSO) regression was performed to understand the driving factors behind mortality rates for each phenocluster and the whole cohort. These regression models were implemented using mortality as the target, and the same 63 selected features used for phenoclustering. Weights of features were compared between phenoclusters and to that of the whole cohort to understand the similarities and differences in risk factors.

## Results

### Participants and Descriptive Data

There were 2,350 people in the training cohort, and 526 held out for testing. 67% were from Wave 1, 66% were males, 72% were age 60 or over, and 41% were identified as white (**Table 1**). The largest number of patients was in the age group 60-79. Wave 2 was older in age, had more males and higher mortality rates. Surprisingly, race categories did not have significant differences in mortality among ARDS patients. Overall, approximately half of patients who were discharged were sent home extubated (298 of 601). The other half of discharged patients were sent to a facility, including 27% (84 of 303) who were still intubated (**Table 2**). The average length of stay was 22.8 days across both waves.

### Feature Differences Between Higher Mortality and Lower Mortality Groups to Full Dataset

Using 5-fold cross validation within our clustering framework, we selected a feature selection cut-off parameter of 96%, 63 features were selected from 268 variables available. The selected features include 3 demographic features, six of each of comorbidities and home medications, 11 vitals features and 30 lab features. (**Table S4**). Principal component analysis reduced these to five principal components with explained variance of 33%, which were used for clustering into 10 phenoclusters. Using the selected parameters (63 features, 33% explained variance, and 10 phenoclusters) a model was trained on the full training cohort. Each of phenoclusters 1 and 4 (C1 and C4) have significantly lower mortality when compared to the rest of the cohort (p = 1.59e-14 C1; p = 7.90e-6 C4) and we describe these as the lower mortality group. Each of phenoclusters 2 and 5 (C2 and C5) have significantly higher mortality than the rest of the cohort (p = 5.65e-6 C2; p = 3.93e-9 C5), and we describe these as the higher mortality group (**Table 3**).

**Table 33:** Patient statistics by cluster (training set). The two clusters in the higher mortality group and lower mortality group are highlighted in pink and blue respectively (enriched mortality, meaning that the mean proportion of deaths was statistically significantly different than the mean in the rest of the training cohort data at the 95% confidence level).

| Cluster | Number in cluster<br>(% in cluster) | Hospital Mortality | Number in cluster, Wave 1<br>(% of Wave 1 in cluster) | Hospital Mortality, Wave 1 | Number in cluster, Wave 2<br>(% of Wave 2 in cluster) | Hospital Mortality, Wave 2 |
| --- | --- | --- | --- | --- | --- | --- |
| **C 1** | 563 (24%) | 62.0% | 413 (26.2%) | 55.4% | 150 (19.4%) | 80.0% |
| **C 2** | 481 (20.5%) | 82.5% | 258 (16.4%) | 77.9% | 223 (28.8%) | 87.9% |
| **C 3** | 325 (13.8%) | 77.2% | 241 (15.3%) | 75.1% | 84 (10.9%) | 83.3% |
| **C 4** | 262 (11.1%) | 63.0% | 203 (12.9%) | 64.0% | 59 (7.6%) | 59.3% |
| **C 5** | 232 (9.9%) | 89.7% | 151 (9.6%) | 87.4% | 81 (10.5%) | 93.8% |
| **C 6** | 212 (9.0%) | 81.6% | 160 (10.1%) | 79.4% | 52 (6.7%) | 88.5% |
| **C 7** | 113 (4.8%) | 77.0% | 40 (2.5%) | 72.5% | 73 (9.4%) | 79.5% |
| **C 8** | 84 (3.6%) | 82.1% | 72 (4.6%) | 81.9% | 12 (1.6%) | 83.3% |
| **C 9** | 70 (3.0%) | 70.0% | 35 (2.2%) | 65.7% | 35 (4.5%) | 74.3% |
| **C10** | 8 (0.3%) | 87.5% | 4 (0.3%) | 75.0% | 4 (0.5%) | 100.0% |
| **Totals** | 2350 (100%) | 74.7% | 1577 (100%) | 70.6% | 773 (100%) | 82.9% |

There are notable differences in average values for several variables when comparing the lower versus higher mortality groups (C1 or C4 vs. C2 or C5), and between phenoclusters within mortality group (C1 vs. C4 and C2 vs. C5) ((**Figure 2**; **Supplemental Methods**). Although these descriptive differences characterize each phenocluster, they do not imply causal link to increased or decreased mortality. The higher mortality group had more people who were older, white, and had more hospitalizations in Wave 2. In addition, there were higher rates of Atrial Fibrillation or Diabetes, home prescriptions for beta blockers, calcium channel blockers, and NSAIDs. The lower mortality group had fewer people with diagnosis of dementia or other neurological illness.

**Figure 2:**
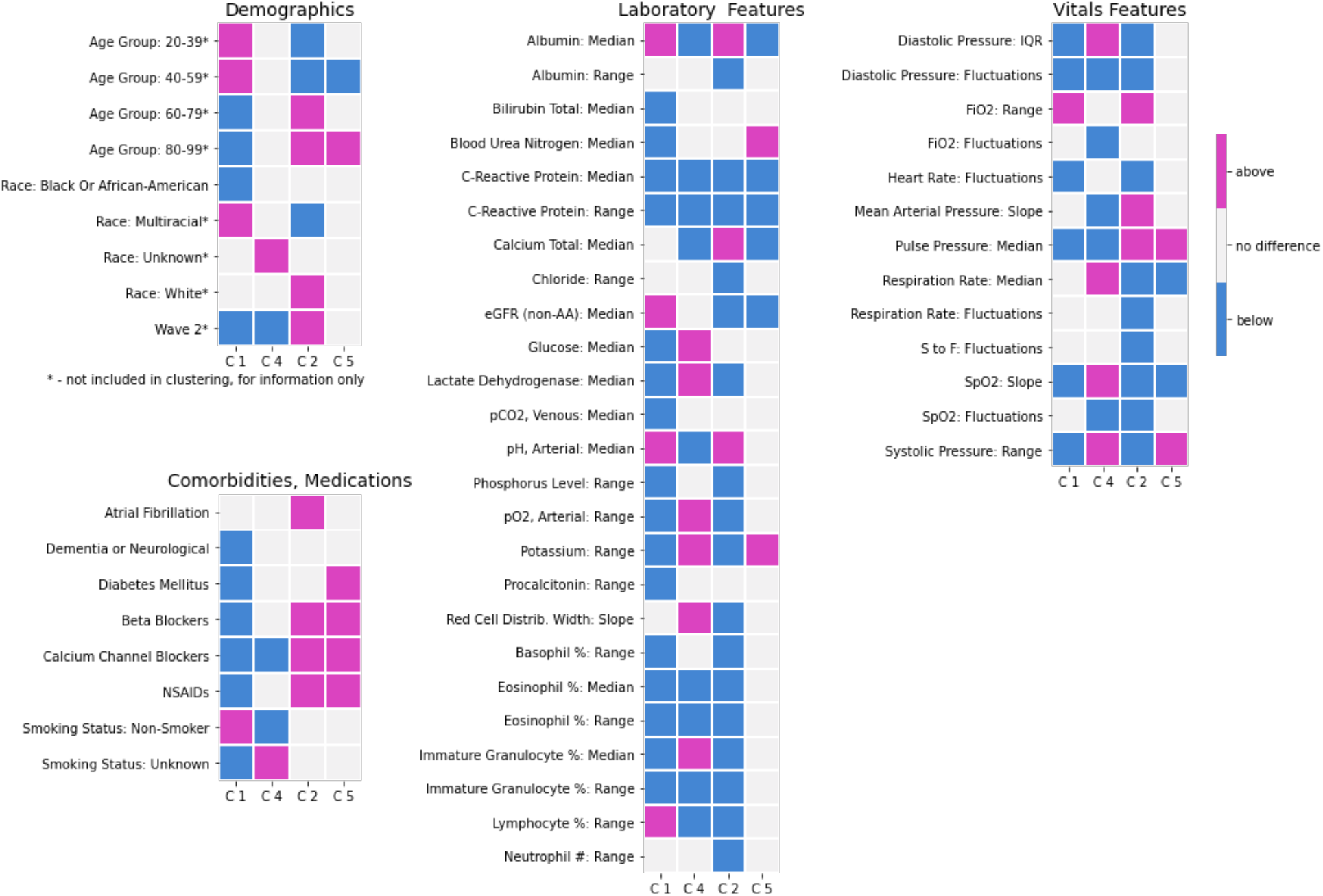
Heatmap of descriptive features separated by feature type. Displayed here are variables that are significantly enriched (mean values that were significantly different from the rest of the whole cohort, p value < 0.05) within indicated clusters. The two left columns in each figure represent the lower mortality group (C1 and C4) and the two columns on the right represent the higher mortality group (C2 and C5). Pink indicates a higher mean value for the feature compared to the rest of the cohort, blue indicates a lower mean value and grey indicates no significant difference from the rest of the cohort. Of note, for some features there are opposite patterns seen within the same mortality group. For example, for systolic pressure range C1 has a lower value but C4 has a higher value in the lower mortality group. Alternately, C2 has a lower value and C5 has a higher value in the higher mortality group.

Surprisingly, both higher and lower mortality groups had lower CRP levels than those in the other phenoclusters, with average values in the 20mg/L range compared to the whole cohort average of 41 units mg/L. Of note, values for all the phenoclusters are highly elevated compared to normal/healthy people (normal levels 8-10mg/L). Both C2 and C5 had higher median pulse pressure (a known risk factor for cardiovascular mortality especially in older people^12^), lower median respiratory rate, and lower SpO2 slope meaning less rapidly increasing SpO2 over the hospital course. RDW slope indicated more rapidly increasing RDW over the course of the hospital stay (higher slope) in the lower mortality group (C4); and less rapidly increasing, even slightly decreasing RDW over time in the hospital lower in the higher mortality group (C2).

### Risk Factors for Mortality for the Overall Cohort

When looking at the overall cohort, categorical features that predicted increased mortality were home medications including ACE inhibitors, NSAIDs, beta blockers or calcium channel blockers (**Figures 3** and **4**). Surprisingly, a diagnosis of dementia or other neurologic disease was found to be protective. In subsequent analyses exploring potential reasons for this surprising finding, we discovered that people with diagnosis of dementia were intubated earlier on average (**Figure 5**). Laboratory values predicting increased mortality were high levels of median CRP, LDH, bilirubin, calcium, and BUN. Increased levels of eGFR and arterial PH were protective. Rapidly decreasing glucose predicted increased mortality possibly capturing the patients who had hyperglycemia or diabetic ketoacidosis, which was seen in many patients with COVID-19 critical illness.^15^ Several vital sign trends were protective, including rapidly increasing SpO2, high arterial PO2 range, high median respiratory rate, and high range in systolic blood pressure and FiO2.

**Figure 3:**
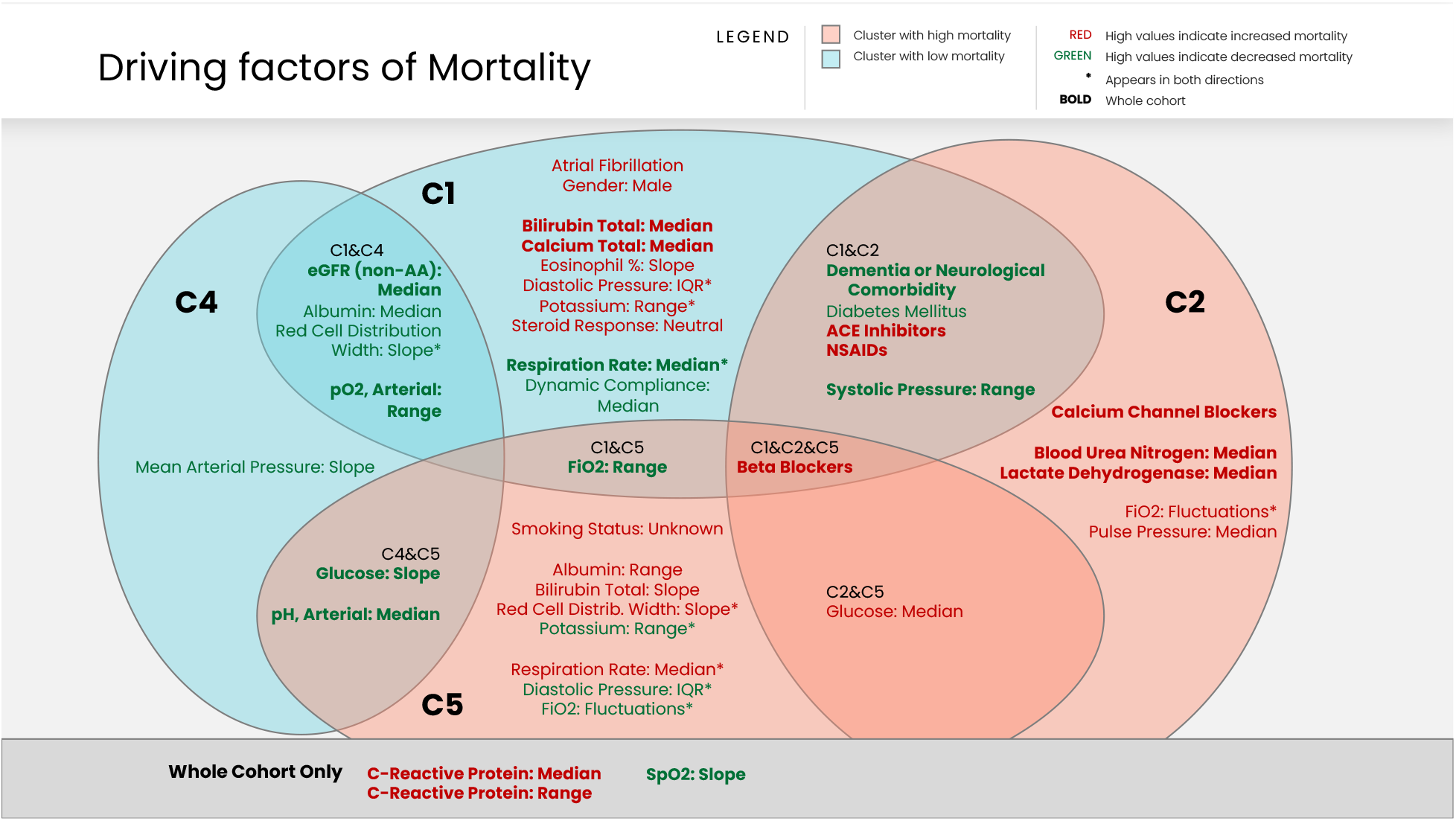
Venn diagram comparing the driving factors of mortality. This diagram shows features that are associated with mortality in one or more of clusters, as well as in the whole patient cohort. Background color indicates to which mortality group a cluster belongs. Text color indicates the direction of the association: Green text means that higher values are associated with decreased mortality and lower values are associated with increased mortality; Red text means the opposite, that higher values are associated with increased mortality and lower values are associated with decreased mortality.

**Figure 4:**
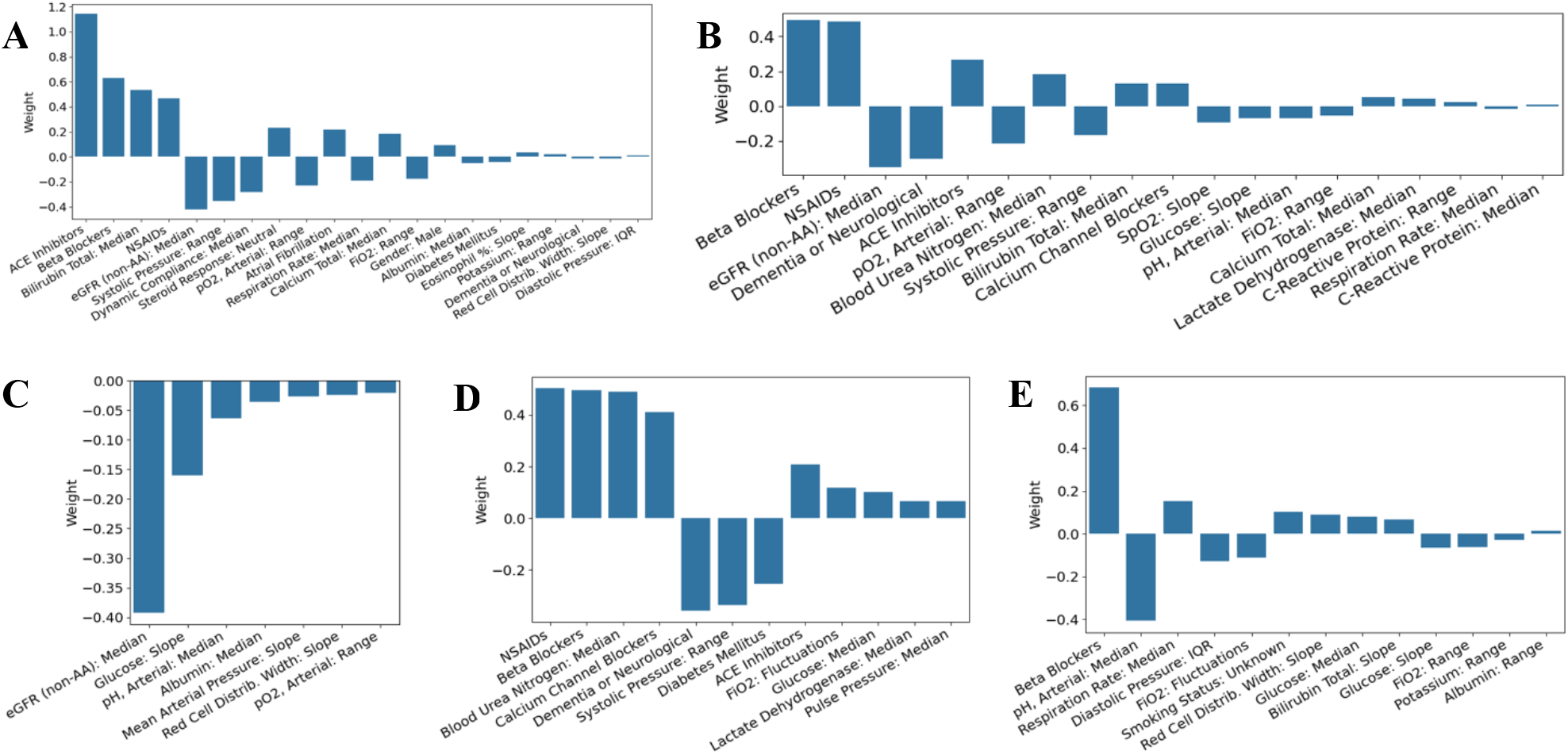
Feature importance coefficients for predicting mortality. Positive values indicate that patients above average in these values relative to the whole cohort have higher mortality risk. Negative values indicate that patients with values that are above average relative to the whole cohort have lower mortality risk. The ‘weight’ on the y axis are the coefficients of a logistic (linear) regression and represent the relative weight each standardized variable has on the outcome (hospital death). Positive weights mean that when that variable is higher, all else equal, the likelihood of dying is higher. The larger the weight, the more impact the variable has on the outcome. A one unit change for a variable with a weight of 0.4 will have double the impact on the outcome than a one unit change for a variable with a weight of 0.2. (A) whole cohort, (B) C1, (C) C4, (D) C2, (E) C5.

**Figure 5:**
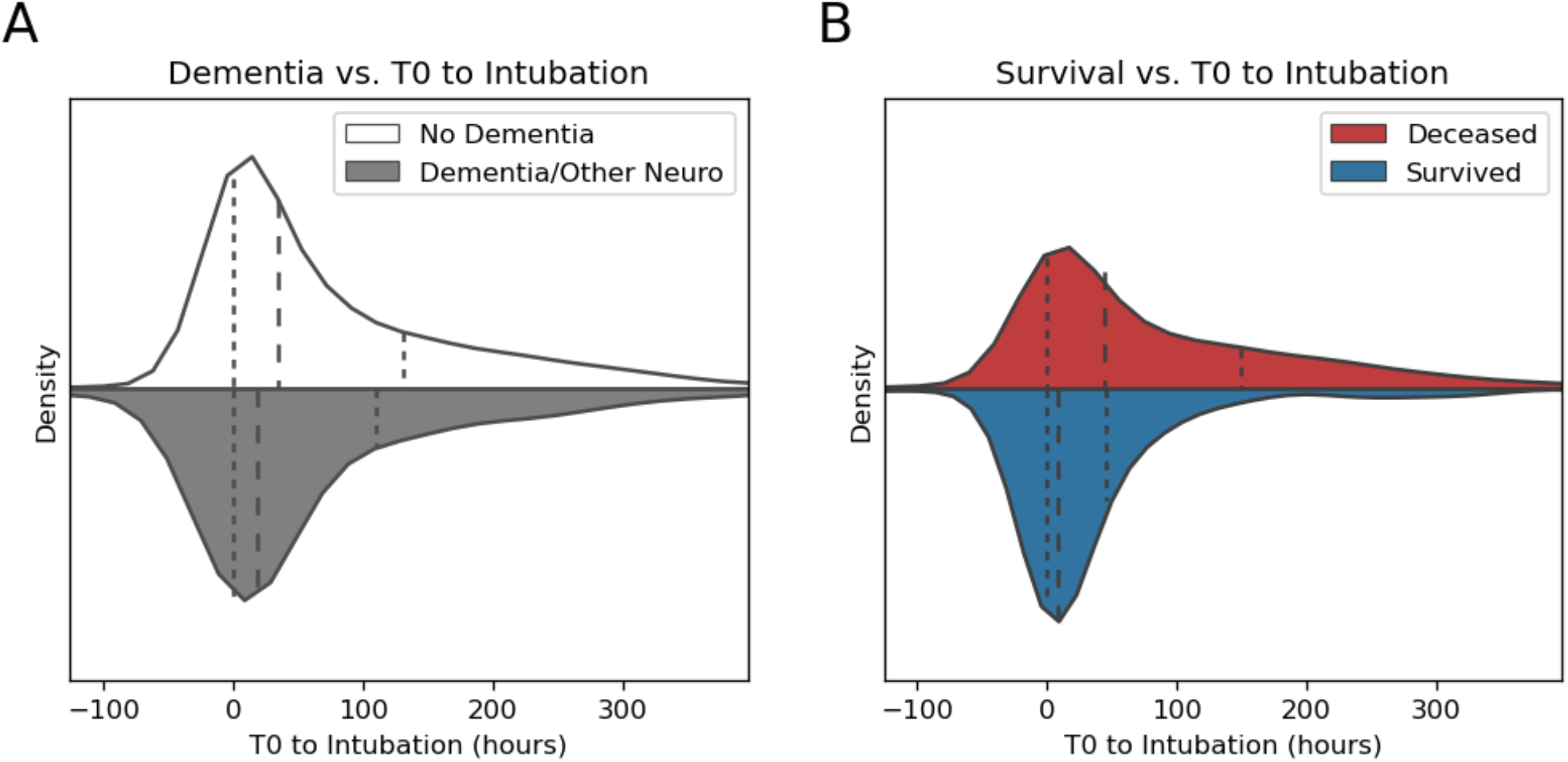
Violin plots demonstrating the outcome of the Mann-Whitney tests. In (A) the distributions of time from onset of hypoxemia to intubation in people with a dementia or other neurological diagnosis and people who do not such a diagnosis. It can be seen that the median (long dashed lines) and the IQR (short dashed lines) for those with dementia (grey) are lower than those without. In (B) the analogous comparison in made between people who survived and people who were deceased. Again, though more dramatically, this shows that the median and IQR for those who survived is lower than for those who did not.

### Comparison of Overall Findings with Phenocluster-specific risk factors

Figure 3. summarizes the overlapping and unique risk factors that increase or decrease mortality in the whole cohort, and in the two lower mortality versus higher mortality phenoclusters. There were several additional variables that predicted mortality when looking within phenoclusters. The lower mortality group’s risk factors were high values for eGFR, median albumin, arterial PO2, and rapidly increasing RDW, which were all protective. Interestingly, CRP was not a significant predictor of mortality when looking within phenoclusters.

Male gender was only a mortality risk factor for those in C1, in addition to atrial fibrillation comorbidity, high median bilirubin, calcium, rapidly increasing eosinophil %, higher diastolic blood pressure IQR, and potassium range. A lack of response to steroids, and a low pulmonary compliance were also both only predictive of increased mortality in C1. On the other hand, high median respiratory rate was protective. Dementia and diabetes comorbidities were protective in C1 as well as in one of the higher mortality phenoclusters C2, as was seen in the overall cohort analysis. Home medications that predicted increased mortality included ACE Inhibitors and NSAIDS for those in C1 and C2, and beta blockers for those in C1, C2, and C5. Calcium channel blockers were only a risk factor for those in C2. No baseline medications were identified as predicting increased mortality risk in C4.

In C5, ‘unknown’ smoking status predicted higher mortality likely reflecting people who were hospitalized without the opportunity to obtain a social history. High albumin range, rapidly increasing bilirubin and low potassium range predicted higher mortality. High median glucose predicted higher mortality in the high mortality group but was not a risk factor in the lower mortality group. However, rapidly decreasing glucose level predicted higher mortality in one of the phenoclusters from the low and high mortality groups (C4 and C5), which was also found when looking at the whole (un-clustered) cohort. High median pulse pressure was only a mortality predictor in C2.

Several features had opposite direction of risk depending on the phenocluster. Specifically, in contrast to C1, a high potassium range was protective in C5, as was a higher diastolic blood pressure IQR. On the other hand, in C5, a rapidly increasing RDW predicted increased mortality, as did a high median respiratory rate. Within the higher mortality group, in phenocluster C2, fluctuations in FiO2 predicted higher mortality but predicted lower mortality for those in C5.

## Discussion

Our analysis of 2876 patients with COVID-19 ARDS found 10 distinct phenoclusters, with varied risk factors for hospital mortality. These cluster-specific risk factors likely reflect different pathophysiologic processes that lead to death, suggesting avenues for more tailored targets for treatments.

An ML framework allowed us to understand these heterogeneous risk factors. Unsupervised techniques were used to define the phenoclusters strictly based on inherent patient features, not based on patient outcomes. Supervised methods were used to ensure that features considered were relevant to outcomes, and later to analyze the importance of features within phenoclusters for predicting outcomes. We identified not only a robust set of sub-groups among COVID-ARDS patients, but also a list of features predictive of outcome within these subgroups, which is distinct from the predictive features for the entire patient set. We anticipate that this process of phenocluster-based risk factor identification will better predict outcomes in patients with COVID-19 ARDS than cohort-based risk assessment. External validation with large cohorts must still be performed and we invite collaborators to apply the open-access algorithms for this purpose.

Most of the variables included in our models are found within electronic health records and represent real-world data – at least within health systems that have the financial means for repeat laboratory testing and vital sign recording. Efforts to decrease heterogeneity among ARDS diagnoses may require more than a binary partition of for example hyper-vs. hypo-inflammatory types, at least in the case of COVID-19 ARDS, and this identification needs large numbers of patients. Importantly, characteristics defining a subgroup do not necessarily align with risk factors for mortality. For example, although beta blockers and calcium channel blockers characterized C1, they were not risk factors for mortality in that phenocluster. Different phenoclusters may have different causative pathways to mortality which require analysis of dimensions beyond laboratory values, to include changes in hemodynamics, demographics, and response to treatments. Further, ARDS biomarker identification may be more successful within phenoclusters.

The features that predicted mortality in one phenocluster but were protective in another could provide insights into different pathophysiology and eventual targeted treatments. For example, an increase in RDW was protective in both C1 and C4 (lower mortality group) but detrimental in C5. RDW has been described as a marker of inflammation and high RDW – as a single measure - has been found to predict mortality in critical illness and in COVID-19 critical illness^13,14^. An increase in RDW might indicate that more blood transfusions were given, although that data was not available^15^. Alternately, an increase in RDW could signal increase in reticulocytes marking a robust myeloid stress response to peripheral inflammatory RBC destruction. In fact, an upregulation of GMCSF (which increases reticulocyte production) has been found in those with increased risk for COVID-19 critical illness^16^. These opposite trends suggest that there is more to explore in uncovering the relationship between RDW and critical illness mortality. Also interesting is that increased FiO2 fluctuations were protective in C5 but predicted increased mortality in the other high mortality phenocluster C2. Were there different parameters for different patient types that were used for FiO2 titration or does one phenocluster require a different approach to FiO2 management for the same hypoxemia parameters?

The anti-hypertensives that have been found as predictors of severe COVID-19 disease and mortality are also found in this analysis as risk factors for the COVID-19 ARDS phenoclusters, although the type of antihypertensive medication that posed an increased risk of in-hospital mortality was different between the phenoclusters. Of note, a recent analysis demonstrated that there was in fact no increased risk of COVID-19 illness and mortality for those using anti-hypertensives and points out that prior studies were erroneous due to collider bias^17^. Our study asks a different question by looking at risk factors for death rather than diagnosis of COVID-19 disease. Therefore, controlling for the underlying disease of hypertension and multiple comorbidities as confounders is appropriate in our analyses and does not pose a risk for collider bias. Additionally, a large recent meta-analysis identified that NSAIDs are not associated with mortality in COVID-19, although several studies do report NSAIDs as a risk factor^18^. In our analysis, only phenoclusters C1 and C2 had NSAIDs as risk factors for mortality. It is very likely that this discrepancy between observational study results is due to heterogeneity of COVID-19 patients included.

Contrary to two other COVID-19 ARDS analyses, our study did not find the inflammatory markers that were available in the dataset (LDH, CRP, ferritin) to differentiate between phenoclusters. Although for the overall cohort elevated LDH and CRP did predict increased mortality, within phenoclusters they were not found to be risk factors. Specifically, Ranjeva *et al*.’s study of 263 ICU ARDS COVID-19 people found that troponin and d-dimer were more elevated^7^, and albumin was lower in the higher mortality group, which was similar to the findings of Sinha and colleagues^8^. However, Ranjeva *et al*. did not find that CRP was different between the two phenoclusters and was around 20mg/dL which is similar to our findings. Notably, our outcome was hospital mortality whereas Sinha used 90-day mortality, which may explain this difference. Additionally, among other nuanced differences between our studies, we included pre-hypoxemia time frame laboratory and vital sign variables in our risk factor identification whereas Sinha et al used values within 24 hours of IMV initiation. In addition, Sinha *et al*. only include hypertension and Diabetes as comorbidities and did not have home medications.

The strengths of our study include the large sample size from a carefully curated database of COVID-19 ARDS patients across multiple hospitals within a healthcare system, with a diverse socio-economic and racial patient mix. This represents the largest real-world data analysis of COVID-19 ARDS patients. Due to the low missingness, many more variables could be included in the analysis, and we were able to include trends over time. We also uniquely included people who met ARDS criteria before intubation unlike other COVID-19 ARDS studies. The examination of steroid response and mortality was also novel.

A previously mentioned, there were some limitations that we tried to address in our methodology. The limitations of our study have to do with the inherent nature of real-world data analytics, particularly when the prospective collection of data was not pre-structured. Potential errors and biases to consider with such a dataset include whether variables represent the actual parameter of interest. To overcome this barrier the NorthCARDS database was designed by a team of front-line clinicians, health information technology specialists, medical informaticists and data science researchers. A further limitation is our lack of external validation with data from another health system. As with almost all hospitalized COVID-19 studies, the duration of illness before hospitalization is not clear and therefore the duration of ARDS pre-treatments for those who presented with ARDS on hospital admission versus those who deteriorated during the admission is not known. This is an important consideration when comparing outcomes and treatment effects. In fact, 687 of 2876 people in the full dataset were intubated within one hour of hospital admission and the duration of severe hypoxemia prior to hospital admission is unknown. Despite these limitations, we believe that our findings contribute to endeavours to decrease the heterogeneity of patients with ARDS and in COVID-19 ARDS.

## Data Availability

All data produced in the present study are available upon reasonable request to the authors

## Registration to Clinical Trials

ClinicalTrials.gov Identifier: NCT04729075 https://clinicaltrials.gov/ct2/show/NCT04729075?term=hajizadeh&draw=2&rank=3

