## Supplemental Methods for "Drivers of Mortality in COVID ARDS Depend on Patient Sub-Type"

#### Data Collection

Relational databases containing EMR data were queried to extract data relevant to this study, and housed in a new relational database. Following this extraction, a data cleaning and preparation pipeline was developed by a group of data scientists in consultation with a team of eight clinicians (NorthCARDS - described previously[1]). The NorthCARDS dataset is a retrospective real-world dataset which includes patients with COVID-19 ARDS as defined by the Berlin criteria, and is grouped into two time periods during the COVID-19 pandemic. The first period, wave 1, consisted of 1,910 hospitalizations from 1,901 distinct patients admitted between Feb. 16$^{th}$, 2020, and Apr. 30$^{th}$, 2020. Wave 1 index hospitalization and re-admission data is available until Dec. 10$^{th}$, 2020. The second period, wave 2, consisted of 966 hospitalizations from 963 distinct patients admitted between Nov. 1$^{st}$, 2020, and Feb. 28$^{th}$, 2021, with follow-up data available until July 16$^{th}$, 2021. All of wave 1 and wave 2 hospitalizations have a final discharge disposition (i.e., their hospital outcomes are known).

Data regarding comorbidities, medications, and smoking status were extracted from anonymized EMRs and annotated in binary format. Demographics used one of 6 racial categories: White, Black, Multiracial, Native American, Asian, and Unknown. Religious status was not used. Age was grouped into four bins ($x<40$, $40<=x<60$, $60<=x<80$, $80<=x$), Charlson Comorbidity Index (CCI) values were converted to probabilities (probability of 10-year survival). Outcomes were organized into discharge disposition, mortality, and mortality in hospital. Vital signs included for this analysis included oxygen saturation as detected by non-invasive pulse oximetry ($SpO_2$), temperature, systolic and diastolic blood pressure, heart rate, respiration rate, pulse pressure (systolic minus diastolic blood pressure), mean arterial pressure, supplemental oxygen concentration ($FiO_2$), and ratio of $SpO_2$ to $FiO_2$ (S:F). The ratio of $PaO_2$ to $FiO_2$ was also included but $PaO_2$ is obtained from laboratory values (from arterial blood gas) and is discussed with the full list of 119 laboratory values (

**Table** S2).

#### Feature Engineering

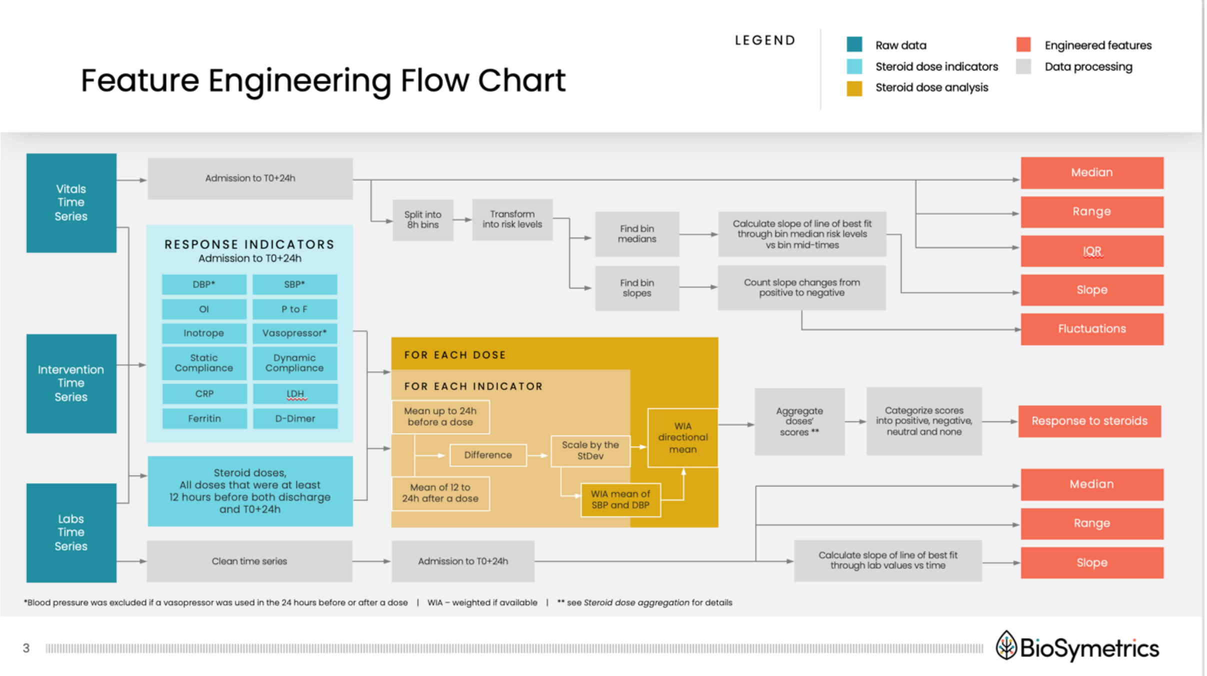


*Figure S1: Feature engineering flow chart. Standard deviations were calculated using only the training data, with the predefined transformation applied to the held-out test data separately. A weighted if available mean is a weighted mean where the weight is 0 if the value is NaN, and 1 if it is defined. The WIA directional mean is the same but with +1 and -1 indicating if an increase or a decrease in value is considered an improvement respectively.*

The vital signs time series for each patient was aggregated into five features, where possible: median, range, inter-quartile range, slope, and fluctuations (**Figure S1**). These features were aggregated from time of admission to 24 hours after Time Zero (T0+24h). Time Zero (T0) indicated the first timepoint where severe hypoxemia or respiratory failure was recorded suggesting the onset of ARDS. This was defined as the earlier time between: 1) First incident of requiring supplemental oxygen with FiO2>=80% for at least four hours, and 2) time of intubation. A 24-hour window after onset of ARDS was chosen to identify clinical features that could ultimately be used to manage patient care and better facilitate clinical trial enrolment.

For vital sign slope calculations, the time series was split into eight-hour bins (time windows), except when there were more than 20 days from admission to T0, in which case admission to T0-20 days (T0 minus 20 days) was treated as one bin (this was only the case for 45 cases). Each value was transformed into a risk level of normal, moderate, and high risk for both decreased and elevated values based on clinical thresholds (for specific thresholds see **Table S1)**. The slope was calculated as the slope of the line of best fit through the median risk level of each eight-hour bin against the midpoint of each bin's time window (**Figure S2**). We also captured the number of fluctuations for a given vital sign by calculating the number of changes from positive slope to negative slope, or vice versa (**Figure S3**).

*Table S1: Vitals risk levels. Vitals measurements were discretized according to the table before aggregating into slope and fluctuations.*

| | High risk, lowered (-2) | Moderate risk, lowered (-1) | Normal (0) | Moderate risk, elevated (1) | High risk, elevated (2) | Highest risk, elevated (3) |
|---|---|---|---|---|---|---|
| SpO2 | (0, 84] | (84, 94] | (94, 100] | | | |
| Temperature | (30, 34] | (34, 36] | (36, 37.5] | (37.5, 40] | (40, 45] | |
| Systolic pressure | (0, 80] | (80, 100] | (100, 130] | (130, 150] | (150, 1000] | |
| Diastolic pressure | (0, 40] | (40, 60] | (60, 80] | (80, 90] | (90, 1000] | |
| Heart rate | (0, 50] | (50, 60] | (60, 110] | (110, 180] | (180, 1000] | |
| Respiratory rate | (0, 8] | (8, 12] | (12, 24] | (24, 35] | (35, 200] | |
| FiO2 | | | (21, 21] | (21, 60] | (60, 80] | (80, 100] |
| *SpO2:FiO2 ratio* | (0, 100] | (100, 300] | (300, 477] | | | |
| *Pulse pressure (SBP-DBP)* | (0, 25] | (25, 30] | (30, 40] | (40, 60] | (60, 1000] | |
| Mean Arterial Pressure | (0, 55] | (55, 65] | (65, 75] | (75, 100] | (100, 1000] | |

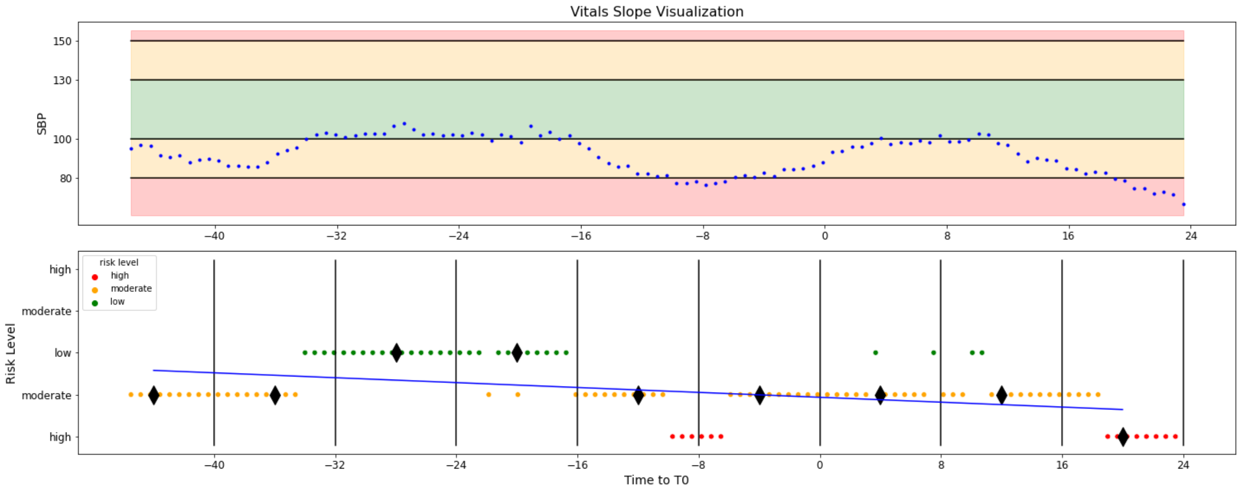


*Figure S2: Vitals Slopes Visualization: The raw measurements are shown plotted against the time of the measurement relative to T0, the time of onset of hypoxemia. They are split into horizontal segments representing the risk levels. In this example the patient was in three of five possible ranges: low risk, moderate risk due to decreased value, and high risk due to decreased value; their values were never elevated. These risk levels are then plotted on the lower figure, the coloured points. These are then split up into 8-hour time windows where the median of the risk values is calculated, shown as black diamonds. Finally, the line of best fit through the medians is calculated and the slope of that line is the slope for that vital for that patient.*

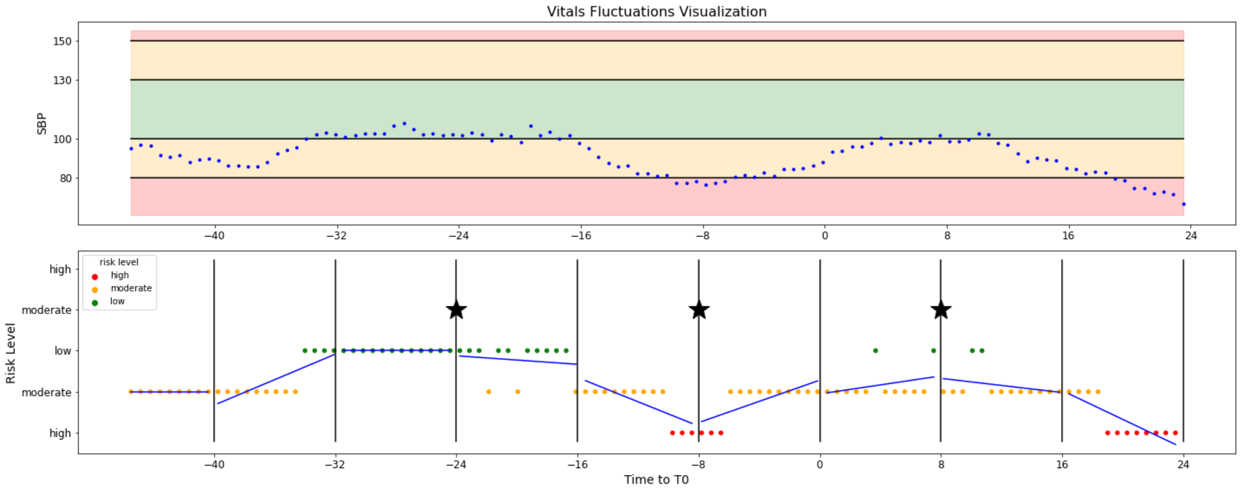


*Figure S3: Vitals Fluctuations Visualization: This starts the same way as the slopes, but rather than calculating the medians in each time window the lines of best fit within each window are calculated. Finally, number of times that the slopes change from positive to negative, or vice versa, are counted, ignoring flat time windows. These changes are marked with a star. The number of such changes is the value for fluctuations for that vital for that patient.*

For laboratory values, numeric values were first extracted from EMR exports by removing units and other strings. For example, recordings often included inequalities, such as “<100”. For these cases it was assumed that the value could be anything up to this boundary value, so the number in the inequality was used (e.g., <100 was taken as 100). After processing, there were 119 numeric lab fields used in our analysis (**Table S2**). The time series for each lab field, for a given patient, were aggregated into three features to summarize lab values from admission to T0+24h. These three aggregated lab features were: median, range, and slope (**Figure S1**). Slope was calculated as the slope of the line of best fit through the lab values plotted against observation time.

*Table S2: All 119 lab tests that were considered, plus dynamic and static compliance as these were populated similarly to labs and P to F ratio as it depends on pO2 values, are listed here. Only medians and ranges that had at least 66.6% of values populated and 55% of slopes populated in the whole dataset, as well as 60% of median and ranges, and 50% of slopes in each wave populated were considered, all other features were excluded for lack of data. Numbers of valid, used observations are listed; values that were below the thresholds are not listed. The total possible number of values is 2876 – this includes both the train and test data. Three aggregations for 119 lab tests gives 357 possible lab features; however, exclusion due to lack of data results in 164 lab features remaining (including 6 compliance values). Further all three stats were kept for labs marked with an asterisk because of a known interest in them.*

| Laboratory Test | Number of Valid Observations | | |
|---|---|---|---|
| | Median | Range | Slope |
| ABS CD3 | --- | --- | --- |
| Activated Partial Thromboplastin Time | 2110 | 2110 | --- |
| Alanine Aminotransferase (ALT/SGPT) | 2867 | 2867 | 2468 |
| Albumin, Serum | 2868 | 2868 | 2482 |
| Alkaline Phosphatase, Serum | 2869 | 2869 | 2474 |
| Anion Gap, Serum | 2873 | 2873 | 2687 |
| Aspartate Aminotransferase (AST/SGOT) | 2867 | 2867 | 2472 |
| Auto Basophil # | 2853 | 2853 | 1975 |
| Auto Basophil % | 2853 | 2853 | 1975 |
| Auto Eosinophil # | 2851 | 2851 | 1972 |
| Auto Eosinophil % | 2853 | 2853 | 1976 |
| Auto Immature Granulocyte % | 2486 | 2486 | --- |
| Auto Lymphocyte # | 2853 | 2853 | 1975 |
| Auto Lymphocyte % | 2853 | 2853 | 1978 |
| Auto Monocyte # | 2853 | 2853 | 1975 |
| Auto Monocyte % | 2853 | 2853 | 1978 |
| Auto Neutrophil # | 2853 | 2853 | 1976 |
| Auto Neutrophil % | 2853 | 2853 | 1977 |
| Bacteria | --- | --- | --- |
| Band Neutrophils % | --- | --- | --- |
| Base Excess, Arterial | --- | --- | --- |
| Base Excess, Venous | --- | --- | --- |
| Bilirubin Direct, Serum | --- | --- | --- |
| Bilirubin Total, Serum | 2869 | 2869 | 2476 |
| Blood Gas Arterial - Calcium, Ionized | --- | --- | --- |
| Blood Gas Arterial - Chloride | --- | --- | --- |
| Blood Gas Arterial - FIO2 | --- | --- | --- |
| Blood Gas Arterial - Glucose | --- | --- | --- |
| Blood Gas Arterial - Hematocrit | --- | --- | --- |
| Blood Gas Arterial - Hemoglobin | --- | --- | --- |
| Blood Gas Arterial - Lactate | --- | --- | --- |
| Blood Gas Arterial - Lytes,Hgb,iCa,Lact Result | --- | --- | --- |
| Blood Gas Arterial - Potassium | --- | --- | --- |
| Blood Gas Arterial - Sodium | --- | --- | --- |

| Laboratory Test | Number of Valid Observations | | |
|---|---|---|---|
| | Median | Range | Slope |
| Blood Gas Arterial, Lactate | --- | --- | --- |
| Blood Gas Calcium, Ionized - Venous | --- | --- | --- |
| Blood Gas Comments | --- | --- | --- |
| Blood Gas Comments Arterial | --- | --- | --- |
| Blood Gas Venous - Chloride | --- | --- | --- |
| Blood Gas Venous - Glucose | --- | --- | --- |
| Blood Gas Venous - Lactate | --- | --- | --- |
| Blood Gas Venous - Potassium | --- | --- | --- |
| Blood Gas Venous - Sodium | --- | --- | --- |
| Blood Urea Nitrogen, Serum | 2873 | 2873 | 2688 |
| C-Reactive Protein, Serum* | 2520 | 2520 | 1197 |
| CD3 % | --- | --- | --- |
| Calcium, Total Serum | 2873 | 2873 | 2686 |
| Carbon Dioxide, Serum | 2873 | 2873 | 2687 |
| Chloride, Serum | 2873 | 2873 | 2688 |
| Creatine Kinase, Serum | --- | --- | --- |
| Creatinine, Serum | 2873 | 2873 | 2706 |
| D-Dimer Assay, Quantitative* | 2269 | 2269 | 1024 |
| Epithelial Cells | --- | --- | --- |
| FIO2, Arterial | --- | --- | --- |
| Ferritin, Serum* | 2453 | 2453 | 1093 |
| Fibrinogen Assay | --- | --- | --- |
| Glucose Qualitative, Urine | --- | --- | --- |
| Glucose, Serum | 2873 | 2873 | 2687 |
| HCO3, Arterial | --- | --- | --- |
| HCO3, Venous | --- | --- | --- |
| Haptoglobin, Serum | --- | --- | --- |
| Hematocrit | 2873 | 2873 | 2619 |
| Hematocrit, Calculated | --- | --- | --- |
| Hemoglobin | 2874 | 2874 | 2620 |
| IANC | --- | --- | --- |
| INR | 2146 | 2146 | --- |
| Indirect Reacting Bilirubin | --- | --- | --- |
| Ketone - Urine | --- | --- | --- |
| Lactate Dehydrogenase, Serum* | 1945 | 1945 | 626 |
| Lactate, Blood | --- | --- | --- |
| MPV | --- | --- | --- |
| Magnesium, Serum | 2300 | 2300 | --- |
| Mean Cell Hemoglobin | 2873 | 2873 | 2618 |
| Mean Cell Hemoglobin Conc | 2873 | 2873 | 2619 |

| Laboratory Test | Number of Valid Observations | | |
|---|---|---|---|
| | Median | Range | Slope |
| Mean Cell Volume | 2873 | 2873 | 2618 |
| Metamyelocytes % | --- | --- | --- |
| Myelocytes % | --- | --- | --- |
| Nucleated RBC | 2160 | 2160 | 1736 |
| Nucleated RBC # | --- | --- | --- |
| Oxygen Content | --- | --- | --- |
| Oxygen Saturation, Arterial | --- | --- | --- |
| Oxygen Saturation, Venous | --- | --- | --- |
| POCT Blood Glucose. | 1990 | 1990 | 1670 |
| Phosphorus Level, Serum | 1949 | 1949 | --- |
| Platelet Count - Automated | 2874 | 2874 | 2617 |
| Potassium, Serum | 2871 | 2871 | 2681 |
| Procalcitonin, Serum | 2415 | 2415 | --- |
| Protein Total, Serum | 2867 | 2867 | 2476 |
| Protein, Urine | --- | --- | --- |
| Prothrombin Time, Plasma | 2146 | 2146 | --- |
| RBC Count | 2874 | 2874 | 2619 |
| Reactive Lymphocytes % | --- | --- | --- |
| Red Blood Cell - Urine | --- | --- | --- |
| Red Cell Distrib Width | 2873 | 2873 | 2619 |
| Sedimentation Rate, Erythrocyte | --- | --- | --- |
| Serum Pro-Brain Natriuretic Peptide | --- | --- | --- |
| Sodium, Serum | 2873 | 2873 | 2688 |
| Specific Gravity | --- | --- | --- |
| Thyroid Stimulating Hormone, Serum | --- | --- | --- |
| Total CO2, Arterial | --- | --- | --- |
| Total Hemoglobin, Calculated | --- | --- | --- |
| Triglycerides, Serum | --- | --- | --- |
| Troponin I, Serum | --- | --- | --- |
| Troponin T, High Sensitivity Result | --- | --- | --- |
| Troponin T, Serum | --- | --- | --- |
| Urobilinogen | --- | --- | --- |
| Vancomycin Level, Trough | --- | --- | --- |
| WBC Count | 2874 | 2874 | 2620 |
| White Blood Cell - Urine | --- | --- | --- |
| eGFR if African American | 2873 | 2873 | 2706 |
| eGFR if Non African American | 2873 | 2873 | 2706 |
| pCO2, Arterial* | 1835 | 1835 | 1275 |
| pCO2, Venous* | 1552 | 1552 | 383 |
| pH Urine | --- | --- | --- |

| Laboratory Test | Number of Valid Observations | | |
|---|---|---|---|
| | Median | Range | Slope |
| pH, Arterial* | 1646 | 1646 | 1171 |
| pH, Blood | --- | --- | --- |
| pH, Venous* | 1552 | 1552 | 386 |
| pO2, Arterial* | 1836 | 1836 | 1276 |
| pO2, Venous* | 1551 | 1551 | 386 |
| Dynamic_compliance* | 982 | 982 | 705 |
| Static_compliance* | 889 | 889 | 631 |
| PtoF | --- | --- | --- |

To evaluate steroid response, we identified timing for changes suggestive of improved clinical status which we defined as removal from vasopressor or inotrope, improved oxygenation (Oxygenation Index, (OI) and P:F), improved lung compliance, increased blood pressure (if not on a vasopressor), and decrease in CRP, LDH, D-dimer, and Ferritin (**Figure S1** diagrams this process). Specifically, the mean of each of these values for up to 24 hours before receiving steroids was compared to the mean values for at least 12 hours (and up to 24 hours) after each dose, and the difference was calculated. The differences were scaled to have unit standard deviation across all differences where there were values available to compare. The scaling was fit to only the training data from the COVID-19 ARDS dataset (see Pre-processing section below). Diastolic and Systolic blood pressure were averaged to create a single indicator. The response to each indicator was combined in a directional mean, based on direction of change. Therefore, we derived one response level for each steroid dose. We calculated a weighted average to aggregate steroid response over multiple doses, factoring in varying responses for different doses, and that one strong positive response would indicate a good response compared to multiple non-responses, Specifically, if x is a list of the n scores for a patient sorted in decreasing order, then the weighted score was calculated as

$$s = \frac{x_1}{2^n} + \sum_{j=1}^{n} \frac{x_j}{2^j}$$

This decreasing weighting was done to give the heaviest weight to the strongest improvement around a steroid dose. Whether a positive response was after the first dose, or a cumulative effect seen after multiple doses, it would be considered a positive response. Finally, the scores were transformed to categorize patients based on response to steroids, using a threshold of ±0.25, where a ‘positive’ label was applied for s>0.25, ‘negative’ for s<-0.25, ‘neutral’ for $-0.25 \leq s \leq 0.25$, and ‘none’ if *s* could not be calculated. ‘None’ meant that there were no indicators of steroid response available in the data for a given time interval because steroids were not administered at the onset of ARDS (using hypoxemia threshold definition). This missingness was seen among 45% of the patients in the examined time period. Note that only steroid response up to the first 24 hours after ARDS onset were examined, because the goal was to enable prospective identification of the phenotypic subgroup.

#### Steroid dose aggregation example

Say a patient received 4 doses in the relevant timeframe, and they have the directional means, ordered by time of dose: -0.2, -0.8, -0.3, 2.1 – based on the first three doses the patient’s condition is deteriorating, despite the steroids, but on the 4th dose the positive impact is more drastic than the prior deterioration. First these scores are ordered from largest to smallest: 2.1, -0.2, -0.3, -0.8 and the following formula is applied:

$$s = \frac{x_1}{2^n} + \sum\nolimits_{j=1}^{n} \frac{x_j}{2^j}$$

This calculates a weighted mean, where the largest value receives the largest weight. For our example we would have:

$$s = \frac{2.1}{16} + \frac{2.1}{2} + \frac{-0.2}{4} + \frac{-0.3}{8} + \frac{-1.5}{16} = 0.9125$$

These weights give the relevant importance to the positive responses so that the metric recognizes that this is a patient that did respond to the cumulative effect of these steroid doses. If a simple mean had been used the value would have been completely neutral at 0, and indistinguishable from the case of one strong negative response and multiple slightly positive responses: -2.1, 0.2, 0.3, 1.5. Whereas the weighted mean would give this opposite case a value of 0.6125, not as strong as the first response, but still a positive response.

#### Pre-processing

Features were either categorical or continuous. Categorical features included demographics, smoking, outcomes, comorbidities, and home medications. Categorical features with only two categories were encoded into one Boolean feature. Those with more than two categories were encoded into *n* Boolean features. There were no missing categorical variables in this dataset, although race and smoking status, ventilator type and religion had an "unknown" label in the electronic medical records which was reflected in an 'unknown' category. Continuous features, including laboratory values, were all standardized to have a mean of 0 and a standard deviation of 1 to ensure consistent contribution to models by features which have a variety of inherent scales. Unlike the categorical features, there were missing values for some continuous features. These missing values were imputed to the median value within the training dataset.

#### Feature Selection

For each of the four discharge dispositions (expired, discharged home, discharged to facility intubated, and discharged to facility extubated) a predictive model was built as follows. Models for all four discharge dispositions were investigated to understand overlapping or distinct feature sets. First, a binary vector was created (i.e., one-versus-others for each discharge disposition value) and used as model targets. Next, from the 268 total available features 1000 different randomly sampled feature sets were selected as predictors for regression models. Logistic regression models with LASSO regularization were then trained using each predictor set and each model, then evaluated using 5-fold cross-validation. This type of model only includes the most predictive subset of the sampled features (excluding the rest). Subsequently, features were ranked based on the proportion of times that each was included relative to the total number of times that it was sampled for each model. The feature importance ranking of all four models was then filtered based on multiple minimum cut-off levels to select features providing the most information about the discharge dispositions. Which cut-off level to select features was selected in conjunction with other parameters (explained variance and number of clusters) in the model selection step detailed below.

#### Patient clustering and cluster comparison

To facilitate clustering and further separate signal from noise within the filtered feature set, Principal Component Analysis (PCA) was performed on the features derived from feature selection. K-means clustering, an unsupervised algorithm that assigns each observation to the cluster with the nearest mean or centroid, was used to group patients based on principal components outputted from the PCA. Models were fit for a variety of feature selection cut-off thresholds, PCA explained variance levels and number of K-means clusters were examined. The selected model parameters were chosen simultaneously because they are interdependent. A three-step process was implemented to select the final model. It primarily prioritized models that have clusters with distinct mortality characteristics as well as clusters that are distinguished by a variety of other characteristics of interest. This process is detailed in **Table S3**.

*Table S3: Model selection iterations*

| | | |
|---|---|---|
| **Step 1:** | | |
| | Run on all 5-training folds | |
| | Used all permutations of the following two sets of parameters<br>- (cut-off: 75%, 85%, 95%; explained variance: 50%, 70%, 90%; number of clusters: 7,10,13) to guide parameter selection to areas of interest: higher cut-off and lower explained variance.<br>- (cut-off: 95%, 96%, 97%, 98%; explained variance: 25%, 30%, 33%, 40%, 50%; number of clusters: 7,8,9,10,11,12,13) | |
| | Models were ranked based on the number of clusters with mortality enrichment across the minimum, median and maximum of the 5-folds in both the training fold and the validation fold. Ranking was done in the following order:<br>Validation fold median (for consistency, avoiding “overfitting”), train fold median, train fold minimum, then train fold maximum | |
| | 33 models were selected in this step based on having a validation fold median of at least 1 cluster enriched (note that because of the smaller test sample sizes the threshold for significant differences is higher than for the larger train folds), a train fold median of at least 3 clusters enriched, and a train fold minimum of at least 2 clusters enriched | |
| **Step 2:** | | |
| | The 33 models from step 1 were as follows, (cut-off, explained variance, number of clusters):<br>(96%, 33%, 10), (96%, 33%, 12), (97%, 30%, 10), (97%, 33%, 10), (96%, 33%, 11), (96%, 33%, 13), (97%, 33%, 9), (96%, 33%, 9), (97%, 30%, 11), (95%, 40%, 13), (96%, 33%, 7), (96%, 40%, 12), (97%, 30%, 12), (96%, 33%, 8), (97%, 33%, 12), (97%, 33%, 11), (98%, 40%, 12), (96%, 50%, 12), (96%, 50%, 10), (96%, 50%, 11), (97%, 33%, 8), (97%, 30%, 8), (96%, 40%, 7), (97%, 40%, 13), (98%, 40%, 13), (96%, 50%, 7), (96%, 50%, 8), (96%, 50%, 9), (97%, 40%, 12), (97%, 40%, 8), (97%, 50%, 7), (98%, 40%, 11), (98%, 50%, 13). | |
| | Models for the above parameters were run on the 5 training folds and compared amongst themselves as follows:<br>- For each attribute group {labels, features} | |
| | | For each model characteristic {# components, # clusters, explained variance, cut-off}<br>- Plot the median number of clusters with enrichments of the attribute group against the model characteristic; calculate each model’s residual from the line of best fit; scale each residual by the standard deviation of the set of residuals – call this the absolute residuals<br>- Plot the median number of clusters with enrichments relative to the total number of clusters of the attribute group against the model characteristic; calculate each model’s residual from the line of best fit; scale each residual by the standard deviation of the set of residuals – call this the relative residuals<br>- Plot the relative residuals against the absolute residuals and for the points in the first quadrant (those that outperformed expectation in both the absolute and relative comparison) calculate their radius and rank them against each other. |
| | | Based on the radii and rankings calculate the following 3 metrics for each model:<br>- Score: points for were allocated based on rankings, 20 points for 1$^{st}$, decreasing by one point per placing; the score metric is the sum of the points based on the individual attribute group, model characteristic pair rankings. The maximum possible is 160.<br>- Count: the number of attribute group, model characteristic pairs for hich the model was in the first quadrant, the maximum possible is 8. |

| | | |
|---|---|---|
| | | - Sum: the sum of the radii when the model was in the first quadrant. There is technically no upper bound, but the standardization of the residuals limits the extreme values. |
| | | Models are selected by choosing the top few models based on the three metrics. Here two were chosen: (96%, 33%, 10) and (96%, 33%, 12) |
| | | Before moving on to the next step the selections are confirmed to be valid choices based on:<br>- Small clusters: The number of small clusters (less than (10/number of clusters)% of data) not more than 2<br>- Small clusters: Percent of small clusters is less that 20%<br>- Dominant clusters: No clusters that contain more than (300/number of clusters)% of data<br>- Overfitting: No discrepancies between the average proportion of data in each cluster from train fold to test fold greater than 5% |
| **Step 3:** | | |
| | Fit the selected two models to the whole train set. The goal of this step is to select one model, ensuring consistency from the 5 folds to the whole training set. | |
| | Both the models that were selected used 63 features (96% cut-off) reduced to 5 components (33% explained variance), as was the mean number of components across the 5 folds. | |
| | Comparing the proportion of data in mortality enriched clusters: | |
| | | The 10 cluster model has 35% of hospitalizations in low mortality clusters, and 30% in the high mortality clusters for a total of 65% of hospitalizations in mortality enriched clusters. |
| | | The 12 cluster model has 29% of hospitalizations in low mortality clusters, and 21% in the high mortality clusters for a total of 50% of hospitalizations in mortality enriched clusters |
| | Comparing total label and feature enrichment: | |
| | | The 10 cluster model has label enrichment in 8 clusters across 20 labels. It has feature enrichment in all 10 clusters across 50 features. |
| | | The 12 cluster model has label enrichment in 11 clusters across 19 labels. It has feature enrichment in all 12 clusters across 52 features. |
| | Based on the cluster enrichments there is not an obvious reason to choose one model over the other, but the proportion of hospitalizations in a mortality enriched cluster is notably higher in the 10 cluster model than in the 12 cluster model, that is the main reason that 10 is selected over 12 clusters. As further reasoning in the 12 cluster model 33% of clusters have mortality enrichment compared to 40% in the 10 cluster model. Moderately small clusters: 5 clusters in the 12 cluster model have less than 100 hospitalizations compared to 3 in the 10 cluster model. | |

#### Analysis of Factors Driving Mortality

Once the clusters were identified, to understand the driving factors behind mortality rates for each cluster, an L1 regularized (LASSO) regression was performed. First, the hospitalizations in each cluster were stratified into five folds based on mortality. Cross validation was performed to select the ideal regularization strength based on both model performance (i.e., ROC AUC) and a small (5 to 20) number of driving factors. Once regularization strength was selected, the model was fit to the whole cluster from the training data to extract the feature set that defined the driving factors for mortality within that cluster. With the hypothesis that some features would not be predictive when applied to the non-clustered dataset this process was also performed for the full training dataset, not segmented by cluster assignment. Regression models were implemented with scikit-learn using mortality as the target, and the same 63 selected features used for clustering as predictors. Weights of features were compared between cluster sets and compared to the weights of features for the full dataset to understand the similarities and differences in risk factors in the each phenocluster and in the whole cohort.

*Table S4: Categorization of Selected Features. * Cystic_fibrosis_0 indicates that a patient did not have CF since none of the patients in our dataset had CF. Since the feature was composed of all 1's the LASSO models in feature selection regularly included it as part of the constant. It was not allowed to contribute to the phenocluster definitions though because of the preliminary step of PCA feature reduction. A constant feature has zero variance, so it is entirely represented in the component with the smallest explained variance and by limiting the total explained variance it is excluded entirely.*

| Feature Type | | Number | Features |
|---|---|---|---|
| Demographics | | 3+2 | Gender: Male, Race: Asian, Race: Black or African-American, Smoking Status: Non-Smoker, Smoking Status: Unknown |
| Comorbidities | | 6+1 | Atrial Fibrillation, Autoimmune Disorder, Diabetes Mellitus, Dementia or Neurological, Interstitial Lung Disease, Pulmonary Hypertension, Cystic Fibrosis 0* |
| Home Medications | | 6 | ACE Inhibitors, Immunomodulator Suppressants, Inhaled Pulmonary Therapies, NSAIDs, Beta Blockers, Calcium Channel Blockers |
| Labs | Median | 12 | Albumin, Eosinophil %, Immature Granulocyte %, Bilirubin Total, Blood Urea Nitrogen, C-Reactive Protein, Calcium Total, Glucose, Lactate Dehydrogenase, eGFR (non-AA), pCO2, Venous, pH, Arterial |
| | Range | 12 | Albumin, Basophil %, Eosinophil %, Immature Granulocyte %, Lymphocyte %, Neutrophil #, C-Reactive Protein, Chloride, Phosphorus Level, Potassium, Procalcitonin, pO2, Arterial |
| | Slope | 6 | Eosinophil %, Monocyte %, Bilirubin Total, Glucose, Red Cell Distrib. Width, pH, Venous |
| Vitals | Median | 2+1 | Pulse Pressure, Respiration Rate, Dynamic Compliance |
| | Range | 1+1 | Systolic Pressure, FiO2 |
| | IQR | 1 | Diastolic Pressure |
| | Slope | 2 | Mean Arterial Pressure, SpO2 |
| | Fluctuations | 5+1 | Diastolic Pressure, Heart Rate, Respiration Rate, SpO2, S to F, FiO2 |
| Steroid Response | | 1 | Steroid Response: Neutral |

#### Means with Confidence Intervals

Means and 90% confidence intervals for the mean in the whole training set and among each phenocluster within the training set. No missing values are imputed nor are the values shown here standardized. Numeric features represent that value, binary features are the proportion of the set that fall into that category.

Feature Selected "N/A" means that it was not included in the feature selection process by design; "yes" means that the feature was selected by feature selection and therefore was clustered on and included in the feature importance analysis; if there is no value listed the feature was included in feature selection but was not selected and therefore not included in analysis after that.

##### Outcomes

*Table S5a: Outcomes - means and 90% confidence intervals for the whole training set and among each phenocluster. Days values are in days, all other values are the proportion of the set that fall into that category.*

| Outcomes | Feature Selected | all | C 1 | C 2 | C 3 | C 4 | C 5 | C 6 | C 7 | C 8 | C 9 | C10 |
|---|---|---|---|---|---|---|---|---|---|---|---|---|
| **Days Admitted** | N/A | **22.7531 (22.0667, 23.4395)** | 28.3605 (26.8864, 29.8346) | 21.1714 (19.7752, 22.5676) | 20.8906 (19.2232, 22.5579) | 20.2201 (17.9176, 22.5227) | 16.0679 (14.1857, 17.9501) | 23.1623 (21.0078, 25.3169) | 29.0348 (26.1931, 31.8765) | 14.3966 (11.0463, 17.7469) | 28.3427 (23.6632, 33.0222) | 14.7831 (9.0965, 20.4696) |
| **Days Intubated** | N/A | **13.5973 (13.0774, 14.1173)** | 16.5989 (15.4912, 17.7067) | 12.1096 (10.9540, 13.2652) | 14.0924 (12.7511, 15.4338) | 14.4300 (12.7916, 16.0683) | 10.0877 (8.5098, 11.6655) | 13.9077 (12.3029, 15.5126) | 12.8006 (10.5955, 15.0057) | 11.4283 (8.7505, 14.1060) | 9.9345 (7.3756, 12.4934) | 4.0652 (2.0792, 6.0512) |
| **Discharge Disposition: Expired** | N/A | **0.7443 (0.7294, 0.7591)** | 0.6181 (0.5844, 0.6518) | 0.8212 (0.7924, 0.8500) | 0.7723 (0.7340, 0.8106) | 0.6298 (0.5806, 0.6789) | 0.8922 (0.8587, 0.9258) | 0.8160 (0.7722, 0.8599) | 0.7522 (0.6851, 0.8193) | 0.8214 (0.7523, 0.8906) | 0.7000 (0.6093, 0.7907) | 0.8750 (0.6694, 1.0806) |
| **Discharge Disposition: Facility Extubated** | N/A | **0.0932 (0.0833, 0.1031)** | 0.0959 (0.0755, 0.1163) | 0.0977 (0.0754, 0.1200) | 0.0769 (0.0526, 0.1013) | 0.1374 (0.1024, 0.1725) | 0.0603 (0.0346, 0.0861) | 0.0849 (0.0533, 0.1165) | 0.1416 (0.0874, 0.1958) | 0.0000 (0.0000, 0.0357) | 0.1286 (0.0623, 0.1949) | 0.0000 (0.0000, 0.3750) |
| **Discharge Disposition: Facility Intubated** | N/A | **0.0357 (0.0294, 0.0420)** | 0.0337 (0.0212, 0.0463) | 0.0333 (0.0198, 0.0467) | 0.0615 (0.0396, 0.0835) | 0.0420 (0.0216, 0.0624) | 0.0129 (0.0007, 0.0252) | 0.0047 (0.0000, 0.0125) | 0.0265 (0.0016, 0.0515) | 0.0833 (0.0334, 0.1332) | 0.0571 (0.0112, 0.1031) | 0.0000 (0.0000, 0.3750) |

| Outcomes | Feature Selected | all | C 1 | C 2 | C 3 | C 4 | C 5 | C 6 | C 7 | C 8 | C 9 | C10 |
|---|---|---|---|---|---|---|---|---|---|---|---|---|
| **Discharge Disposition: Home** | N/A | **0.1268 (0.1155, 0.1381)** | 0.2522 (0.2221, 0.2824) | 0.0478 (0.0318, 0.0638) | 0.0892 (0.0632, 0.1153) | 0.1908 (0.1508, 0.2308) | 0.0345 (0.0147, 0.0542) | 0.0943 (0.0612, 0.1274) | 0.0796 (0.0376, 0.1217) | 0.0952 (0.0422, 0.1482) | 0.1143 (0.0513, 0.1773) | 0.1250 (0.0000, 0.3306) |
| **Outcome: Resolved** | N/A | **0.2532 (0.2384, 0.2679)** | 0.3801 (0.3464, 0.4138) | 0.1746 (0.1461, 0.2031) | 0.2277 (0.1894, 0.2660) | 0.3702 (0.3211, 0.4194) | 0.1034 (0.0705, 0.1364) | 0.1840 (0.1401, 0.2278) | 0.2301 (0.1647, 0.2955) | 0.1786 (0.1094, 0.2477) | 0.3000 (0.2093, 0.3907) | 0.1250 (0.0000, 0.3306) |
| **Outcome (in hosp): Expired After** | N/A | **0.0026 (0.0008, 0.0043)** | 0.0018 (0.0000, 0.0047) | 0.0042 (0.0000, 0.0090) | 0.0000 (0.0000, 0.0092) | 0.0000 (0.0000, 0.0115) | 0.0043 (0.0000, 0.0114) | 0.0000 (0.0000, 0.0142) | 0.0177 (0.0000, 0.0382) | 0.0000 (0.0000, 0.0357) | 0.0000 (0.0000, 0.0429) | 0.0000 (0.0000, 0.3750) |
| **Outcome (in hosp): Expired in Hosp** | N/A | **0.7443 (0.7294, 0.7591)** | 0.6181 (0.5844, 0.6518) | 0.8212 (0.7924, 0.8500) | 0.7723 (0.7340, 0.8106) | 0.6298 (0.5806, 0.6789) | 0.8922 (0.8587, 0.9258) | 0.8160 (0.7722, 0.8599) | 0.7522 (0.6851, 0.8193) | 0.8214 (0.7523, 0.8906) | 0.7000 (0.6093, 0.7907) | 0.8750 (0.6694, 1.0806) |
| **Outcome (in hosp): Resolved** | N/A | **0.2532 (0.2384, 0.2679)** | 0.3801 (0.3464, 0.4138) | 0.1746 (0.1461, 0.2031) | 0.2277 (0.1894, 0.2660) | 0.3702 (0.3211, 0.4194) | 0.1034 (0.0705, 0.1364) | 0.1840 (0.1401, 0.2278) | 0.2301 (0.1647, 0.2955) | 0.1786 (0.1094, 0.2477) | 0.3000 (0.2093, 0.3907) | 0.1250 (0.0000, 0.3306) |

#### Demographics

*Table S5b: Demographics - means and 90% confidence intervals for the whole training set and among each phenocluster. BMI and captured comorbidity count represent that value, CCI and MEWS are the probability transformation of those scores, all other values are the proportion of the set that fall into that category.*

| Demographics | Feature Selected | all | C 1 | C 2 | C 3 | C 4 | C 5 | C 6 | C 7 | C 8 | C 9 | C10 |
|---|---|---|---|---|---|---|---|---|---|---|---|---|
| **BMI** | | **29.8691 (29.5868, 30.1514)** | 30.0288 (29.4809, 30.5767) | 30.8428 (30.2115, 31.4742) | 29.4602 (28.7096, 30.2108) | 30.4631 (29.5088, 31.4174) | 29.4626 (28.5632, 30.3619) | 28.5519 (27.7318, 29.3721) | 28.6190 (27.1827, 30.0552) | 30.0021 (28.3946, 31.6095) | 28.4991 (27.0787, 29.9194) | 33.1438 (27.5786, 38.7091) |
| **Captured Comorbidity Count** | | **2.9800 (2.9122, 3.0478)** | 2.2700 (2.1576, 2.3824) | 3.4324 (3.2852, 3.5797) | 2.8523 (2.6748, 3.0299) | 2.7214 (2.5441, 2.8987) | 4.4784 (4.2367, 4.7202) | 2.2358 (2.0588, 2.4129) | 3.7522 (3.4077, 4.0967) | 2.5000 (2.1817, 2.8183) | 3.7857 (3.3520, 4.2194) | 2.7500 (1.9424, 3.5576) |
| **CCI** | | **0.3718 (0.3590, 0.3847)** | 0.5836 (0.5587, 0.6086) | 0.2200 (0.1981, 0.2419) | 0.4057 (0.3711, 0.4403) | 0.4504 (0.4109, 0.4900) | 0.0752 (0.0549, 0.0954) | 0.4878 (0.4458, 0.5298) | 0.1845 (0.1379, 0.2311) | 0.3854 (0.3198, 0.4509) | 0.2071 (0.1381, 0.2762) | 0.1225 (0.0000, 0.2805) |

| Demographics | Feature Selected | all | C 1 | C 2 | C 3 | C 4 | C 5 | C 6 | C 7 | C 8 | C 9 | C10 |
|---|---|---|---|---|---|---|---|---|---|---|---|---|
| **MEWS** | N/A | **0.1864 (0.1825, 0.1903)** | 0.1664 (0.1591, 0.1737) | 0.2046 (0.1950, 0.2143) | 0.1875 (0.1776, 0.1973) | 0.1887 (0.1778, 0.1997) | 0.1920 (0.1791, 0.2049) | 0.1907 (0.1784, 0.2029) | 0.1832 (0.1586, 0.2077) | 0.2122 (0.1933, 0.2312) | 0.1776 (0.1495, 0.2057) | 0.2015 (0.1066, 0.2964) |
| **Age Group: 20-39** | | **0.0353 (0.0291, 0.0416)** | 0.0728 (0.0548, 0.0909) | 0.0062 (0.0003, 0.0121) | 0.0185 (0.0062, 0.0308) | 0.0687 (0.0429, 0.0945) | 0.0129 (0.0007, 0.0252) | 0.0283 (0.0095, 0.0471) | 0.0265 (0.0016, 0.0515) | 0.0119 (0.0000, 0.0315) | 0.0286 (0.0000, 0.0616) | 0.0000 (0.0000, 0.3750) |
| **Age Group: 40-59** | | **0.2387 (0.2243, 0.2532)** | 0.3748 (0.3412, 0.4084) | 0.0915 (0.0698, 0.1131) | 0.2215 (0.1836, 0.2595) | 0.3092 (0.2621, 0.3562) | 0.1336 (0.0968, 0.1704) | 0.2736 (0.2231, 0.3241) | 0.1770 (0.1177, 0.2363) | 0.2500 (0.1718, 0.3282) | 0.3143 (0.2224, 0.4062) | 0.1250 (0.0000, 0.3306) |
| **Age Group: 60-79** | | **0.5634 (0.5466, 0.5802)** | 0.4956 (0.4609, 0.5303) | 0.6486 (0.6128, 0.6845) | 0.6092 (0.5646, 0.6538) | 0.4847 (0.4338, 0.5356) | 0.5560 (0.5023, 0.6098) | 0.6368 (0.5823, 0.6913) | 0.5310 (0.4534, 0.6085) | 0.5952 (0.5066, 0.6839) | 0.4143 (0.3167, 0.5118) | 0.6250 (0.3240, 0.9260) |
| **Age Group: 80-99** | | **0.1626 (0.1500, 0.1751)** | 0.0568 (0.0408, 0.0729) | 0.2536 (0.2210, 0.2863) | 0.1508 (0.1181, 0.1835) | 0.1374 (0.1024, 0.1725) | 0.2974 (0.2479, 0.3469) | 0.0613 (0.0342, 0.0885) | 0.2655 (0.1969, 0.3341) | 0.1429 (0.0797, 0.2060) | 0.2429 (0.1579, 0.3278) | 0.2500 (0.0000, 0.5192) |
| **Gender: Male** | yes | **0.6609 (0.6448, 0.6769)** | 0.6519 (0.6188, 0.6849) | 0.6528 (0.6171, 0.6885) | 0.6585 (0.6151, 0.7018) | 0.6870 (0.6398, 0.7342) | 0.6810 (0.6306, 0.7315) | 0.6651 (0.6117, 0.7185) | 0.5664 (0.4893, 0.6434) | 0.7738 (0.6983, 0.8493) | 0.6571 (0.5632, 0.7511) | 0.5000 (0.1892, 0.8108) |
| **Race: Asian** | yes | **0.1034 (0.0931, 0.1137)** | 0.0941 (0.0739, 0.1144) | 0.0894 (0.0680, 0.1108) | 0.0585 (0.0370, 0.0799) | 0.1221 (0.0888, 0.1555) | 0.0905 (0.0595, 0.1216) | 0.2547 (0.2054, 0.3041) | 0.0619 (0.0245, 0.0994) | 0.0952 (0.0422, 0.1482) | 0.0857 (0.0303, 0.1411) | 0.0000 (0.0000, 0.3750) |
| **Race: Black Or African-American** | yes | **0.1566 (0.1443, 0.1689)** | 0.0835 (0.0643, 0.1027) | 0.1518 (0.1248, 0.1787) | 0.1815 (0.1463, 0.2168) | 0.1031 (0.0721, 0.1340) | 0.2241 (0.1790, 0.2693) | 0.2500 (0.2010, 0.2990) | 0.1770 (0.1177, 0.2363) | 0.3095 (0.2261, 0.3930) | 0.1286 (0.0623, 0.1949) | 0.2500 (0.0000, 0.5192) |
| **Race: Multiracial** | | **0.2757 (0.2606, 0.2909)** | 0.3535 (0.3203, 0.3866) | 0.1933 (0.1637, 0.2230) | 0.2954 (0.2537, 0.3371) | 0.3282 (0.2804, 0.3761) | 0.1983 (0.1551, 0.2414) | 0.2972 (0.2454, 0.3489) | 0.2124 (0.1488, 0.2760) | 0.2976 (0.2151, 0.3802) | 0.2000 (0.1208, 0.2792) | 0.2500 (0.0000, 0.5192) |
| **Race: Native American** | | **0.0051 (0.0027, 0.0075)** | 0.0089 (0.0024, 0.0154) | 0.0062 (0.0003, 0.0121) | 0.0062 (0.0000, 0.0133) | 0.0000 (0.0000, 0.0115) | 0.0000 (0.0000, 0.0129) | 0.0047 (0.0000, 0.0125) | 0.0088 (0.0000, 0.0234) | 0.0000 (0.0000, 0.0357) | 0.0000 (0.0000, 0.0429) | 0.0000 (0.0000, 0.3750) |
| **Race: Unknown** | | **0.0494 (0.0420, 0.0567)** | 0.0373 (0.0242, 0.0504) | 0.0499 (0.0335, 0.0662) | 0.0615 (0.0396, 0.0835) | 0.0954 (0.0655, 0.1253) | 0.0302 (0.0117, 0.0487) | 0.0330 (0.0128, 0.0533) | 0.0442 (0.0123, 0.0762) | 0.0714 (0.0249, 0.1179) | 0.0143 (0.0000, 0.0378) | 0.0000 (0.0000, 0.3750) |

| Demographics | Feature Selected | all | C 1 | C 2 | C 3 | C 4 | C 5 | C 6 | C 7 | C 8 | C 9 | C10 |
|---|---|---|---|---|---|---|---|---|---|---|---|---|
| **Race: White** | | **0.4098 (0.3931, 0.4265)** | 0.4227 (0.3885, 0.4570) | 0.5094 (0.4718, 0.5469) | 0.3969 (0.3522, 0.4416) | 0.3511 (0.3025, 0.3997) | 0.4569 (0.4030, 0.5108) | 0.1604 (0.1188, 0.2019) | 0.4956 (0.4179, 0.5733) | 0.2262 (0.1507, 0.3017) | 0.5714 (0.4734, 0.6694) | 0.5000 (0.1892, 0.8108) |

#### Comorbidities

*Table S5c: Comorbidities - means and 90% confidence intervals for the whole training set and among each phenocluster. All values are the are the proportion of the set that diagnosis at admission.*

| Comorbidities | Feature Selected | all | C 1 | C 2 | C 3 | C 4 | C 5 | C 6 | C 7 | C 8 | C 9 | C10 |
|---|---|---|---|---|---|---|---|---|---|---|---|---|
| **Asthma** | | **0.0800 (0.0708, 0.0892)** | 0.1012 (0.0803, 0.1222) | 0.0852 (0.0643, 0.1062) | 0.0708 (0.0473, 0.0942) | 0.0802 (0.0525, 0.1078) | 0.0388 (0.0179, 0.0597) | 0.0943 (0.0612, 0.1274) | 0.0619 (0.0245, 0.0994) | 0.0595 (0.0168, 0.1022) | 0.0714 (0.0204, 0.1224) | 0.0000 (0.0000, 0.3750) |
| **Atrial Fibrillation** | yes | **0.1991 (0.1856, 0.2127)** | 0.1581 (0.1328, 0.1834) | 0.2599 (0.2269, 0.2928) | 0.1969 (0.1606, 0.2333) | 0.1641 (0.1264, 0.2018) | 0.2759 (0.2275, 0.3242) | 0.0849 (0.0533, 0.1165) | 0.3363 (0.2629, 0.4097) | 0.1190 (0.0606, 0.1775) | 0.2286 (0.1454, 0.3117) | 0.1250 (0.0000, 0.3306) |
| **Autoimmune Disorder** | yes | **0.0340 (0.0279, 0.0402)** | 0.0302 (0.0183, 0.0421) | 0.0353 (0.0215, 0.0492) | 0.0431 (0.0245, 0.0616) | 0.0267 (0.0103, 0.0431) | 0.0302 (0.0117, 0.0487) | 0.0377 (0.0162, 0.0593) | 0.0442 (0.0123, 0.0762) | 0.0000 (0.0000, 0.0357) | 0.0714 (0.0204, 0.1224) | 0.0000 (0.0000, 0.3750) |
| **Cancer** | | **0.1298 (0.1184, 0.1412)** | 0.0995 (0.0787, 0.1202) | 0.1206 (0.0961, 0.1450) | 0.1292 (0.0986, 0.1599) | 0.1260 (0.0922, 0.1597) | 0.1681 (0.1276, 0.2086) | 0.0802 (0.0494, 0.1109) | 0.2566 (0.1888, 0.3245) | 0.0714 (0.0249, 0.1179) | 0.3000 (0.2093, 0.3907) | 0.5000 (0.1892, 0.8108) |
| **Cardiac Defibrillator** | | **0.0136 (0.0097, 0.0176)** | 0.0053 (0.0003, 0.0104) | 0.0146 (0.0056, 0.0235) | 0.0185 (0.0062, 0.0308) | 0.0115 (0.0006, 0.0223) | 0.0388 (0.0179, 0.0597) | 0.0000 (0.0000, 0.0142) | 0.0177 (0.0000, 0.0382) | 0.0119 (0.0000, 0.0315) | 0.0143 (0.0000, 0.0378) | 0.0000 (0.0000, 0.3750) |
| **Chronic Kidney Disease** | | **0.1489 (0.1369, 0.1610)** | 0.0231 (0.0127, 0.0335) | 0.2121 (0.1814, 0.2427) | 0.0985 (0.0712, 0.1257) | 0.0840 (0.0557, 0.1122) | 0.4914 (0.4373, 0.5455) | 0.0519 (0.0268, 0.0770) | 0.2301 (0.1647, 0.2955) | 0.1190 (0.0606, 0.1775) | 0.2857 (0.1963, 0.3752) | 0.0000 (0.0000, 0.3750) |
| **Chronic Liver Disorder** | | **0.0162 (0.0119, 0.0205)** | 0.0107 (0.0035, 0.0178) | 0.0125 (0.0041, 0.0208) | 0.0185 (0.0062, 0.0308) | 0.0076 (0.0000, 0.0165) | 0.0172 (0.0032, 0.0313) | 0.0047 (0.0000, 0.0125) | 0.0708 (0.0309, 0.1107) | 0.0000 (0.0000, 0.0357) | 0.0714 (0.0204, 0.1224) | 0.0000 (0.0000, 0.3750) |
| **Chronic Obstructive Pulmonary Disease** | | **0.0885 (0.0789, 0.0982)** | 0.0657 (0.0485, 0.0829) | 0.1268 (0.1018, 0.1518) | 0.0954 (0.0685, 0.1222) | 0.0725 (0.0461, 0.0989) | 0.1034 (0.0705, 0.1364) | 0.0519 (0.0268, 0.0770) | 0.1327 (0.0800, 0.1855) | 0.0238 (0.0000, 0.0513) | 0.0857 (0.0303, 0.1411) | 0.2500 (0.0000, 0.5192) |

| Comorbidities | Feature Selected | all | C 1 | C 2 | C 3 | C 4 | C 5 | C 6 | C 7 | C 8 | C 9 | C10 |
|---|---|---|---|---|---|---|---|---|---|---|---|---|
| **Congestive Heart Failure** | | **0.1004 (0.0902, 0.1106)** | 0.0302 (0.0183, 0.0421) | 0.1393 (0.1133, 0.1653) | 0.0769 (0.0526, 0.1013) | 0.0687 (0.0429, 0.0945) | 0.2716 (0.2234, 0.3197) | 0.0189 (0.0035, 0.0343) | 0.1770 (0.1177, 0.2363) | 0.0714 (0.0249, 0.1179) | 0.2143 (0.1330, 0.2955) | 0.1250 (0.0000, 0.3306) |
| **Coronary Artery Disease** | | **0.1677 (0.1550, 0.1803)** | 0.1030 (0.0819, 0.1241) | 0.2287 (0.1972, 0.2602) | 0.1415 (0.1097, 0.1734) | 0.1183 (0.0854, 0.1512) | 0.3060 (0.2562, 0.3559) | 0.1274 (0.0896, 0.1651) | 0.1947 (0.1331, 0.2562) | 0.1310 (0.0700, 0.1919) | 0.2429 (0.1579, 0.3278) | 0.1250 (0.0000, 0.3306) |
| **Cystic Fibrosis 0** | yes | **1.0000 (0.9987, 1.0000)** | 1.0000 (0.9947, 1.0000) | 1.0000 (0.9938, 1.0000) | 1.0000 (0.9908, 1.0000) | 1.0000 (0.9885, 1.0000) | 1.0000 (0.9871, 1.0000) | 1.0000 (0.9858, 1.0000) | 1.0000 (0.9735, 1.0000) | 1.0000 (0.9643, 1.0000) | 1.0000 (0.9571, 1.0000) | 1.0000 (0.6250, 1.0000) |
| **Dementia or Neurological** | yes | **0.2600 (0.2451, 0.2749)** | 0.1936 (0.1662, 0.2210) | 0.2869 (0.2529, 0.3209) | 0.2677 (0.2272, 0.3082) | 0.2290 (0.1862, 0.2718) | 0.3405 (0.2892, 0.3918) | 0.1792 (0.1358, 0.2227) | 0.4425 (0.3653, 0.5197) | 0.2024 (0.1298, 0.2749) | 0.4571 (0.3585, 0.5558) | 0.1250 (0.0000, 0.3306) |
| **Diabetes Mellitus** | yes | **0.4570 (0.4401, 0.4739)** | 0.3393 (0.3064, 0.3721) | 0.5031 (0.4656, 0.5407) | 0.3600 (0.3161, 0.4039) | 0.4466 (0.3959, 0.4972) | 0.6250 (0.5726, 0.6774) | 0.5660 (0.5099, 0.6222) | 0.4248 (0.3480, 0.5016) | 0.6667 (0.5816, 0.7518) | 0.4571 (0.3585, 0.5558) | 0.7500 (0.4808, 1.0192) |
| **End Stage Renal Disorder** | | **0.0557 (0.0480, 0.0635)** | 0.0142 (0.0060, 0.0224) | 0.0353 (0.0215, 0.0492) | 0.0092 (0.0005, 0.0180) | 0.0267 (0.0103, 0.0431) | 0.2888 (0.2397, 0.3378) | 0.0189 (0.0035, 0.0343) | 0.0973 (0.0513, 0.1434) | 0.0714 (0.0249, 0.1179) | 0.1143 (0.0513, 0.1773) | 0.0000 (0.0000, 0.3750) |
| **HIV** | | **0.0085 (0.0054, 0.0116)** | 0.0089 (0.0024, 0.0154) | 0.0083 (0.0015, 0.0151) | 0.0092 (0.0005, 0.0180) | 0.0115 (0.0006, 0.0223) | 0.0043 (0.0000, 0.0114) | 0.0047 (0.0000, 0.0125) | 0.0177 (0.0000, 0.0382) | 0.0119 (0.0000, 0.0315) | 0.0000 (0.0000, 0.0429) | 0.0000 (0.0000, 0.3750) |
| **Hypertension** | | **0.6638 (0.6478, 0.6799)** | 0.5293 (0.4947, 0.5639) | 0.7963 (0.7660, 0.8265) | 0.6308 (0.5867, 0.6749) | 0.6374 (0.5885, 0.6864) | 0.7974 (0.7539, 0.8409) | 0.6887 (0.6362, 0.7411) | 0.6549 (0.5810, 0.7288) | 0.6190 (0.5314, 0.7067) | 0.6571 (0.5632, 0.7511) | 0.5000 (0.1892, 0.8108) |
| **Interstitial Lung Disease** | yes | **0.2987 (0.2832, 0.3143)** | 0.3481 (0.3151, 0.3812) | 0.3098 (0.2751, 0.3445) | 0.4092 (0.3643, 0.4542) | 0.3550 (0.3062, 0.4037) | 0.3276 (0.2768, 0.3784) | 0.0472 (0.0232, 0.0712) | 0.1947 (0.1331, 0.2562) | 0.1786 (0.1094, 0.2477) | 0.1143 (0.0513, 0.1773) | 0.0000 (0.0000, 0.3750) |
| **Pace Maker** | | **0.0217 (0.0168, 0.0266)** | 0.0089 (0.0024, 0.0154) | 0.0270 (0.0149, 0.0392) | 0.0185 (0.0062, 0.0308) | 0.0115 (0.0006, 0.0223) | 0.0474 (0.0244, 0.0704) | 0.0189 (0.0035, 0.0343) | 0.0354 (0.0067, 0.0641) | 0.0000 (0.0000, 0.0357) | 0.0714 (0.0204, 0.1224) | 0.0000 (0.0000, 0.3750) |
| **Pregnancy** | | **0.0009 (0.0000, 0.0018)** | 0.0018 (0.0000, 0.0047) | 0.0000 (0.0000, 0.0062) | 0.0000 (0.0000, 0.0092) | 0.0038 (0.0000, 0.0101) | 0.0000 (0.0000, 0.0129) | 0.0000 (0.0000, 0.0142) | 0.0000 (0.0000, 0.0265) | 0.0000 (0.0000, 0.0357) | 0.0000 (0.0000, 0.0429) | 0.0000 (0.0000, 0.3750) |

| Comorbidities | Feature Selected | all | C 1 | C 2 | C 3 | C 4 | C 5 | C 6 | C 7 | C 8 | C 9 | C10 |
|---|---|---|---|---|---|---|---|---|---|---|---|---|
| **Pulmonary Hypertension** | yes | **0.1000 (0.0898, 0.1102)** | 0.1083 (0.0868, 0.1299) | 0.0832 (0.0624, 0.1039) | 0.1262 (0.0958, 0.1565) | 0.1069 (0.0754, 0.1383) | 0.0991 (0.0668, 0.1315) | 0.0566 (0.0304, 0.0828) | 0.1327 (0.0800, 0.1855) | 0.0476 (0.0092, 0.0861) | 0.1286 (0.0623, 0.1949) | 0.2500 (0.0000, 0.5192) |
| **Sickle or Thalassemia** | | **0.0026 (0.0008, 0.0043)** | 0.0036 (0.0000, 0.0077) | 0.0000 (0.0000, 0.0062) | 0.0031 (0.0000, 0.0081) | 0.0000 (0.0000, 0.0115) | 0.0129 (0.0007, 0.0252) | 0.0000 (0.0000, 0.0142) | 0.0000 (0.0000, 0.0265) | 0.0000 (0.0000, 0.0357) | 0.0000 (0.0000, 0.0429) | 0.0000 (0.0000, 0.3750) |
| **Stroke** | | **0.1157 (0.1049, 0.1266)** | 0.0782 (0.0595, 0.0968) | 0.1247 (0.0999, 0.1495) | 0.1169 (0.0876, 0.1463) | 0.1183 (0.0854, 0.1512) | 0.1724 (0.1315, 0.2133) | 0.0943 (0.0612, 0.1274) | 0.1770 (0.1177, 0.2363) | 0.0714 (0.0249, 0.1179) | 0.1857 (0.1087, 0.2627) | 0.0000 (0.0000, 0.3750) |
| **Transplant** | | **0.0170 (0.0126, 0.0214)** | 0.0089 (0.0024, 0.0154) | 0.0229 (0.0116, 0.0341) | 0.0123 (0.0022, 0.0224) | 0.0153 (0.0028, 0.0278) | 0.0216 (0.0058, 0.0373) | 0.0094 (0.0000, 0.0204) | 0.0531 (0.0182, 0.0879) | 0.0238 (0.0000, 0.0513) | 0.0143 (0.0000, 0.0378) | 0.0000 (0.0000, 0.3750) |

#### Home Medications

*Table S5d: Home Medications - means and 90% confidence intervals for the whole training set and among each phenocluster. All values are the are the proportion of the set that were using that medication at admission.*

| Home Medications | Feature Selected | all | C 1 | C 2 | C 3 | C 4 | C 5 | C 6 | C 7 | C 8 | C 9 | C10 |
|---|---|---|---|---|---|---|---|---|---|---|---|---|
| **ACE Inhibitors** | yes | **0.0902 (0.0805, 0.0999)** | 0.0817 (0.0627, 0.1007) | 0.1289 (0.1037, 0.1541) | 0.0769 (0.0526, 0.1013) | 0.0496 (0.0275, 0.0717) | 0.1164 (0.0817, 0.1511) | 0.0849 (0.0533, 0.1165) | 0.0796 (0.0376, 0.1217) | 0.0833 (0.0334, 0.1332) | 0.0714 (0.0204, 0.1224) | 0.0000 (0.0000, 0.3750) |
| **Anticoagulants** | | **0.0698 (0.0611, 0.0784)** | 0.0409 (0.0271, 0.0546) | 0.0936 (0.0717, 0.1154) | 0.0923 (0.0659, 0.1188) | 0.0458 (0.0245, 0.0671) | 0.1164 (0.0817, 0.1511) | 0.0377 (0.0162, 0.0593) | 0.0708 (0.0309, 0.1107) | 0.0119 (0.0000, 0.0315) | 0.1286 (0.0623, 0.1949) | 0.1250 (0.0000, 0.3306) |
| **Beta Blockers** | yes | **0.2030 (0.1893, 0.2166)** | 0.0941 (0.0739, 0.1144) | 0.3139 (0.2791, 0.3488) | 0.1600 (0.1265, 0.1935) | 0.1450 (0.1092, 0.1809) | 0.3966 (0.3436, 0.4495) | 0.2028 (0.1573, 0.2484) | 0.1770 (0.1177, 0.2363) | 0.1310 (0.0700, 0.1919) | 0.2143 (0.1330, 0.2955) | 0.2500 (0.0000, 0.5192) |
| **Calcium Channel Blockers** | yes | **0.1460 (0.1340, 0.1579)** | 0.0853 (0.0659, 0.1046) | 0.2058 (0.1755, 0.2362) | 0.1446 (0.1125, 0.1768) | 0.0763 (0.0493, 0.1034) | 0.2543 (0.2072, 0.3014) | 0.1462 (0.1062, 0.1862) | 0.1593 (0.1024, 0.2162) | 0.1190 (0.0606, 0.1775) | 0.1571 (0.0851, 0.2292) | 0.0000 (0.0000, 0.3750) |

| Home Medications | Feature Selected | all | C 1 | C 2 | C 3 | C 4 | C 5 | C 6 | C 7 | C 8 | C 9 | C10 |
|---|---|---|---|---|---|---|---|---|---|---|---|---|
| **GLP-1 Receptor Agonists** | | **0.0157 (0.0115, 0.0200)** | 0.0053 (0.0003, 0.0104) | 0.0208 (0.0101, 0.0315) | 0.0154 (0.0041, 0.0266) | 0.0115 (0.0006, 0.0223) | 0.0302 (0.0117, 0.0487) | 0.0142 (0.0008, 0.0275) | 0.0177 (0.0000, 0.0382) | 0.0476 (0.0092, 0.0861) | 0.0000 (0.0000, 0.0429) | 0.0000 (0.0000, 0.3750) |
| **Immunomodulator Suppressants** | yes | **0.0400 (0.0333, 0.0467)** | 0.0355 (0.0227, 0.0484) | 0.0416 (0.0266, 0.0566) | 0.0431 (0.0245, 0.0616) | 0.0305 (0.0130, 0.0481) | 0.0388 (0.0179, 0.0597) | 0.0377 (0.0162, 0.0593) | 0.0708 (0.0309, 0.1107) | 0.0119 (0.0000, 0.0315) | 0.0857 (0.0303, 0.1411) | 0.0000 (0.0000, 0.3750) |
| **Inhaled Pulmonary Therapies** | yes | **0.1106 (0.1000, 0.1213)** | 0.0977 (0.0771, 0.1183) | 0.1351 (0.1095, 0.1608) | 0.1354 (0.1041, 0.1666) | 0.0725 (0.0461, 0.0989) | 0.1078 (0.0742, 0.1413) | 0.0943 (0.0612, 0.1274) | 0.1593 (0.1024, 0.2162) | 0.0357 (0.0022, 0.0692) | 0.1143 (0.0513, 0.1773) | 0.3750 (0.0740, 0.6760) |
| **Insulin** | | **0.0047 (0.0024, 0.0070)** | 0.0036 (0.0000, 0.0077) | 0.0021 (0.0000, 0.0055) | 0.0092 (0.0005, 0.0180) | 0.0038 (0.0000, 0.0101) | 0.0129 (0.0007, 0.0252) | 0.0047 (0.0000, 0.0125) | 0.0000 (0.0000, 0.0265) | 0.0000 (0.0000, 0.0357) | 0.0000 (0.0000, 0.0429) | 0.0000 (0.0000, 0.3750) |
| **NSAIDs** | yes | **0.2034 (0.1897, 0.2171)** | 0.1226 (0.0998, 0.1453) | 0.2807 (0.2469, 0.3144) | 0.1938 (0.1577, 0.2300) | 0.1336 (0.0989, 0.1682) | 0.3060 (0.2562, 0.3559) | 0.1981 (0.1530, 0.2432) | 0.2389 (0.1727, 0.3052) | 0.1548 (0.0895, 0.2201) | 0.2714 (0.1834, 0.3595) | 0.5000 (0.1892, 0.8108) |
| **Sodium glucose transporter inhibitors** | | **0.0153 (0.0112, 0.0195)** | 0.0124 (0.0047, 0.0201) | 0.0291 (0.0165, 0.0417) | 0.0123 (0.0022, 0.0224) | 0.0038 (0.0000, 0.0101) | 0.0129 (0.0007, 0.0252) | 0.0142 (0.0008, 0.0275) | 0.0000 (0.0000, 0.0265) | 0.0119 (0.0000, 0.0315) | 0.0429 (0.0028, 0.0830) | 0.0000 (0.0000, 0.3750) |

#### Other Indicators

*Table S5e: Other Indicators - means and 90% confidence intervals for the whole training set and among each phenocluster. All values are the are the proportion of the set that were using that medication at admission.*

| Other Indicators | Feature Selected | all | C 1 | C 2 | C 3 | C 4 | C 5 | C 6 | C 7 | C 8 | C 9 | C10 |
|---|---|---|---|---|---|---|---|---|---|---|---|---|
| **Steroid response Negative** | | **0.1881 (0.1748, 0.2013)** | 0.1528 (0.1278, 0.1777) | 0.1913 (0.1617, 0.2208) | 0.2000 (0.1634, 0.2366) | 0.2519 (0.2077, 0.2961) | 0.1336 (0.0968, 0.1704) | 0.2830 (0.2320, 0.3340) | 0.0531 (0.0182, 0.0879) | 0.3095 (0.2261, 0.3930) | 0.1286 (0.0623, 0.1949) | 0.1250 (0.0000, 0.3306) |
| **Steroid response None** | | **0.4532 (0.4363, 0.4701)** | 0.5098 (0.4751, 0.5445) | 0.3659 (0.3297, 0.4021) | 0.5077 (0.4620, 0.5534) | 0.5076 (0.4567, 0.5585) | 0.5000 (0.4459, 0.5541) | 0.3585 (0.3042, 0.4128) | 0.2655 (0.1969, 0.3341) | 0.5714 (0.4821, 0.6608) | 0.4429 (0.3445, 0.5412) | 0.3750 (0.0740, 0.6760) |

| Other Indicators | Feature Selected | all | C 1 | C 2 | C 3 | C 4 | C 5 | C 6 | C 7 | C 8 | C 9 | C10 |
|---|---|---|---|---|---|---|---|---|---|---|---|---|
| **Steroid response Positive** | | **0.1911 (0.1777, 0.2044)** | 0.1545 (0.1295, 0.1796) | 0.2516 (0.2190, 0.2841) | 0.1477 (0.1153, 0.1801) | 0.1031 (0.0721, 0.1340) | 0.1853 (0.1433, 0.2274) | 0.1509 (0.1104, 0.1915) | 0.5221 (0.4445, 0.5998) | 0.0357 (0.0022, 0.0692) | 0.3714 (0.2757, 0.4671) | 0.3750 (0.0740, 0.6760) |
| **Steroid Response: Neutral** | yes | **0.1677 (0.1550, 0.1803)** | 0.1829 (0.1561, 0.2098) | 0.1913 (0.1617, 0.2208) | 0.1446 (0.1125, 0.1768) | 0.1374 (0.1024, 0.1725) | 0.1810 (0.1394, 0.2227) | 0.2075 (0.1616, 0.2535) | 0.1593 (0.1024, 0.2162) | 0.0833 (0.0334, 0.1332) | 0.0571 (0.0112, 0.1031) | 0.1250 (0.0000, 0.3306) |
| **Smoking Status: Non-Smoker** | yes | **0.8532 (0.8412, 0.8652)** | 0.9432 (0.9271, 0.9592) | 0.8919 (0.8686, 0.9152) | 0.8646 (0.8334, 0.8959) | 0.7099 (0.6637, 0.7561) | 0.7802 (0.7354, 0.8250) | 0.8774 (0.8402, 0.9145) | 0.7788 (0.7142, 0.8433) | 0.6429 (0.5563, 0.7294) | 0.8857 (0.8227, 0.9487) | 0.8750 (0.6694, 1.0806) |
| **Smoking Status: Smoker** | | **0.0387 (0.0322, 0.0453)** | 0.0107 (0.0035, 0.0178) | 0.0374 (0.0232, 0.0517) | 0.0369 (0.0197, 0.0542) | 0.0802 (0.0525, 0.1078) | 0.0603 (0.0346, 0.0861) | 0.0142 (0.0008, 0.0275) | 0.0885 (0.0444, 0.1326) | 0.0238 (0.0000, 0.0513) | 0.0571 (0.0112, 0.1031) | 0.1250 (0.0000, 0.3306) |
| **Smoking Status: Unknown** | yes | **0.1081 (0.0975, 0.1186)** | 0.0462 (0.0316, 0.0607) | 0.0707 (0.0514, 0.0899) | 0.0985 (0.0712, 0.1257) | 0.2099 (0.1685, 0.2514) | 0.1595 (0.1199, 0.1991) | 0.1085 (0.0733, 0.1437) | 0.1327 (0.0800, 0.1855) | 0.3333 (0.2482, 0.4184) | 0.0571 (0.0112, 0.1031) | 0.0000 (0.0000, 0.3750) |
| **Ventilator Group: Conventional** | N/A | **0.6311 (0.6147, 0.6474)** | 0.6359 (0.6025, 0.6693) | 0.7110 (0.6770, 0.7451) | 0.5662 (0.5209, 0.6114) | 0.6107 (0.5610, 0.6603) | 0.6638 (0.6127, 0.7149) | 0.4906 (0.4340, 0.5472) | 0.8319 (0.7737, 0.8900) | 0.4048 (0.3161, 0.4934) | 0.7000 (0.6093, 0.7907) | 0.5000 (0.1892, 0.8108) |
| **Ventilator Group: Portable** | N/A | **0.3434 (0.3273, 0.3595)** | 0.3464 (0.3133, 0.3794) | 0.2599 (0.2269, 0.2928) | 0.4062 (0.3613, 0.4510) | 0.3626 (0.3136, 0.4115) | 0.3147 (0.2644, 0.3649) | 0.4811 (0.4246, 0.5377) | 0.1681 (0.1100, 0.2263) | 0.5238 (0.4336, 0.6140) | 0.2571 (0.1706, 0.3437) | 0.5000 (0.1892, 0.8108) |
| **Ventilator Group: Unknown** | N/A | **0.0187 (0.0141, 0.0233)** | 0.0089 (0.0024, 0.0154) | 0.0208 (0.0101, 0.0315) | 0.0215 (0.0083, 0.0348) | 0.0267 (0.0103, 0.0431) | 0.0129 (0.0007, 0.0252) | 0.0189 (0.0035, 0.0343) | 0.0000 (0.0000, 0.0265) | 0.0714 (0.0249, 0.1179) | 0.0286 (0.0000, 0.0616) | 0.0000 (0.0000, 0.3750) |
| **Ventilator Group: None** | N/A | **0.0068 (0.0040, 0.0096)** | 0.0089 (0.0024, 0.0154) | 0.0083 (0.0015, 0.0151) | 0.0062 (0.0000, 0.0133) | 0.0000 (0.0000, 0.0115) | 0.0086 (0.0000, 0.0186) | 0.0094 (0.0000, 0.0204) | 0.0000 (0.0000, 0.0265) | 0.0000 (0.0000, 0.0357) | 0.0143 (0.0000, 0.0378) | 0.0000 (0.0000, 0.3750) |

#### Vitals

*Table S5f: Vitals - means and 90% confidence intervals for the whole training set and among each phenocluster. All values represent that feature's value.*

| Vitals | Feature Selected | all | C 1 | C 2 | C 3 | C 4 | C 5 | C 6 | C 7 | C 8 | C 9 | C10 |
|---|---|---|---|---|---|---|---|---|---|---|---|---|
| **Diastolic pressure: Median** | | **67.4821 (67.1710, 67.7933)** | 68.6066 (68.0443, 69.1688) | 68.9491 (68.2759, 69.6223) | 68.7138 (67.8636, 69.5641) | 65.1927 (64.3069, 66.0786) | 63.7996 (62.7144, 64.8847) | 70.6627 (69.6168, 71.7087) | 64.4115 (63.1936, 65.6294) | 63.8452 (62.0724, 65.6181) | 63.2786 (61.6734, 64.8838) | 65.9375 (60.9532, 70.9218) |
| **Diastolic pressure: Range** | | **41.8740 (41.1331, 42.6150)** | 34.7833 (33.7026, 35.8641) | 35.8108 (34.4981, 37.1236) | 32.9846 (31.4739, 34.4954) | 57.1794 (54.7512, 59.6076) | 46.4655 (44.1508, 48.7802) | 37.5236 (35.5268, 39.5204) | 63.0265 (59.2533, 66.7998) | 64.8810 (60.1838, 69.5781) | 60.2143 (55.5945, 64.8341) | 46.6250 (32.3702, 60.8798) |
| **Diastolic Pressure: IQR** | yes | **11.8826 (11.6838, 12.0813)** | 10.3512 (10.0263, 10.6762) | 11.0884 (10.6940, 11.4827) | 9.9777 (9.5745, 10.3809) | 14.3311 (13.6625, 14.9997) | 12.1422 (11.5727, 12.7118) | 12.5236 (11.8740, 13.1731) | 13.2721 (12.6596, 13.8846) | 20.8214 (18.9870, 22.6558) | 13.5607 (12.5989, 14.5225) | 11.9062 (8.3425, 15.4700) |
| **Diastolic pressure: Slope** | | **-0.2376 (-0.2662, -0.2090)** | -0.1737 (-0.2116, -0.1357) | -0.1256 (-0.1780, -0.0733) | -0.0187 (-0.0983, 0.0610) | -0.6897 (-0.7917, -0.5876) | -0.2065 (-0.2771, -0.1360) | -0.2748 (-0.3902, -0.1593) | -0.0324 (-0.0528, -0.0120) | -1.2252 (-1.5458, -0.9046) | -0.0654 (-0.1198, -0.0109) | 0.3861 (-0.0274, 0.7996) |
| **Diastolic Pressure: Fluctuations** | yes | **1.7260 (1.6292, 1.8228)** | 1.2530 (1.1690, 1.3371) | 1.2594 (1.1547, 1.3641) | 0.9568 (0.8379, 1.0756) | 1.1169 (1.0201, 1.2136) | 1.4384 (1.2776, 1.5992) | 1.2256 (1.0418, 1.4094) | 7.0360 (6.2014, 7.8707) | 0.9028 (0.7680, 1.0376) | 4.9118 (4.0993, 5.7242) | 2.4286 (0.0000, 4.8856) |
| **Heart rate: Median** | | **87.4711 (86.9411, 88.0011)** | 87.0329 (86.1311, 87.9347) | 84.3067 (83.2833, 85.3300) | 87.7631 (86.4003, 89.1259) | 95.0420 (93.0599, 97.0241) | 82.1767 (80.3563, 83.9971) | 88.2123 (86.5466, 89.8779) | 85.9646 (83.6807, 88.2485) | 98.7500 (95.6254, 101.875) | 86.3357 (83.2248, 89.4467) | 95.4375 (90.1452, 100.730) |
| **Heart rate: Range** | | **46.6515 (45.8404, 47.4626)** | 41.9147 (40.5085, 43.3210) | 40.2412 (38.7130, 41.7693) | 39.8277 (38.0779, 41.5775) | 57.4237 (55.1361, 59.7112) | 47.1767 (44.9773, 49.3761) | 42.2170 (39.4665, 44.9675) | 73.7699 (69.2322, 78.3076) | 61.0595 (56.5846, 65.5344) | 67.4429 (61.9540, 72.9318) | 75.8750 (46.3928, 105.357) |
| **Heart rate: IQR** | | **15.1861 (14.8533, 15.5188)** | 13.8495 (13.2973, 14.4017) | 13.2651 (12.6381, 13.8921) | 13.2792 (12.5629, 13.9956) | 19.4828 (18.1953, 20.7703) | 15.2446 (14.1329, 16.3563) | 15.0755 (13.8463, 16.3047) | 18.7854 (17.3686, 20.2022) | 21.7054 (19.2898, 24.1209) | 18.4393 (16.6094, 20.2692) | 14.9688 (10.5225, 19.4150) |
| **Heart rate: Slope** | | **-0.1687 (-0.1861, -0.1514)** | -0.1274 (-0.1519, -0.1029) | -0.0878 (-0.1184, -0.0572) | -0.2132 (-0.2623, -0.1640) | -0.3240 (-0.3921, -0.2559) | -0.1320 (-0.1933, -0.0707) | -0.2030 (-0.2596, -0.1465) | -0.0139 (-0.0303, 0.0025) | -0.5867 (-0.7695, -0.4039) | -0.0795 (-0.1532, -0.0058) | 0.1524 (-0.0527, 0.3574) |

| Vitals | Feature Selected | all | C 1 | C 2 | C 3 | C 4 | C 5 | C 6 | C 7 | C 8 | C 9 | C10 |
|---|---|---|---|---|---|---|---|---|---|---|---|---|
| **Heart Rate: Fluctuations** | yes | **1.3128 (1.2079, 1.4176)** | 0.9412 (0.8338, 1.0486) | 0.8421 (0.7133, 0.9709) | 0.7097 (0.5984, 0.8210) | 0.9919 (0.8543, 1.1295) | 1.2330 (1.0526, 1.4134) | 0.8333 (0.6493, 1.0174) | 4.0000 (3.3051, 4.6949) | 0.6429 (0.4585, 0.8273) | 2.0385 (1.6006, 2.4763) | 1.2500 (0.0164, 2.4836) |
| **Mean Arterial Pressure: Median** | | **86.1369 (85.7961, 86.4777)** | 86.0927 (85.4983, 86.6870) | 89.9785 (89.2294, 90.7276) | 86.2441 (85.3245, 87.1637) | 81.2564 (80.3357, 82.1771) | 84.5898 (83.3791, 85.8005) | 89.6454 (88.5242, 90.7666) | 83.1755 (81.8868, 84.4642) | 81.5575 (79.5341, 83.5809) | 82.7738 (81.1646, 84.3831) | 84.9792 (79.6318, 90.3266) |
| **Mean Arterial Pressure: Range** | | **45.8960 (45.0961, 46.6960)** | 37.7052 (36.4908, 38.9195) | 39.3056 (37.9433, 40.6679) | 35.4338 (33.7565, 37.1112) | 65.6069 (63.0726, 68.1411) | 52.4411 (50.0888, 54.7934) | 38.5802 (36.3474, 40.8130) | 67.6932 (64.3618, 71.0246) | 72.3651 (67.6300, 77.1001) | 65.0095 (60.2056, 69.8135) | 49.0833 (34.3306, 63.8361) |
| **Mean Arterial Pressure: IQR** | | **13.7874 (13.5482, 14.0266)** | 12.0620 (11.6672, 12.4569) | 12.9482 (12.4749, 13.4215) | 11.3759 (10.8848, 11.8670) | 17.0242 (16.1629, 17.8855) | 14.5920 (13.9685, 15.2154) | 13.1069 (12.3638, 13.8500) | 15.7891 (15.0973, 16.4809) | 23.9891 (21.7894, 26.1887) | 16.4214 (15.1730, 17.6699) | 13.8958 (9.5784, 18.2132) |
| **Mean Arterial Pressure: Slope** | yes | **-0.2015 (-0.2277, -0.1753)** | -0.1418 (-0.1717, -0.1120) | -0.0733 (-0.1190, -0.0276) | -0.0034 (-0.0726, 0.0658) | -0.6587 (-0.7654, -0.5519) | -0.1226 (-0.1927, -0.0526) | -0.1877 (-0.2692, -0.1062) | -0.0324 (-0.0554, -0.0095) | -1.3276 (-1.6250, -1.0302) | -0.0865 (-0.1748, 0.0017) | 0.4599 (-0.0307, 0.9504) |
| **Mean Arterial Pressure: Fluctuations** | | **1.8011 (1.6924, 1.9098)** | 1.3352 (1.2400, 1.4304) | 1.1655 (1.0719, 1.2592) | 0.9545 (0.8354, 1.0737) | 1.1379 (1.0432, 1.2326) | 1.5102 (1.3331, 1.6873) | 1.2456 (1.0605, 1.4307) | 7.0179 (6.0881, 7.9476) | 1.1364 (0.9178, 1.3550) | 5.3235 (4.4108, 6.2363) | 1.5000 (0.5744, 2.4256) |
| **Pulse Pressure: Median** | yes | **55.2074 (54.7670, 55.6479)** | 51.6679 (51.0174, 52.3183) | 62.3077 (61.3636, 63.2518) | 52.1600 (51.1871, 53.1329) | 47.6355 (46.5819, 48.6891) | 62.1336 (60.3210, 63.9462) | 55.8844 (54.5795, 57.1894) | 55.0708 (53.1740, 56.9676) | 51.9762 (49.3449, 54.6075) | 56.7500 (53.8137, 59.6863) | 52.7500 (47.9949, 57.5051) |
| **Pulse pressure: Range** | | **53.6004 (52.7807, 54.4201)** | 44.3393 (42.9288, 45.7497) | 50.5198 (48.9088, 52.1307) | 43.5446 (41.7080, 45.3812) | 64.7786 (62.5653, 66.9919) | 64.3319 (61.9402, 66.7236) | 45.9717 (43.6806, 48.2628) | 79.3982 (75.7232, 83.0733) | 72.0476 (67.5668, 76.5285) | 76.5714 (71.5840, 81.5589) | 64.8750 (52.7542, 76.9958) |
| **Pulse pressure: IQR** | | **16.4863 (16.2076, 16.7650)** | 13.7318 (13.2603, 14.2032) | 16.3051 (15.7490, 16.8612) | 14.5523 (13.8798, 15.2249) | 18.0067 (17.1490, 18.8644) | 19.8815 (18.9665, 20.7964) | 15.5330 (14.6175, 16.4486) | 20.1460 (19.0951, 21.1969) | 23.8899 (21.6240, 26.1557) | 19.4714 (17.8773, 21.0656) | 21.2500 (15.8789, 26.6211) |
| **Pulse pressure: Slope** | | **-0.0573 (-0.0866, -0.0280)** | -0.0204 (-0.0582, 0.0173) | -0.0093 (-0.0549, 0.0364) | 0.0381 (-0.0526, 0.1287) | -0.3956 (-0.5215, -0.2697) | 0.1252 (0.0378, 0.2125) | -0.0562 (-0.1563, 0.0438) | 0.0138 (-0.0139, 0.0416) | -0.5528 (-0.8886, -0.2171) | -0.0085 (-0.1038, 0.0868) | 0.0510 (-0.1501, 0.2521) |
| **Pulse pressure: Fluctuations** | | **1.7703 (1.6780, 1.8627)** | 1.4564 (1.3645, 1.5483) | 1.2145 (1.0984, 1.3306) | 0.9859 (0.8883, 1.0835) | 1.2346 (1.1340, 1.3351) | 1.4775 (1.2898, 1.6653) | 1.1783 (0.9838, 1.3728) | 6.6847 (5.9362, 7.4331) | 1.2740 (1.0534, 1.4946) | 4.7794 (3.9446, 5.6143) | 3.3333 (1.0201, 5.6465) |

| Vitals | Feature Selected | all | C 1 | C 2 | C 3 | C 4 | C 5 | C 6 | C 7 | C 8 | C 9 | C10 |
|---|---|---|---|---|---|---|---|---|---|---|---|---|
| **Respiration Rate: Median** | yes | **22.5900 (22.4408, 22.7392)** | 22.1012 (21.8175, 22.3850) | 20.9667 (20.7335, 21.2000) | 22.3877 (22.0474, 22.7280) | 26.5172 (26.0491, 26.9852) | 21.3103 (20.9073, 21.7134) | 23.8231 (23.3009, 24.3453) | 21.4469 (20.8386, 22.0552) | 27.3690 (26.4545, 28.2836) | 20.8214 (20.0221, 21.6207) | 20.0625 (18.5521, 21.5729) |
| **Respiration rate: Range** | | **17.9987 (17.4851, 18.5124)** | 17.9591 (16.8834, 19.0349) | 12.4158 (11.7198, 13.1118) | 15.4400 (14.4240, 16.4560) | 23.0954 (21.6382, 24.5526) | 17.6078 (16.1206, 19.0949) | 17.2453 (15.6328, 18.8578) | 30.0708 (26.2387, 33.9029) | 23.2738 (20.9341, 25.6135) | 27.5286 (22.3187, 32.7384) | 15.5000 (11.1039, 19.8961) |
| **Respiration rate: IQR** | | **4.6521 (4.5182, 4.7861)** | 4.6634 (4.3650, 4.9618) | 3.6169 (3.3802, 3.8537) | 4.6354 (4.2877, 4.9831) | 5.6536 (5.2276, 6.0796) | 4.5237 (4.1258, 4.9217) | 4.9493 (4.4966, 5.4020) | 5.9181 (5.3029, 6.5334) | 6.1220 (5.2556, 6.9885) | 3.8107 (3.2265, 4.3949) | 3.8750 (2.2246, 5.5254) |
| **Respiration rate: Slope** | | **-0.0908 (-0.1149, -0.0666)** | -0.0056 (-0.0445, 0.0334) | -0.0808 (-0.1240, -0.0375) | -0.1796 (-0.2594, -0.0997) | -0.1580 (-0.2601, -0.0560) | -0.0613 (-0.1266, 0.0040) | -0.2607 (-0.3483, -0.1730) | 0.0543 (0.0335, 0.0750) | -0.0766 (-0.2728, 0.1197) | -0.0578 (-0.1414, 0.0257) | 0.2557 (-0.0421, 0.5535) |
| **Respiration Rate: Fluctuations** | yes | **1.4080 (1.2985, 1.5174)** | 1.1875 (1.0741, 1.3009) | 1.0197 (0.9013, 1.1381) | 0.7519 (0.6431, 0.8607) | 1.0556 (0.9447, 1.1664) | 1.1750 (0.9969, 1.3531) | 0.9011 (0.7556, 1.0466) | 4.8636 (3.8756, 5.8517) | 0.8372 (0.6481, 1.0263) | 2.4583 (1.8586, 3.0581) | 1.5000 (0.6776, 2.3224) |
| **SpO2: Median** | | **95.2604 (95.1708, 95.3500)** | 94.7789 (94.6205, 94.9372) | 94.6746 (94.4804, 94.8689) | 94.7123 (94.4553, 94.9693) | 96.0992 (95.8178, 96.3807) | 96.1315 (95.8554, 96.4075) | 95.1816 (94.9124, 95.4508) | 96.4469 (96.1344, 96.7594) | 96.1845 (95.6304, 96.7387) | 96.6429 (96.1073, 97.1784) | 97.4375 (96.3120, 98.5630) |
| **SpO2: Range** | | **17.9132 (17.5667, 18.2597)** | 16.2664 (15.7270, 16.8058) | 15.6320 (14.9877, 16.2764) | 17.6031 (16.6128, 18.5933) | 24.4466 (23.2197, 25.6734) | 17.4526 (16.3400, 18.5652) | 17.1462 (16.0235, 18.2689) | 19.1858 (17.8828, 20.4889) | 23.3690 (20.8497, 25.8884) | 18.8143 (16.5762, 21.0524) | 20.1250 (9.1726, 31.0774) |
| **SpO2: IQR** | | **3.9820 (3.8949, 4.0692)** | 3.8237 (3.7061, 3.9413) | 3.7464 (3.5916, 3.9012) | 4.0100 (3.7306, 4.2894) | 4.8206 (4.4499, 5.1913) | 3.7457 (3.4663, 4.0250) | 3.8644 (3.6046, 4.1242) | 3.8451 (3.6147, 4.0756) | 4.8839 (4.0983, 5.6696) | 3.9357 (3.4186, 4.4529) | 3.5312 (2.4012, 4.6613) |
| **SpO2: Slope** | yes | **0.2337 (0.2085, 0.2589)** | 0.0863 (0.0503, 0.1223) | 0.0225 (-0.0252, 0.0701) | 0.4622 (0.3913, 0.5330) | 0.8669 (0.7741, 0.9597) | 0.0587 (-0.0122, 0.1297) | 0.1703 (0.0751, 0.2655) | -0.0046 (-0.0225, 0.0133) | 0.7493 (0.5780, 0.9205) | 0.0312 (-0.0486, 0.1111) | -0.0939 (-0.3662, 0.1785) |
| **SpO2: Fluctuations** | yes | **1.5296 (1.4564, 1.6027)** | 1.4224 (1.3260, 1.5187) | 1.3041 (1.1968, 1.4114) | 0.8385 (0.7384, 0.9387) | 1.0055 (0.8989, 1.1122) | 1.4251 (1.2573, 1.5930) | 1.1203 (0.9642, 1.2764) | 5.1111 (4.5133, 5.7090) | 0.6545 (0.4880, 0.8211) | 2.7627 (2.2735, 3.2519) | 2.6667 (0.6006, 4.7328) |
| **StoF: Median** | | **1.6886 (1.6615, 1.7157)** | 1.7623 (1.7127, 1.8119) | 1.7319 (1.6739, 1.7899) | 1.3436 (1.2919, 1.3952) | 1.3019 (1.2483, 1.3556) | 1.7550 (1.6686, 1.8415) | 1.6115 (1.5319, 1.6911) | 2.6232 (2.4863, 2.7600) | 1.2486 (1.1621, 1.3351) | 2.7690 (2.5445, 2.9936) | 2.6478 (1.9421, 3.3535) |

| Vitals | Feature Selected | all | C 1 | C 2 | C 3 | C 4 | C 5 | C 6 | C 7 | C 8 | C 9 | C10 |
|---|---|---|---|---|---|---|---|---|---|---|---|---|
| **StoF: Range** | | **2.3751 (2.3337, 2.4165)** | 2.8217 (2.7571, 2.8864) | 2.5628 (2.4818, 2.6438) | 1.5422 (1.4294, 1.6550) | 1.9016 (1.7791, 2.0240) | 2.2870 (2.1495, 2.4244) | 2.2352 (2.0993, 2.3711) | 3.4351 (3.3365, 3.5336) | 1.5355 (1.3192, 1.7518) | 3.1106 (2.9110, 3.3101) | 2.6783 (1.7653, 3.5913) |
| **StoF: IQR** | | **0.7791 (0.7526, 0.8056)** | 0.9743 (0.9180, 1.0306) | 0.9098 (0.8508, 0.9688) | 0.3845 (0.3383, 0.4307) | 0.4004 (0.3572, 0.4435) | 0.7791 (0.6907, 0.8675) | 0.6851 (0.6063, 0.7639) | 1.3776 (1.2324, 1.5228) | 0.5272 (0.3978, 0.6566) | 1.1410 (0.9734, 1.3087) | 1.1374 (0.5418, 1.7330) |
| **StoF: Slope** | | **-0.2518 (-0.2728, -0.2308)** | -0.3487 (-0.3771, -0.3202) | -0.3088 (-0.3531, -0.2645) | -0.2370 (-0.3022, -0.1719) | -0.2155 (-0.2970, -0.1341) | -0.1723 (-0.2414, -0.1033) | -0.2408 (-0.3103, -0.1713) | -0.1050 (-0.1246, -0.0853) | -0.0295 (-0.2196, 0.1605) | -0.0886 (-0.1985, 0.0213) | -0.1365 (-0.2227, -0.0502) |
| **StoF: Fluctuations** | yes | **1.0603 (0.9934, 1.1272)** | 0.9435 (0.8525, 1.0345) | 0.7868 (0.6861, 0.8875) | 0.8000 (0.6715, 0.9285) | 0.9196 (0.8333, 1.0060) | 1.0649 (0.8816, 1.2482) | 0.8868 (0.7006, 1.0730) | 2.1047 (1.7681, 2.4412) | 0.6296 (0.4508, 0.8084) | 1.3913 (0.8563, 1.9263) | 2.0000 (0.7259, 3.2741) |
| **Systolic pressure: Median** | | **123.0302 (122.497, 123.564)** | 120.6137 (119.772, 121.456) | 131.3690 (130.197, 132.542) | 120.9585 (119.635, 122.282) | 113.2443 (111.945, 114.543) | 126.3017 (124.244, 128.360) | 127.0660 (125.345, 128.788) | 120.1593 (118.057, 122.262) | 116.5357 (113.297, 119.775) | 120.9214 (118.084, 123.759) | 121.7500 (113.991, 129.509) |
| **Systolic Pressure: Range** | yes | **68.9413 (67.7615, 70.1210)** | 56.0906 (54.2327, 57.9485) | 60.9064 (58.8383, 62.9745) | 53.7569 (51.1935, 56.3203) | 95.1985 (91.7446, 98.6523) | 81.3190 (78.0457, 84.5923) | 55.4387 (52.1818, 58.6955) | 102.7080 (97.6087, 107.807) | 104.2738 (97.7533, 110.794) | 101.6286 (93.3297, 109.927) | 78.2500 (57.2735, 99.2265) |
| **Systolic pressure: IQR** | | **21.3428 (20.9604, 21.7251)** | 18.1639 (17.5297, 18.7980) | 20.2822 (19.5270, 21.0375) | 17.6162 (16.8037, 18.4286) | 26.3416 (25.0302, 27.6530) | 24.5603 (23.5454, 25.5753) | 19.0035 (17.7926, 20.2145) | 25.4845 (24.1563, 26.8128) | 35.6250 (32.1952, 39.0548) | 24.9929 (22.8448, 27.1409) | 24.7812 (17.7093, 31.8532) |
| **Systolic pressure: Slope** | | **-0.2207 (-0.2513, -0.1901)** | -0.1495 (-0.1883, -0.1107) | -0.0945 (-0.1520, -0.0370) | -0.0130 (-0.0966, 0.0707) | -0.7247 (-0.8415, -0.6079) | -0.0348 (-0.1141, 0.0445) | -0.2975 (-0.3868, -0.2082) | -0.0156 (-0.0417, 0.0104) | -1.4369 (-1.8015, -1.0722) | -0.0753 (-0.1427, -0.0078) | 0.3452 (0.0432, 0.6473) |
| **Systolic pressure: Fluctuations** | | **1.9019 (1.7974, 2.0064)** | 1.3723 (1.2743, 1.4703) | 1.4277 (1.2980, 1.5574) | 1.0196 (0.9036, 1.1356) | 1.1790 (1.0778, 1.2802) | 1.7638 (1.5575, 1.9701) | 1.3385 (1.1332, 1.5437) | 7.4821 (6.5875, 8.3768) | 1.1216 (0.9604, 1.2829) | 5.6029 (4.7135, 6.4924) | 2.8571 (0.8131, 4.9011) |
| **Temperature C: Median** | | **37.1075 (37.0856, 37.1294)** | 37.2909 (37.2515, 37.3302) | 37.0859 (37.0421, 37.1296) | 37.0223 (36.9590, 37.0856) | 37.2328 (37.1620, 37.3037) | 36.8879 (36.8102, 36.9657) | 37.1005 (37.0312, 37.1698) | 36.8642 (36.7958, 36.9325) | 37.1423 (36.9867, 37.2979) | 36.8407 (36.7306, 36.9509) | 36.8188 (36.6851, 36.9524) |
| **Temperature C: Range** | | **2.2984 (2.2609, 2.3359)** | 2.3675 (2.3021, 2.4328) | 2.1963 (2.1206, 2.2719) | 1.9237 (1.8310, 2.0164) | 2.3019 (2.1878, 2.4160) | 2.3466 (2.2114, 2.4817) | 2.1340 (2.0131, 2.2548) | 3.1062 (2.9320, 3.2803) | 2.2774 (2.0215, 2.5333) | 3.1129 (2.8783, 3.3474) | 3.3250 (2.4014, 4.2486) |

| Vitals | Feature Selected | all | C 1 | C 2 | C 3 | C 4 | C 5 | C 6 | C 7 | C 8 | C 9 | C10 |
|---|---|---|---|---|---|---|---|---|---|---|---|---|
| **Temperature C: IQR** | | **0.8085 (0.7916, 0.8255)** | 0.8370 (0.8059, 0.8681) | 0.7608 (0.7264, 0.7951) | 0.7155 (0.6721, 0.7589) | 0.9227 (0.8650, 0.9804) | 0.8279 (0.7689, 0.8869) | 0.8132 (0.7477, 0.8787) | 0.7509 (0.6976, 0.8042) | 0.8994 (0.7813, 1.0175) | 0.8396 (0.7449, 0.9344) | 0.6125 (0.4283, 0.7967) |
| **Temperature C: Slope** | | **-0.1420 (-0.1646, -0.1194)** | -0.1356 (-0.1696, -0.1017) | -0.1400 (-0.1804, -0.0995) | -0.2066 (-0.2714, -0.1418) | -0.0516 (-0.1538, 0.0506) | -0.1463 (-0.2334, -0.0592) | -0.2506 (-0.3138, -0.1874) | -0.0238 (-0.0383, -0.0092) | -0.2312 (-0.4344, -0.0281) | -0.0093 (-0.1076, 0.0889) | -0.0045 (-0.0639, 0.0548) |
| **Temperature C: Fluctuations** | | **1.3582 (1.2837, 1.4327)** | 1.1544 (1.0535, 1.2552) | 1.2216 (1.0864, 1.3568) | 0.9367 (0.7631, 1.1104) | 0.9213 (0.8003, 1.0424) | 1.0000 (0.8503, 1.1497) | 1.1176 (0.9241, 1.3112) | 2.9787 (2.5783, 3.3791) | 0.7500 (0.5025, 0.9975) | 2.7273 (2.2569, 3.1977) | 0.2500 (0.0000, 0.6612) |

#### Interventions

*Table S5g: Interventions - means and 90% confidence intervals for the whole training set and among each phenocluster. All values represent that feature's value.*

| Interventions | Feature Selected | all | C 1 | C 2 | C 3 | C 4 | C 5 | C 6 | C 7 | C 8 | C 9 | C10 |
|---|---|---|---|---|---|---|---|---|---|---|---|---|
| **Dynamic Compliance: Median** | yes | **25.2983 (24.5671, 26.0295)** | 27.0666 (24.7770, 29.3562) | 24.7489 (22.6433, 26.8544) | 27.6576 (23.9506, 31.3645) | 25.6539 (24.2465, 27.0614) | 24.8504 (23.4270, 26.2739) | 23.0389 (21.4746, 24.6032) | 23.2325 (21.8843, 24.5808) | 24.4175 (22.1075, 26.7275) | 23.1624 (21.1183, 25.2066) | 20.0152 (13.7841, 26.2464) |
| **Dynamic compliance: Range** | | **7.6844 (6.9047, 8.4641)** | 6.7395 (5.0444, 8.4345) | 6.1798 (3.8224, 8.5373) | 5.9127 (4.5568, 7.2687) | 9.8545 (7.5693, 12.1396) | 5.7390 (4.2220, 7.2559) | 7.4599 (5.6326, 9.2872) | 9.2734 (7.3430, 11.2037) | 9.8730 (6.1190, 13.6269) | 7.0466 (4.8134, 9.2797) | 4.4634 (1.6939, 7.2329) |
| **Dynamic compliance: Slope** | | **-2.0344 (-4.3939, 0.3252)** | -3.0040 (-10.325, 4.3168) | 2.8984 (-2.3264, 8.1231) | 1.0402 (-7.4095, 9.4899) | -5.4390 (-11.019, 0.1413) | -1.2814 (-6.6487, 4.0859) | -1.8410 (-7.5916, 3.9095) | 7.2309 (1.3046, 13.1572) | -11.1819 (-20.841, -1.5232) | 0.8010 (-1.9233, 3.5252) | -2.2795 (-9.0254, 4.4665) |
| **Static compliance: Median** | | **32.6773 (31.8065, 33.5481)** | 36.7148 (34.0104, 39.4193) | 31.3219 (28.9691, 33.6747) | 34.5254 (31.5238, 37.5271) | 31.4604 (29.5933, 33.3275) | 32.6856 (30.6013, 34.7699) | 30.1778 (27.2344, 33.1211) | 32.4552 (30.0041, 34.9064) | 31.4700 (28.3744, 34.5656) | 29.0714 (26.5279, 31.6150) | 28.8000 (18.4570, 39.1430) |
| **Static compliance: Range** | | **12.5234 (11.3038, 13.7430)** | 12.1953 (9.4402, 14.9504) | 8.9589 (6.0604, 11.8574) | 10.8475 (6.9709, 14.7241) | 15.3957 (12.3172, 18.4741) | 11.8144 (8.9670, 14.6619) | 4.3111 (1.8816, 6.7406) | 23.6716 (17.4768, 29.8665) | 9.9200 (6.4443, 13.3957) | 9.5714 (7.0934, 12.0494) | 2.0000 (-0.0806, 4.0806) |

| Interventions | Feature Selected | all | C 1 | C 2 | C 3 | C 4 | C 5 | C 6 | C 7 | C 8 | C 9 | C10 |
|---|---|---|---|---|---|---|---|---|---|---|---|---|
| **Static compliance: Slope** | | **2.4469 (-1.3309, 6.2246)** | 3.6236 (-6.8883, 14.1355) | 14.3521 (5.2793, 23.4250) | 2.4386 (-9.6352, 14.5125) | 0.3104 (-7.0009, 7.6218) | -6.5330 (-19.736, 6.6697) | 14.2428 (-5.6735, 34.1591) | 2.0108 (-9.5798, 13.6014) | 5.5127 (-3.1270, 14.1524) | -1.4647 (-14.842, 11.9128) | -17.5163 (-43.984, 8.9513) |
| **FiO2: Median** | | **67.8850 (67.0773, 68.6926)** | 63.4773 (61.9950, 64.9595) | 65.2817 (63.5381, 67.0253) | 78.2385 (76.5310, 79.9459) | 82.2882 (80.2122, 84.3641) | 65.8147 (63.2027, 68.4266) | 69.7005 (67.1790, 72.2219) | 42.1504 (39.5465, 44.7544) | 84.9286 (81.6511, 88.2060) | 43.8786 (39.1951, 48.5621) | 48.8125 (32.1263, 65.4987) |
| **FiO2: Range** | yes | **58.5392 (57.8073, 59.2711)** | 64.7016 (63.6630, 65.7402) | 61.6403 (60.2976, 62.9831) | 42.6523 (40.2647, 45.0399) | 57.6527 (55.2715, 60.0338) | 56.4698 (53.9509, 58.9888) | 56.0005 (53.6486, 58.3524) | 72.8230 (71.3517, 74.2943) | 49.9643 (45.4309, 54.4976) | 66.7143 (63.6822, 69.7463) | 56.8750 (38.9548, 74.7952) |
| **FiO2: IQR** | | **24.6248 (23.9222, 25.3274)** | 30.4352 (28.9508, 31.9196) | 28.4912 (26.8909, 30.0914) | 15.6323 (14.0391, 17.2255) | 21.0315 (19.1339, 22.9290) | 24.6929 (22.4022, 26.9836) | 21.6608 (19.5708, 23.7509) | 24.5221 (21.4052, 27.6391) | 22.4137 (18.7672, 26.0602) | 18.4750 (14.9783, 21.9717) | 21.3125 (10.0628, 32.5622) |
| **FiO2: Slope** | | **0.5434 (0.5010, 0.5857)** | 0.8871 (0.8222, 0.9520) | 0.8135 (0.7261, 0.9010) | 0.5009 (0.3725, 0.6293) | 0.0399 (-0.1110, 0.1908) | 0.2889 (0.1548, 0.4229) | 0.6743 (0.5262, 0.8224) | 0.1734 (0.1414, 0.2054) | -0.3263 (-0.6502, -0.0025) | 0.1019 (-0.0449, 0.2487) | 0.1955 (0.0076, 0.3834) |
| **FiO2: Fluctuations** | yes | **1.0897 (1.0331, 1.1463)** | 0.9844 (0.8995, 1.0693) | 0.9709 (0.8681, 1.0737) | 0.6250 (0.5121, 0.7379) | 0.8333 (0.7232, 0.9434) | 1.0316 (0.8712, 1.1920) | 1.0000 (0.7960, 1.2040) | 2.0500 (1.7872, 2.3128) | 0.7143 (0.4950, 0.9335) | 1.7115 (1.3381, 2.0849) | 1.0000 (0.4799, 1.5201) |

#### Labs

*Table S5h: Labs - means and 90% confidence intervals for the whole training set and among each phenocluster. All values represent that feature's value.*

| Labs | Feature Selected | all | C 1 | C 2 | C 3 | C 4 | C 5 | C 6 | C 7 | C 8 | C 9 | C10 |
|---|---|---|---|---|---|---|---|---|---|---|---|---|
| **Activated Partial Thromboplastin Time: Median** | | **37.0999 (36.3302, 37.8696)** | 33.9224 (32.8655, 34.9792) | 34.9763 (33.3491, 36.6036) | 36.2438 (34.3593, 38.1283) | 36.9587 (35.0547, 38.8627) | 42.9211 (39.3774, 46.4648) | 37.9205 (34.9575, 40.8835) | 40.7308 (37.7566, 43.7050) | 41.9455 (36.6317, 47.2592) | 38.0228 (35.3719, 40.6737) | 42.4100 (29.9594, 54.8606) |
| **Activated Partial Thromboplastin Time: Range** | | **11.6446 (10.2779, 13.0113)** | 2.2994 (1.0972, 3.5017) | 5.6981 (3.4772, 7.9190) | 8.0060 (5.0326, 10.9793) | 12.9265 (8.9344, 16.9186) | 18.3561 (12.9512, 23.7611) | 10.2335 (6.0967, 14.3704) | 39.6242 (30.5435, 48.7050) | 23.1403 (14.1373, 32.1432) | 25.3926 (16.2702, 34.5151) | 41.3400 (-15.8277, 98.5077) |
| **Alanine Aminotransferase (ALT/SGPT): Median** | | **64.3107 (58.8779, 69.7435)** | 50.5453 (47.7519, 53.3387) | 47.5136 (40.2000, 54.8272) | 58.0699 (51.8315, 64.3083) | 97.6489 (75.2050, 120.0927) | 68.5260 (51.1881, 85.8638) | 46.3538 (42.3103, 50.3973) | 73.4425 (25.1462, 121.7387) | 195.0774 (107.0770, 283.0778) | 55.3714 (36.8283, 73.9146) | 126.3750 (-24.2299, 276.9799) |

| Labs | Feature Selected | all | C 1 | C 2 | C 3 | C 4 | C 5 | C 6 | C 7 | C 8 | C 9 | C10 |
|---|---|---|---|---|---|---|---|---|---|---|---|---|
| **Alanine Aminotransferase (ALT/SGPT): Range** | | **58.9052 (46.5261, 71.2844)** | 18.0409 (15.5519, 20.5298) | 19.8075 (11.5689, 28.0462) | 12.0932 (9.1605, 15.0258) | 86.0687 (52.1987, 119.9387) | 44.5022 (22.6457, 66.3587) | 24.1651 (11.5940, 36.7362) | 244.0265 (142.3331, 345.7200) | 329.1310 (146.5633, 511.6986) | 295.1286 (44.8442, 545.4129) | 82.7500 (-3.1936, 168.6936) |
| **Alanine Aminotransferase (ALT/SGPT): Slope** | | **32.0223 (18.6810, 45.3635)** | -1.8229 (-3.2999, -0.3458) | 10.1908 (-2.6465, 23.0281) | -1.3819 (-8.0595, 5.2957) | 78.2607 (33.6404, 122.8810) | 24.6852 (7.5979, 41.7725) | 6.0404 (-5.1914, 17.2723) | 15.3788 (-0.3764, 31.1341) | 440.6230 (134.7789, 746.4670) | 31.3214 (5.9948, 56.6481) | 82.6540 (-55.7954, 221.1034) |
| **Albumin, Serum: Slope** | | **-0.3918 (-0.4152, -0.3685)** | -0.3385 (-0.3734, -0.3037) | -0.3221 (-0.3552, -0.2889) | -0.3579 (-0.4127, -0.3030) | -0.6290 (-0.7506, -0.5074) | -0.4075 (-0.4929, -0.3221) | -0.4166 (-0.4751, -0.3581) | -0.1181 (-0.1429, -0.0933) | -0.9378 (-1.1244, -0.7512) | -0.2736 (-0.3809, -0.1664) | -0.3001 (-0.5517, -0.0485) |
| **Albumin: Median** | yes | **3.0424 (3.0236, 3.0611)** | 3.2330 (3.2034, 3.2626) | 3.2931 (3.2600, 3.3262) | 2.7516 (2.7084, 2.7947) | 2.7011 (2.6474, 2.7549) | 2.7617 (2.6969, 2.8265) | 3.3906 (3.3446, 3.4366) | 2.8301 (2.7411, 2.9191) | 2.9482 (2.8511, 3.0453) | 2.7906 (2.6603, 2.9209) | 2.5563 (2.2282, 2.8843) |
| **Albumin: Range** | yes | **0.5110 (0.4960, 0.5260)** | 0.5131 (0.4898, 0.5365) | 0.4188 (0.3947, 0.4429) | 0.2382 (0.2174, 0.2590) | 0.4977 (0.4587, 0.5367) | 0.5052 (0.4603, 0.5501) | 0.4698 (0.4238, 0.5158) | 1.2425 (1.1546, 1.3304) | 0.6750 (0.5769, 0.7731) | 1.1626 (1.0292, 1.2960) | 0.8000 (0.4770, 1.1230) |
| **Alkaline Phosphatase, Serum: Median** | | **91.4354 (89.2082, 93.6626)** | 83.4112 (79.4949, 87.3275) | 79.8194 (76.7216, 82.9173) | 92.7353 (87.2375, 98.2331) | 102.2099 (95.6687, 108.7511) | 101.3983 (90.9622, 111.8343) | 88.6557 (83.2322, 94.0791) | 104.6991 (86.6527, 122.7455) | 113.6726 (99.2746, 128.0706) | 114.5143 (102.0344, 126.9941) | 109.5000 (87.3855, 131.6145) |
| **Alkaline Phosphatase, Serum: Range** | | **21.0644 (19.4902, 22.6386)** | 16.2682 (14.3662, 18.1703) | 12.1670 (10.9711, 13.3629) | 10.3839 (8.7640, 12.0038) | 18.6641 (15.8467, 21.4816) | 21.8571 (18.8613, 24.8530) | 13.3868 (11.2230, 15.5506) | 83.5487 (63.3473, 103.7500) | 33.0476 (21.5561, 44.5391) | 81.7571 (59.8159, 103.6984) | 42.2500 (20.3143, 64.1857) |
| **Alkaline Phosphatase, Serum: Slope** | | **-2.5743 (-4.6469, -0.5018)** | -1.5872 (-2.8933, -0.2812) | -2.2250 (-3.8897, -0.5603) | -5.3344 (-9.1246, -1.5441) | -8.3699 (-12.5191, -4.2208) | -9.4719 (-16.6415, -2.3023) | -1.8572 (-4.2399, 0.5255) | 4.8836 (-1.9433, 11.7106) | 19.6059 (-25.5775, 64.7894) | -0.8968 (-8.6582, 6.8646) | -2.1190 (-12.3397, 8.1016) |
| **Anion Gap, Serum: Median** | | **13.7523 (13.6030, 13.9017)** | 12.8339 (12.6048, 13.0631) | 13.8135 (13.5409, 14.0862) | 11.5880 (11.1762, 11.9998) | 14.0649 (13.5687, 14.5611) | 15.4418 (14.9198, 15.9638) | 15.6085 (15.2368, 15.9802) | 13.1150 (12.4055, 13.8246) | 18.5536 (17.4130, 19.6942) | 13.3857 (12.5595, 14.2119) | 15.7500 (13.2487, 18.2513) |
| **Anion Gap, Serum: Range** | | **4.4902 (4.3370, 4.6434)** | 3.4174 (3.2318, 3.6031) | 3.4438 (3.2368, 3.6507) | 2.5123 (2.2474, 2.7773) | 5.1298 (4.6663, 5.5933) | 4.8319 (4.4683, 5.1955) | 4.5566 (4.0529, 5.0603) | 10.9823 (9.6583, 12.3063) | 8.7619 (7.6158, 9.9081) | 9.6857 (8.4861, 10.8853) | 8.2500 (5.5987, 10.9013) |
| **Anion Gap, Serum: Slope** | | **-1.2272 (-1.4766, -0.9778)** | -0.4637 (-0.6892, -0.2382) | -0.7916 (-1.1536, -0.4297) | -1.5302 (-2.1107, -0.9497) | -1.7794 (-3.0263, -0.5324) | -0.7525 (-1.5240, 0.0189) | -2.1693 (-3.3092, -1.0294) | -0.1776 (-0.3601, 0.0049) | -7.6873 (-10.3302, -5.0444) | -0.0495 (-0.8803, 0.7814) | -0.3075 (-1.0747, 0.4598) |
| **Aspartate Aminotransferase (AST/SGOT): Median** | | **109.7207 (93.3999, 126.0414)** | 67.4654 (64.5390, 70.3917) | 71.6705 (61.0676, 82.2734) | 96.2127 (80.3950, 112.0305) | 175.3187 (127.7244, 222.9130) | 138.9113 (97.9203, 179.9022) | 70.6226 (64.5978, 76.6475) | 234.2345 (-48.8535, 517.3226) | 329.6726 (201.6266, 457.7187) | 70.8786 (43.6466, 98.1105) | 217.1250 (-51.0331, 485.2831) |

| Labs | Feature Selected | all | C 1 | C 2 | C 3 | C 4 | C 5 | C 6 | C 7 | C 8 | C 9 | C10 |
|---|---|---|---|---|---|---|---|---|---|---|---|---|
| **Aspartate Aminotransferase (AST/SGOT): Range** | | **139.1182 (107.0813, 171.1551)** | 30.6909 (25.8638, 35.5180) | 32.8619 (21.0357, 44.6882) | 27.3944 (17.6074, 37.1814) | 226.6641 (141.7767, 311.5516) | 118.1602 (47.3162, 189.0041) | 44.9858 (25.0947, 64.8770) | 703.9558 (305.5635, 1102.348) | 677.4048 (363.9447, 990.8648) | 698.3857 (83.1348, 1313.637) | 324.0000 (-5.4484, 653.4484) |
| **Aspartate Aminotransferase (AST/SGOT): Slope** | | **63.5960 (28.5177, 98.6743)** | -4.4419 (-7.2270, -1.6567) | 6.2861 (-12.9373, 25.5094) | 0.2470 (-17.8340, 18.3279) | 77.4688 (-130.609, 285.5468) | 85.6015 (20.1935, 151.0096) | 3.7134 (-11.2519, 18.6787) | 92.4179 (-33.5598, 218.3956) | 997.2512 (389.6242, 1604.878) | 66.8139 (16.8832, 116.7445) | 231.3509 (-152.347, 615.0486) |
| **Auto Basophil #: Median** | | **0.0163 (0.0155, 0.0171)** | 0.0111 (0.0103, 0.0118) | 0.0132 (0.0119, 0.0146) | 0.0157 (0.0144, 0.0170) | 0.0201 (0.0167, 0.0235) | 0.0155 (0.0134, 0.0175) | 0.0211 (0.0176, 0.0246) | 0.0169 (0.0140, 0.0197) | 0.0313 (0.0248, 0.0378) | 0.0336 (0.0276, 0.0395) | 0.0513 (0.0263, 0.0762) |
| **Auto Basophil #: Range** | | **0.0155 (0.0143, 0.0166)** | 0.0095 (0.0084, 0.0107) | 0.0075 (0.0065, 0.0084) | 0.0099 (0.0083, 0.0115) | 0.0163 (0.0135, 0.0191) | 0.0125 (0.0101, 0.0149) | 0.0124 (0.0103, 0.0145) | 0.0463 (0.0368, 0.0559) | 0.0213 (0.0148, 0.0278) | 0.0907 (0.0716, 0.1098) | 0.1237 (0.0437, 0.2038) |
| **Auto Basophil #: Slope** | | **0.0031 (0.0011, 0.0051)** | 0.0004 (-0.0011, 0.0020) | -0.0027 (-0.0049, -0.0005) | 0.0002 (-0.0049, 0.0052) | 0.0172 (0.0060, 0.0283) | 0.0051 (0.0015, 0.0087) | 0.0002 (-0.0036, 0.0040) | -0.0003 (-0.0016, 0.0010) | 0.0373 (-0.0021, 0.0768) | 0.0002 (-0.0092, 0.0095) | -0.0028 (-0.0176, 0.0120) |
| **Auto Basophil %: Median** | | **0.1651 (0.1592, 0.1711)** | 0.1315 (0.1235, 0.1395) | 0.1509 (0.1386, 0.1633) | 0.1733 (0.1587, 0.1879) | 0.1660 (0.1419, 0.1900) | 0.1581 (0.1407, 0.1754) | 0.1979 (0.1806, 0.2151) | 0.1781 (0.1412, 0.2150) | 0.1994 (0.1646, 0.2342) | 0.3300 (0.2850, 0.3750) | 0.3750 (0.2033, 0.5467) |
| **Auto Basophil %: Slope** | | **-0.0048 (-0.0197, 0.0100)** | -0.0114 (-0.0287, 0.0060) | -0.0443 (-0.0682, -0.0204) | -0.0320 (-0.0774, 0.0133) | 0.0635 (-0.0044, 0.1313) | 0.0266 (-0.0115, 0.0646) | -0.0210 (-0.0667, 0.0248) | -0.0118 (-0.0218, -0.0018) | 0.1595 (-0.0878, 0.4068) | -0.0247 (-0.0870, 0.0375) | -0.0997 (-0.2263, 0.0270) |
| **Auto Eosinophil #: Median** | | **0.0229 (0.0161, 0.0298)** | 0.0064 (0.0050, 0.0078) | 0.0090 (0.0067, 0.0113) | 0.0297 (0.0243, 0.0351) | 0.0141 (0.0105, 0.0178) | 0.0161 (0.0119, 0.0204) | 0.0125 (0.0095, 0.0154) | 0.0201 (0.0131, 0.0272) | 0.0114 (0.0064, 0.0165) | 0.1589 (0.1047, 0.2132) | 1.4594 (-0.3390, 3.2578) |
| **Auto Eosinophil #: Range** | | **0.0401 (0.0275, 0.0526)** | 0.0142 (0.0112, 0.0173) | 0.0094 (0.0073, 0.0116) | 0.0257 (0.0198, 0.0317) | 0.0185 (0.0125, 0.0245) | 0.0230 (0.0174, 0.0285) | 0.0238 (0.0179, 0.0296) | 0.0704 (0.0530, 0.0879) | 0.0093 (0.0044, 0.0142) | 0.3184 (0.2390, 0.3978) | 3.3225 (0.1647, 6.4803) |
| **Auto Eosinophil #: Slope** | | **-0.0011 (-0.0090, 0.0067)** | 0.0004 (-0.0032, 0.0041) | -0.0054 (-0.0102, -0.0007) | -0.0022 (-0.0203, 0.0158) | -0.0027 (-0.0248, 0.0194) | 0.0061 (-0.0004, 0.0125) | -0.0033 (-0.0110, 0.0045) | 0.0009 (-0.0008, 0.0026) | -0.0003 (-0.0372, 0.0366) | -0.0261 (-0.1890, 0.1368) | 0.2094 (-0.0846, 0.5033) |
| **Auto Lymphocyte #: Median** | | **1.0607 (0.9390, 1.1824)** | 1.0310 (0.7541, 1.3078) | 0.7647 (0.7274, 0.8019) | 1.0733 (0.8696, 1.2770) | 1.0483 (0.9551, 1.1416) | 0.9333 (0.7166, 1.1500) | 0.9467 (0.8747, 1.0187) | 2.7254 (0.7747, 4.6762) | 1.3066 (1.1418, 1.4714) | 1.0995 (0.9142, 1.2848) | 1.1938 (0.7401, 1.6474) |
| **Auto Lymphocyte #: Range** | | **0.4639 (0.3815, 0.5464)** | 0.3482 (0.3149, 0.3816) | 0.2199 (0.1935, 0.2464) | 0.2781 (0.2124, 0.3439) | 0.3763 (0.3123, 0.4403) | 0.4569 (0.2024, 0.7113) | 0.3375 (0.2823, 0.3927) | 2.5641 (1.0112, 4.1170) | 0.4858 (0.2965, 0.6752) | 1.1149 (0.9010, 1.3287) | 1.6763 (0.2476, 3.1049) |
| **Auto Lymphocyte #: Slope** | | **0.0172** | -0.0209 | -0.1016 (-0.1768, | -0.1020 | -0.0825 | 0.2249 | -0.0827 | 0.2210 | 1.0921 | -0.0348 | -0.1231 |

| Labs | Feature Selected | all | C 1 | C 2 | C 3 | C 4 | C 5 | C 6 | C 7 | C 8 | C 9 | C10 |
|---|---|---|---|---|---|---|---|---|---|---|---|---|
| | | **(-0.0505, 0.0849)** | (-0.1064, 0.0646) | -0.0263) | (-0.2718, 0.0677) | (-0.2700, 0.1050) | (-0.0465, 0.4963) | (-0.1908, 0.0254) | (-0.1327, 0.5746) | (-0.1982, 2.3824) | (-0.0886, 0.0190) | (-0.5663, 0.3201) |
| **Auto Lymphocyte %: Median** | | **10.7033 (10.4429, 10.9638)** | 11.9743 (11.4730, 12.4756) | 10.1138 (9.6491, 10.5784) | 11.1144 (10.3031, 11.9257) | 9.1245 (8.5071, 9.7419) | 9.5072 (8.6789, 10.3355) | 10.2302 (9.5845, 10.8760) | 12.8670 (10.5613, 15.1726) | 9.3732 (8.2620, 10.4845) | 11.8564 (10.4666, 13.2463) | 11.6875 (7.0687, 16.3063) |
| **Auto Lymphocyte %: Slope** | | **-1.0503 (-1.4599, -0.6407)** | -0.7494 (-1.6310, 0.1322) | -1.2185 (-1.9384, -0.4986) | -2.3932 (-3.7474, -1.0390) | -2.2381 (-3.5202, -0.9561) | 0.3947 (-0.8979, 1.6873) | -1.4369 (-2.4666, -0.4072) | -0.6103 (-1.0262, -0.1944) | 0.0972 (-4.8200, 5.0144) | 0.1686 (-0.7274, 1.0646) | 1.6907 (-0.9531, 4.3346) |
| **Auto Monocyte #: Median** | | **0.5082 (0.4930, 0.5234)** | 0.4161 (0.3977, 0.4344) | 0.5165 (0.4720, 0.5611) | 0.4900 (0.4592, 0.5208) | 0.5173 (0.4805, 0.5541) | 0.5060 (0.4587, 0.5533) | 0.5266 (0.4774, 0.5758) | 0.6251 (0.5532, 0.6970) | 0.6557 (0.5781, 0.7333) | 0.7832 (0.6275, 0.9389) | 0.8750 (0.4038, 1.3462) |
| **Auto Monocyte #: Range** | | **0.2302 (0.2165, 0.2440)** | 0.1767 (0.1594, 0.1939) | 0.1626 (0.1406, 0.1847) | 0.1600 (0.1313, 0.1886) | 0.2068 (0.1776, 0.2359) | 0.2024 (0.1737, 0.2311) | 0.2097 (0.1744, 0.2450) | 0.6591 (0.5459, 0.7723) | 0.2058 (0.1495, 0.2622) | 0.8973 (0.7171, 1.0775) | 1.3650 (0.3339, 2.3961) |
| **Auto Monocyte #: Slope** | | **-0.0588 (-0.0868, -0.0309)** | -0.0339 (-0.0570, -0.0108) | -0.0800 (-0.1320, -0.0281) | -0.0425 (-0.1194, 0.0344) | -0.0775 (-0.1774, 0.0224) | -0.0481 (-0.0996, 0.0033) | -0.0674 (-0.1180, -0.0169) | 0.0181 (-0.0359, 0.0722) | -0.3126 (-0.9381, 0.3129) | -0.0433 (-0.0922, 0.0056) | -0.1899 (-0.4041, 0.0244) |
| **Auto Monocyte %: Median** | | **5.4678 (5.3634, 5.5722)** | 5.3720 (5.1921, 5.5519) | 5.9894 (5.7344, 6.2444) | 5.5008 (5.1929, 5.8086) | 4.3027 (4.0665, 4.5390) | 5.4699 (5.1097, 5.8301) | 5.2507 (4.9761, 5.5254) | 6.0790 (5.5301, 6.6279) | 4.6482 (4.1899, 5.1066) | 7.3957 (6.6143, 8.1772) | 5.7937 (4.1913, 7.3962) |
| **Auto Monocyte %: Range** | | **2.3037 (2.1841, 2.4234)** | 2.1818 (1.9962, 2.3673) | 1.7427 (1.5566, 1.9288) | 1.6233 (1.3864, 1.8601) | 1.7949 (1.5397, 2.0502) | 2.3175 (1.9974, 2.6376) | 1.8776 (1.6218, 2.1335) | 6.5045 (5.1970, 7.8119) | 1.3655 (1.0209, 1.7100) | 7.4729 (6.2959, 8.6499) | 4.8250 (3.2594, 6.3906) |
| **Auto Neutrophil #: Median** | | **8.2255 (8.0599, 8.3912)** | 6.7810 (6.5442, 7.0178) | 7.3619 (7.0589, 7.6649) | 7.8762 (7.5167, 8.2357) | 10.9690 (10.3647, 11.5733) | 8.5091 (7.9782, 9.0400) | 8.4793 (8.0189, 8.9397) | 9.1230 (7.9782, 10.2679) | 12.8742 (11.8206, 13.9279) | 7.9037 (6.8769, 8.9306) | 13.5719 (4.5962, 22.5475) |
| **Auto Neutrophil #: Slope** | | **1.4168 (1.1528, 1.6807)** | 0.7884 (0.5401, 1.0366) | 0.7721 (0.3786, 1.1655) | 1.1933 (0.7070, 1.6796) | 3.7718 (2.6767, 4.8668) | 1.4097 (0.6640, 2.1554) | 0.6076 (0.0001, 1.2151) | 0.5386 (0.2874, 0.7897) | 9.3007 (3.9064, 14.6949) | 0.3898 (-0.4221, 1.2017) | -0.6031 (-3.2634, 2.0571) |
| **Auto Neutrophil %: Median** | | **80.9118 (80.5727, 81.2509)** | 80.4087 (79.8004, 81.0170) | 81.7385 (81.0755, 82.4016) | 80.2120 (79.1836, 81.2403) | 82.8508 (82.0563, 83.6453) | 81.9421 (80.8030, 83.0813) | 82.4290 (81.6445, 83.2136) | 77.3991 (74.7363, 80.0619) | 81.7482 (80.3888, 83.1077) | 73.4821 (70.7786, 76.1857) | 68.4250 (62.1516, 74.6984) |
| **Auto Neutrophil %: Range** | | **6.5079 (6.2175, 6.7982)** | 6.7376 (6.2192, 7.2559) | 4.4790 (4.0121, 4.9459) | 4.3925 (3.7659, 5.0192) | 4.5553 (3.8604, 5.2501) | 6.0450 (5.2705, 6.8195) | 5.8676 (5.0102, 6.7251) | 17.3848 (15.6407, 19.1289) | 4.5107 (3.3202, 5.7012) | 21.5114 (18.7528, 24.2701) | 27.5250 (21.1032, 33.9468) |
| **Auto Neutrophil %: Slope** | | **2.3470 (1.7478, 2.9461)** | 1.6965 (0.7382, 2.6548) | 3.1818 (2.2854, 4.0781) | 3.6309 (1.6867, 5.5751) | 4.9910 (1.4535, 8.5284) | 1.2068 (-0.4432, 2.8568) | 2.4590 (1.0026, 3.9155) | 0.6981 (0.1640, 1.2323) | 1.1277 (-3.4113, 5.6667) | 0.1616 (-1.8701, 2.1933) | -10.2253 (-29.0081, 8.5576) |

| Labs | Feature Selected | all | C 1 | C 2 | C 3 | C 4 | C 5 | C 6 | C 7 | C 8 | C 9 | C10 |
|---|---|---|---|---|---|---|---|---|---|---|---|---|
| **Basophil %: Range** | yes | **0.1540 (0.1443, 0.1637)** | 0.1238 (0.1119, 0.1357) | 0.0950 (0.0838, 0.1062) | 0.1056 (0.0906, 0.1206) | 0.1163 (0.0964, 0.1363) | 0.1323 (0.1066, 0.1581) | 0.1233 (0.1017, 0.1450) | 0.4027 (0.3330, 0.4723) | 0.1119 (0.0800, 0.1438) | 0.8986 (0.7522, 1.0450) | 0.8375 (0.2754, 1.3996) |
| **Bilirubin Total, Serum: Range** | | **0.2653 (0.2414, 0.2893)** | 0.1886 (0.1734, 0.2038) | 0.1864 (0.1675, 0.2054) | 0.1421 (0.1235, 0.1607) | 0.2748 (0.2349, 0.3147) | 0.2117 (0.1813, 0.2420) | 0.1981 (0.1714, 0.2249) | 0.8699 (0.6683, 1.0715) | 0.4440 (0.2801, 0.6080) | 1.1414 (0.5356, 1.7473) | 0.3000 (0.1187, 0.4813) |
| **Bilirubin Total: Median** | yes | **0.6487 (0.6161, 0.6814)** | 0.5631 (0.5400, 0.5863) | 0.5863 (0.5618, 0.6108) | 0.6762 (0.6338, 0.7186) | 0.7435 (0.6828, 0.8043) | 0.5706 (0.5274, 0.6137) | 0.6139 (0.5756, 0.6522) | 0.6469 (0.5529, 0.7409) | 0.7935 (0.6667, 0.9203) | 1.4843 (0.5136, 2.4550) | 0.5688 (0.3992, 0.7383) |
| **Bilirubin Total: Slope** | yes | **-0.0376 (-0.0533, -0.0220)** | -0.0124 (-0.0284, 0.0036) | -0.0393 (-0.0603, -0.0184) | -0.0699 (-0.1106, -0.0292) | -0.0168 (-0.0778, 0.0442) | -0.0726 (-0.1363, -0.0088) | -0.0669 (-0.1092, -0.0246) | 0.0598 (0.0066, 0.1131) | -0.1872 (-0.4072, 0.0329) | 0.0067 (-0.0674, 0.0809) | 0.0446 (-0.0255, 0.1146) |
| **Blood Urea Nitrogen, Serum: Range** | | **11.1261 (10.5649, 11.6873)** | 6.0195 (5.6253, 6.4137) | 8.5375 (7.7965, 9.2785) | 4.7037 (4.1655, 5.2419) | 8.4542 (7.5946, 9.3138) | 22.3147 (19.9044, 24.7249) | 8.4434 (7.3072, 9.5796) | 39.0265 (33.0438, 45.0093) | 14.0952 (11.6017, 16.5888) | 29.8571 (25.1992, 34.5151) | 30.8750 (11.1308, 50.6192) |
| **Blood Urea Nitrogen, Serum: Slope** | | **1.8699 (1.4694, 2.2704)** | 0.5505 (0.2227, 0.8783) | 1.4851 (0.8747, 2.0956) | 1.4320 (0.4964, 2.3675) | 5.1105 (3.7907, 6.4304) | -0.7025 (-3.3347, 1.9298) | 3.7564 (2.8010, 4.7117) | 0.6675 (-0.1355, 1.4706) | 9.5931 (6.8056, 12.3807) | 0.5989 (-1.8325, 3.0302) | 3.1468 (-1.1706, 7.4642) |
| **Blood Urea Nitrogen: Median** | yes | **30.8264 (30.0169, 31.6359)** | 15.3917 (14.9716, 15.8117) | 29.6531 (28.6705, 30.6358) | 25.1034 (24.0128, 26.1940) | 27.9275 (26.2760, 29.5790) | 77.0560 (73.8057, 80.3064) | 23.7524 (22.2025, 25.3022) | 35.6681 (32.1788, 39.1575) | 50.3274 (44.9919, 55.6629) | 35.0929 (30.5897, 39.5961) | 50.5000 (30.1359, 70.8641) |
| **C-Reactive Protein, Serum: Slope** | | **5.7647 (4.2167, 7.3128)** | 2.6489 (1.4488, 3.8489) | 1.8352 (-0.2099, 3.8804) | 1.6479 (0.3829, 2.9129) | 1.6928 (-1.3451, 4.7308) | 0.4514 (-1.4578, 2.3607) | 32.1646 (19.8908, 44.4384) | 2.2886 (0.6189, 3.9582) | 26.1175 (8.6456, 43.5893) | 5.1493 (-0.1314, 10.4301) | -11.5959 (-30.1946, 7.0028) |
| **C-Reactive Protein: Median** | yes | **41.1081 (38.7104, 43.5058)** | 21.1072 (19.2475, 22.9669) | 26.3775 (23.6195, 29.1355) | 20.1511 (18.1622, 22.1400) | 25.3034 (22.8179, 27.7890) | 23.0984 (19.1586, 27.0382) | 181.7371 (170.1435, 193.3307) | 13.2898 (10.7906, 15.7891) | 72.3767 (54.3717, 90.3818) | 48.5551 (35.2067, 61.9034) | 74.3725 (33.2223, 115.5227) |
| **C-Reactive Protein: Range** | yes | **9.0897 (7.9529, 10.2266)** | 5.3052 (4.4216, 6.1888) | 4.9300 (3.9309, 5.9291) | 1.6003 (1.2737, 1.9268) | 2.2260 (1.6794, 2.7726) | 3.6786 (2.5453, 4.8119) | 37.1552 (28.8747, 45.4356) | 11.3114 (8.3199, 14.3029) | 7.4944 (3.9804, 11.0084) | 43.5759 (26.1461, 61.0058) | 70.1250 (24.0720, 116.1780) |
| **Calcium Total: Median** | yes | **8.5179 (8.4971, 8.5387)** | 8.5155 (8.4878, 8.5431) | 8.8120 (8.7735, 8.8505) | 8.2989 (8.2554, 8.3424) | 8.0479 (7.9891, 8.1067) | 8.3129 (8.2285, 8.3973) | 9.0571 (8.9927, 9.1214) | 8.4522 (8.3571, 8.5473) | 8.5446 (8.4477, 8.6416) | 8.4300 (8.2947, 8.5653) | 8.3688 (7.7796, 8.9579) |
| **Calcium, Total Serum: Range** | | **0.7423 (0.7140, 0.7706)** | 0.5675 (0.5367, 0.5983) | 0.5412 (0.5050, 0.5775) | 0.3836 (0.3470, 0.4203) | 0.9958 (0.8718, 1.1198) | 0.9185 (0.8224, 1.0147) | 0.5882 (0.5181, 0.6583) | 1.8708 (1.7107, 2.0309) | 1.2012 (0.9495, 1.4529) | 1.6629 (1.4535, 1.8722) | 1.4875 (0.8418, 2.1332) |
| **Calcium, Total Serum: Slope** | | **-0.3288 (-0.3732,** | -0.1569 (-0.1915, | -0.2032 (-0.2592, | -0.1504 | -0.9405 (-1.1487, | -0.5096 (-0.6743, | -0.2328 (-0.3117, | -0.0662 (-0.1053, | -1.0515 (-1.3409, | -0.2354 (-0.3369, | -0.4903 |

| Labs | Feature Selected | all | C 1 | C 2 | C 3 | C 4 | C 5 | C 6 | C 7 | C 8 | C 9 | C10 |
|---|---|---|---|---|---|---|---|---|---|---|---|---|
| | | **-0.2845)** | -0.1223) | -0.1472) | (-0.3543, 0.0535) | -0.7323) | -0.3449) | -0.1538) | -0.0271) | -0.7621) | -0.1339) | (-1.1502, 0.1697) |
| **Carbon Dioxide, Serum: Median** | | **22.4864 (22.3493, 22.6235)** | 23.6350 (23.4210, 23.8490) | 22.9656 (22.6947, 23.2365) | 23.6914 (23.2978, 24.0849) | 21.1832 (20.7709, 21.5955) | 20.6185 (20.1132, 21.1239) | 21.6651 (21.2995, 22.0307) | 22.8894 (22.1602, 23.6185) | 18.2619 (17.5524, 18.9714) | 22.4786 (21.7070, 23.2501) | 21.4375 (20.0876, 22.7874) |
| **Carbon Dioxide, Serum: Range** | | **4.1069 (3.9746, 4.2392)** | 3.2966 (3.1212, 3.4721) | 3.2729 (3.0830, 3.4628) | 2.4475 (2.1988, 2.6962) | 4.3588 (3.9806, 4.7369) | 4.4224 (4.0636, 4.7813) | 3.6274 (3.2523, 4.0024) | 11.1062 (10.0242, 12.1882) | 6.2619 (5.5200, 7.0038) | 9.1857 (8.2537, 10.1177) | 7.7500 (5.1726, 10.3274) |
| **Carbon Dioxide, Serum: Slope** | | **0.2921 (0.0796, 0.5047)** | -0.0389 (-0.2537, 0.1759) | 0.1005 (-0.2594, 0.4604) | 0.6748 (0.0557, 1.2938) | -0.5109 (-1.4646, 0.4429) | -0.0265 (-0.5862, 0.5332) | 0.9873 (-0.0496, 2.0242) | 0.0702 (-0.1233, 0.2636) | 4.9941 (2.8735, 7.1147) | -0.5896 (-1.3467, 0.1675) | 1.0888 (-1.5912, 3.7688) |
| **Chloride, Serum: Median** | | **100.4280 (100.2070, 100.6491)** | 99.3863 (99.0544, 99.7183) | 100.1094 (99.7196, 100.4992) | 101.4784 (100.9625, 101.9943) | 100.5592 (99.7441, 101.3743) | 102.5862 (101.6055, 103.5670) | 98.1038 (97.4551, 98.7524) | 102.0973 (101.0406, 103.1541) | 100.9345 (99.1533, 102.7158) | 102.1857 (100.8203, 103.5511) | 100.7500 (98.4552, 103.0448) |
| **Chloride, Serum: Slope** | | **3.1794 (2.8899, 3.4688)** | 2.7044 (2.3592, 3.0496) | 2.5810 (2.2379, 2.9240) | 4.3480 (2.7526, 5.9435) | 6.1723 (5.1307, 7.2138) | 1.8544 (1.1767, 2.5322) | 2.4452 (1.6304, 3.2601) | 0.7054 (0.4341, 0.9768) | 7.6355 (5.7614, 9.5096) | 0.9115 (0.3176, 1.5054) | 0.2433 (-0.9317, 1.4183) |
| **Chloride: Range** | yes | **5.5630 (5.3862, 5.7398)** | 5.2593 (5.0040, 5.5147) | 4.3667 (4.1053, 4.6280) | 2.9414 (2.6378, 3.2449) | 5.8435 (5.3620, 6.3250) | 5.5991 (5.1240, 6.0743) | 4.6934 (4.2771, 5.1097) | 15.2212 (13.8725, 16.5700) | 7.7500 (6.5796, 8.9204) | 11.3286 (9.6923, 12.9648) | 7.8750 (4.7289, 11.0211) |
| **Creatinine, Serum: Median** | | **1.7512 (1.6834, 1.8189)** | 0.8270 (0.8112, 0.8428) | 1.4876 (1.3978, 1.5774) | 1.2986 (1.2383, 1.3589) | 1.6108 (1.5120, 1.7096) | 5.1186 (4.7202, 5.5170) | 1.2534 (1.1089, 1.3980) | 1.9651 (1.6121, 2.3181) | 2.7539 (2.3986, 3.1093) | 2.2419 (1.7613, 2.7224) | 3.2181 (1.8488, 4.5875) |
| **Creatinine, Serum: Range** | | **0.6261 (0.5868, 0.6654)** | 0.2476 (0.2298, 0.2654) | 0.3774 (0.3368, 0.4181) | 0.2722 (0.2429, 0.3016) | 0.6700 (0.5968, 0.7432) | 1.5853 (1.3791, 1.7915) | 0.3798 (0.3060, 0.4535) | 1.9304 (1.5035, 2.3572) | 1.1429 (0.9593, 1.3265) | 1.5199 (1.1639, 1.8758) | 2.1237 (0.6944, 3.5531) |
| **Creatinine, Serum: Slope** | | **0.0319 (-0.0077, 0.0716)** | -0.0795 (-0.0959, -0.0631) | -0.1153 (-0.1542, -0.0764) | -0.1013 (-0.1613, -0.0412) | 0.5663 (0.3174, 0.8153) | -0.1485 (-0.3477, 0.0507) | 0.0027 (-0.0667, 0.0721) | -0.0104 (-0.0670, 0.0462) | 0.9681 (0.7065, 1.2297) | 0.1197 (0.0120, 0.2275) | 0.0102 (-0.3287, 0.3491) |
| **D-Dimer Assay, Quantitative: Median** | | **2813.2858 (2517.552, 3109.019)** | 953.6986 (728.946, 1178.451) | 1252.4099 (953.278, 1551.542) | 3789.1646 (2902.241, 4676.088) | 6853.9158 (5212.204, 8495.627) | 2826.6966 (2063.352, 3590.042) | 2849.1524 (1872.151, 3826.154) | 2042.0435 (1245.955, 2838.132) | 10028.679 (7049.147, 13008.21) | 1805.9918 (968.862, 2643.122) | 2600.7857 (549.824, 4651.748) |
| **D-Dimer Assay, Quantitative: Range** | | **1167.2465 (976.810, 1357.683)** | 553.7397 (383.836, 723.644) | 588.4856 (333.654, 843.318) | 1164.0691 (534.851, 1793.287) | 1923.0000 (1118.942, 2727.058) | 891.7584 (413.112, 1370.405) | 1144.9893 (571.575, 1718.404) | 3491.1630 (1794.977, 5187.349) | 2723.5714 (1413.454, 4033.689) | 2475.8197 (1067.084, 3884.555) | 262.7143 (8.343, 517.085) |
| **D-Dimer Assay, Quantitative: Slope** | | **1866.3641 (607.060, 3125.669)** | 603.6803 (316.095, 891.266) | 257.2559 (-46.587, 561.099) | 4215.6207 (1146.913, 7284.328) | 3327.9307 (-909.034, 7564.895) | 349.3901 (-1035.78, 1734.561) | 405.0747 (-479.063, 1289.212) | 339.0205 (21.377, 656.664) | 19684.597 (-5460.25, 44829.45) | 579.8916 (96.031, 1063.752) | 1359.8000 (264.679, 2454.921) |

| **Labs** | **Feature Selected** | **all** | **C 1** | **C 2** | **C 3** | **C 4** | **C 5** | **C 6** | **C 7** | **C 8** | **C 9** | **C10** |
|---|---|---|---|---|---|---|---|---|---|---|---|---|
| **eGFR (non-AA): Median** | yes | **61.4461 (60.3631, 62.5292)** | 90.5036 (89.2016, 91.8055) | 54.2021 (52.6017, 55.8025) | 62.2654 (59.8897, 64.6412) | 56.1927 (53.2841, 59.1014) | 15.5841 (14.4698, 16.6983) | 71.7429 (68.6820, 74.8038) | 57.9823 (52.7896, 63.1750) | 35.0833 (30.8638, 39.3028) | 54.3929 (47.0678, 61.7179) | 34.6250 (17.1098, 52.1402) |
| **eGFR if African American: Median** | | **71.2251 (69.9695, 72.4807)** | 104.9112 (103.4022, 106.4201) | 62.8312 (60.9731, 64.6894) | 72.1806 (69.4275, 74.9336) | 65.1260 (61.7549, 68.4970) | 18.0733 (16.7843, 19.3623) | 83.1627 (79.6183, 86.7071) | 67.2080 (61.1881, 73.2279) | 40.5714 (35.6835, 45.4593) | 63.0786 (54.5768, 71.5803) | 40.1875 (19.6869, 60.6881) |
| **eGFR if African American: Range** | | **19.3365 (18.6740, 19.9990)** | 20.0853 (18.9406, 21.2299) | 15.4958 (14.4965, 16.4951) | 13.9290 (12.6947, 15.1634) | 26.0153 (23.6684, 28.3621) | 6.6034 (5.6993, 7.5076) | 17.9151 (16.0535, 19.7766) | 46.5575 (41.5543, 51.5607) | 23.2381 (19.2699, 27.2063) | 35.9000 (30.7681, 41.0319) | 33.8750 (15.3712, 52.3788) |
| **eGFR if African American: Slope** | | **2.0653 (1.1344, 2.9962)** | 7.5316 (6.2718, 8.7915) | 7.0811 (5.7584, 8.4039) | 10.4604 (7.8054, 13.1154) | -17.3299 (-22.1793, -12.4805) | -0.3993 (-1.3209, 0.5223) | 4.1708 (0.7802, 7.5614) | -0.2593 (-1.3486, 0.8301) | -24.6599 (-31.8950, -17.4249) | -0.1233 (-2.5736, 2.3271) | 0.4198 (-3.1517, 3.9914) |
| **eGFR if Non African American: Range** | | **16.6899 (16.1154, 17.2645)** | 17.3108 (16.3232, 18.2985) | 13.3729 (12.5142, 14.2316) | 12.0370 (10.9738, 13.1003) | 22.2824 (20.2546, 24.3103) | 5.7241 (4.9439, 6.5044) | 15.4387 (13.8381, 17.0393) | 40.2124 (35.8760, 44.5488) | 20.0714 (16.6448, 23.4980) | 31.7143 (26.9409, 36.4876) | 29.2500 (13.2851, 45.2149) |
| **eGFR if Non African American: Slope** | | **1.9518 (1.1854, 2.7181)** | 6.5348 (5.4479, 7.6216) | 6.1306 (4.9889, 7.2723) | 9.1114 (6.8171, 11.4056) | -13.6411 (-17.3119, -9.9703) | -0.3908 (-1.2041, 0.4225) | 3.5425 (0.6523, 6.4327) | -0.2298 (-1.1718, 0.7123) | -21.1977 (-27.4630, -14.9324) | 0.0202 (-2.1003, 2.1408) | 0.4094 (-2.6552, 3.4740) |
| **Eosinophil %: Median** | yes | **0.2004 (0.1789, 0.2219)** | 0.0724 (0.0580, 0.0867) | 0.1099 (0.0831, 0.1367) | 0.3189 (0.2634, 0.3744) | 0.1204 (0.0900, 0.1508) | 0.1692 (0.1235, 0.2149) | 0.1314 (0.0971, 0.1658) | 0.2344 (0.1524, 0.3163) | 0.0738 (0.0415, 0.1061) | 1.4414 (1.1386, 1.7443) | 5.0812 (2.6050, 7.5575) |
| **Eosinophil %: Range** | yes | **0.3720 (0.3257, 0.4182)** | 0.1708 (0.1369, 0.2047) | 0.1227 (0.0942, 0.1512) | 0.2863 (0.2216, 0.3511) | 0.1358 (0.0984, 0.1732) | 0.2917 (0.2133, 0.3701) | 0.2619 (0.1948, 0.3290) | 0.7679 (0.6020, 0.9338) | 0.0548 (0.0284, 0.0811) | 3.6086 (3.0093, 4.2078) | 15.0750 (10.8152, 19.3348) |
| **Eosinophil %: Slope** | yes | **-0.0288 (-0.0764, 0.0189)** | 0.0042 (-0.0320, 0.0405) | -0.0689 (-0.1260, -0.0117) | -0.0942 (-0.2891, 0.1007) | -0.0163 (-0.1550, 0.1223) | 0.0401 (-0.0251, 0.1054) | -0.0384 (-0.1406, 0.0639) | 0.0023 (-0.0214, 0.0260) | -0.0268 (-0.2284, 0.1749) | -0.1733 (-0.9245, 0.5780) | 0.6114 (-2.2721, 3.4949) |
| **Ferritin, Serum: Median** | | **1957.3146 (1791.531, 2123.099)** | 1279.3226 (1194.820, 1363.826) | 1493.2855 (1274.493, 1712.078) | 1725.8349 (1343.966, 2107.704) | 2592.1451 (2093.847, 3090.443) | 3611.234 (2508.008, 4714.461) | 1527.6186 (1349.957, 1705.281) | 2002.0727 (1088.400, 2915.746) | 4359.8848 (2503.298, 6216.472) | 1926.7187 (1252.635, 2600.802) | 6297.2571 (1277.783, 11316.731 ) |
| **Ferritin, Serum: Range** | | **536.2774 (411.2299, 661.3250)** | 308.7146 (169.7356, 447.6936) | 352.8752 (208.2102, 497.5401) | 117.3455 (69.3163, 165.3746) | 468.0366 (189.8075, 746.2656) | 418.6022 (275.7755, 561.4288) | 356.9197 (210.6828, 503.1567) | 2303.3778 (570.8109, 4035.945) | 1503.2101 (448.4801, 2557.940) | 2440.7557 (590.6927, 4290.819) | 1560.1429 (286.6507, 2833.635) |
| **Ferritin, Serum: Slope** | | **511.5350 (244.0569, 779.0130)** | 132.8555 (77.6040, 188.1069) | 267.3264 (65.8713, 468.7816) | 263.6387 (-97.4909, 624.7682) | 1223.3710 (429.2209, 2017.521) | 283.4224 (35.4279, 531.4168) | 323.6099 (66.4668, 580.7530) | 276.9436 (29.4070, 524.4802) | 5169.5042 (-586.199, 10925.20) | 130.1020 (-54.5555, 314.7595) | 675.9484 (-940.868, 2292.765) |

| Labs | Feature Selected | all | C 1 | C 2 | C 3 | C 4 | C 5 | C 6 | C 7 | C 8 | C 9 | C10 |
|---|---|---|---|---|---|---|---|---|---|---|---|---|
| **Glucose, Serum: Range** | | **82.7474 (79.1133, 86.3816)** | 50.8703 (47.2538, 54.4868) | 65.0979 (59.7904, 70.4054) | 41.8457 (36.5419, 47.1495) | 86.5229 (77.9905, 95.0553) | 107.6250 (96.8155, 118.4345) | 89.6509 (78.3359, 100.9660) | 204.9469 (170.7107, 239.1831) | 227.6548 (188.9809, 266.3286) | 156.2000 (133.0091, 179.3909) | 123.2500 (70.6443, 175.8557) |
| **Glucose: Median** | yes | **175.2694 (172.4411, 178.0976)** | 143.3037 (140.1386, 146.4688) | 172.1042 (167.0362, 177.1721) | 155.8210 (150.6166, 161.0253) | 197.6737 (190.2369, 205.1104) | 188.1250 (179.1402, 197.1098) | 211.4316 (200.5371, 222.3261) | 148.0973 (140.8539, 155.3408) | 345.8333 (315.8141, 375.8526) | 151.5143 (132.8425, 170.1860) | 138.3125 (111.9623, 164.6627) |
| **Glucose: Slope** | yes | **-5.3145 (-9.6587, -0.9704)** | 0.0888 (-3.0408, 3.2185) | 3.0962 (-4.2048, 10.3971) | 6.8679 (-2.2780, 16.0139) | 4.5892 (-10.5965, 19.7748) | -9.4622 (-23.7255, 4.8010) | -10.1312 (-25.1943, 4.9319) | 2.1638 (-2.3922, 6.7198) | -139.0172 (-209.520, -68.5145) | -17.3327 (-39.3400, 4.6746) | 7.8566 (-30.2787, 45.9920) |
| **Hematocrit: Median** | | **38.4309 (38.2232, 38.6387)** | 39.7932 (39.4756, 40.1109) | 38.6823 (38.2662, 39.0984) | 38.9451 (38.4121, 39.4780) | 39.1823 (38.5926, 39.7720) | 34.9623 (34.1735, 35.7511) | 40.0363 (39.4441, 40.6286) | 34.4102 (33.2766, 35.5437) | 40.3125 (38.9319, 41.6931) | 32.0393 (30.8608, 33.2177) | 33.2313 (30.1219, 36.3406) |
| **Hematocrit: Range** | | **4.1301 (3.9972, 4.2631)** | 3.5966 (3.4040, 3.7892) | 3.2296 (3.0344, 3.4248) | 2.4426 (2.2268, 2.6584) | 4.0218 (3.6300, 4.4135) | 4.7017 (4.2499, 5.1535) | 3.5071 (3.1528, 3.8613) | 11.4301 (10.6121, 12.2481) | 4.8750 (3.9768, 5.7732) | 9.6586 (8.8683, 10.4489) | 8.1625 (4.2112, 12.1138) |
| **Hematocrit: Slope** | | **-1.7387 (-1.9771, -1.5004)** | -1.2271 (-1.4405, -1.0138) | -1.3694 (-1.7188, -1.0200) | -1.6262 (-2.1791, -1.0733) | -3.3698 (-4.0476, -2.6921) | -1.9907 (-3.4177, -0.5637) | -0.9912 (-2.2915, 0.3090) | -0.5436 (-0.7198, -0.3674) | -6.4390 (-8.0543, -4.8237) | -1.0716 (-1.6946, -0.4486) | -0.0985 (-1.6030, 1.4060) |
| **Hemoglobin: Median** | | **12.5535 (12.4810, 12.6261)** | 13.1924 (13.0753, 13.3095) | 12.6342 (12.4878, 12.7805) | 12.7140 (12.5257, 12.9024) | 12.7861 (12.5805, 12.9916) | 11.1797 (10.9202, 11.4392) | 13.1108 (12.9115, 13.3102) | 11.1004 (10.7273, 11.4736) | 12.9071 (12.4671, 13.3472) | 10.2421 (9.8423, 10.6419) | 10.7437 (9.8279, 11.6596) |
| **Hemoglobin: Range** | | **1.3470 (1.3038, 1.3902)** | 1.2115 (1.1470, 1.2761) | 1.0215 (0.9596, 1.0833) | 0.7929 (0.7216, 0.8642) | 1.3916 (1.2633, 1.5199) | 1.5056 (1.3583, 1.6529) | 1.1165 (1.0011, 1.2319) | 3.6504 (3.3815, 3.9194) | 1.5405 (1.2578, 1.8232) | 3.1343 (2.8657, 3.4029) | 2.7000 (1.3967, 4.0033) |
| **Hemoglobin: Slope** | | **-0.6798 (-0.7583, -0.6012)** | -0.5066 (-0.5781, -0.4351) | -0.5289 (-0.6357, -0.4222) | -0.7112 (-0.8809, -0.5415) | -1.4540 (-1.6761, -1.2319) | -0.6423 (-1.1247, -0.1600) | -0.3743 (-0.8002, 0.0517) | -0.1809 (-0.2389, -0.1229) | -2.2153 (-2.7326, -1.6979) | -0.3391 (-0.5684, -0.1097) | 0.0939 (-0.5420, 0.7298) |
| **Immature Granulocyte %: Median** | yes | **1.1083 (1.0529, 1.1636)** | 0.7105 (0.6710, 0.7499) | 0.7498 (0.7120, 0.7875) | 1.2741 (1.1379, 1.4103) | 1.5415 (1.3585, 1.7244) | 1.0091 (0.8594, 1.1589) | 1.2960 (1.1240, 1.4679) | 0.9314 (0.8060, 1.0567) | 2.1094 (1.7098, 2.5089) | 3.0530 (2.0428, 4.0632) | 3.8250 (1.1648, 6.4852) |
| **Immature Granulocyte %: Range** | yes | **0.4471 (0.4007, 0.4934)** | 0.2922 (0.2540, 0.3305) | 0.2024 (0.1727, 0.2320) | 0.1712 (0.1179, 0.2244) | 0.2390 (0.1740, 0.3040) | 0.3438 (0.2558, 0.4317) | 0.5794 (0.4345, 0.7243) | 1.1049 (0.8900, 1.3198) | 0.5188 (0.2769, 0.7606) | 3.3015 (2.4238, 4.1792) | 4.1375 (0.7010, 7.5740) |
| **INR: Median** | | **1.3187 (1.2981, 1.3393)** | 1.2357 (1.1972, 1.2743) | 1.2815 (1.2374, 1.3257) | 1.4120 (1.3355, 1.4885) | 1.3450 (1.2960, 1.3939) | 1.3464 (1.2852, 1.4076) | 1.2510 (1.1973, 1.3047) | 1.4648 (1.3317, 1.5978) | 1.3323 (1.2658, 1.3988) | 1.3807 (1.2999, 1.4616) | 1.4892 (1.3086, 1.6697) |

| Labs | Feature Selected | all | C 1 | C 2 | C 3 | C 4 | C 5 | C 6 | C 7 | C 8 | C 9 | C10 |
|---|---|---|---|---|---|---|---|---|---|---|---|---|
| **INR: Range** | | **0.1668 (0.1316, 0.2021)** | 0.0657 (0.0043, 0.1272) | 0.1067 (0.0462, 0.1673) | 0.1286 (0.0209, 0.2363) | 0.1096 (0.0494, 0.1699) | 0.1854 (0.0996, 0.2712) | 0.0658 (0.0236, 0.1079) | 0.8751 (0.5121, 1.2381) | 0.1331 (0.0815, 0.1847) | 0.4613 (0.2193, 0.7034) | 0.3850 (0.0646, 0.7054) |
| **Lactate Dehydrogenase, Serum: Range** | | **88.9699 (66.8033, 111.1365)** | 53.0320 (43.3700, 62.6941) | 29.1750 (22.2458, 36.1042) | 34.7026 (23.1888, 46.2164) | 89.7053 (52.3518, 127.0587) | 43.0294 (29.0736, 56.9852) | 68.1688 (40.1996, 96.1381) | 267.4521 (151.1117, 383.7924) | 523.5522 (95.4377, 951.6667) | 297.7000 (3.4424, 591.9576) | 197.6667 (-64.4199, 459.7532) |
| **Lactate Dehydrogenase, Serum: Slope** | | **211.0817 (43.5086, 378.6548)** | 14.2756 (-32.5768, 61.1281) | 42.9508 (13.4588, 72.4429) | 77.4257 (15.6289, 139.2226) | 46.4839 (-218.653, 311.6209) | -8.8494 (-65.6022, 47.9033) | 27.1591 (-32.4008, 86.7190) | 417.4391 (-191.901, 1026.779) | 2370.3202 (-52.8405, 4793.481) | 23.7534 (-27.6375, 75.1443) | 98.5357 (1.2924, 195.7789) |
| **Lactate Dehydrogenase: Median** | yes | **608.0809 (591.1146, 625.0472)** | 521.9926 (506.9664, 537.0188) | 495.6696 (475.1030, 516.2362) | 618.3578 (586.3856, 650.3299) | 842.9421 (784.2454, 901.6388) | 577.6875 (534.3921, 620.9829) | 601.3377 (565.9183, 636.7570) | 488.9726 (434.5057, 543.4396) | 1169.1493 (923.9289, 1414.370) | 472.9500 (397.9229, 547.9771) | 1016.5833 (398.6444, 1634.522) |
| **Lymphocyte %: Range** | yes | **4.6062 (4.3914, 4.8210)** | 5.4052 (5.0055, 5.8049) | 3.2152 (2.8605, 3.5699) | 2.8870 (2.4330, 3.3409) | 3.1447 (2.7081, 3.5814) | 3.7550 (3.2520, 4.2580) | 4.1657 (3.5351, 4.7963) | 12.9955 (11.2163, 14.7748) | 2.9417 (2.1938, 3.6895) | 12.9171 (11.0241, 14.8102) | 11.6250 (4.9452, 18.3048) |
| **Magnesium, Serum: Median** | | **2.2215 (2.2058, 2.2371)** | 2.1625 (2.1396, 2.1854) | 2.1088 (2.0813, 2.1362) | 2.2644 (2.2143, 2.3146) | 2.2801 (2.2294, 2.3308) | 2.3464 (2.2898, 2.4029) | 2.2281 (2.1841, 2.2721) | 2.1469 (2.1002, 2.1936) | 2.6212 (2.4914, 2.7509) | 2.1191 (2.0436, 2.1947) | 2.3187 (1.7922, 2.8453) |
| **Magnesium, Serum: Range** | | **0.2185 (0.2037, 0.2332)** | 0.1565 (0.1370, 0.1760) | 0.1432 (0.1215, 0.1649) | 0.0929 (0.0371, 0.1487) | 0.1805 (0.1420, 0.2190) | 0.1974 (0.1601, 0.2347) | 0.2057 (0.1657, 0.2457) | 0.7518 (0.6775, 0.8260) | 0.3385 (0.2414, 0.4355) | 0.6191 (0.5294, 0.7088) | 0.3000 (0.1610, 0.4390) |
| **Mean Cell Hemoglobin Conc: Median** | | **32.6454 (32.5947, 32.6962)** | 33.1617 (33.0700, 33.2535) | 32.6209 (32.5120, 32.7299) | 32.6144 (32.4729, 32.7558) | 32.6204 (32.4551, 32.7857) | 32.0050 (31.8430, 32.1669) | 32.8172 (32.6648, 32.9696) | 32.2518 (32.0331, 32.4704) | 31.9940 (31.7443, 32.2438) | 31.9579 (31.6661, 32.2496) | 32.3625 (31.8362, 32.8888) |
| **Mean Cell Hemoglobin Conc: Range** | | **1.1014 (1.0679, 1.1349)** | 0.9822 (0.9329, 1.0315) | 0.9027 (0.8501, 0.9553) | 0.6904 (0.6311, 0.7498) | 1.1282 (1.0411, 1.2154) | 1.1586 (1.0605, 1.2568) | 0.8939 (0.8012, 0.9865) | 2.7752 (2.5312, 3.0192) | 1.3333 (1.1343, 1.5324) | 2.5900 (2.3195, 2.8605) | 1.9000 (0.9136, 2.8864) |
| **Mean Cell Hemoglobin Conc: Slope** | | **-0.2769 (-0.3350, -0.2188)** | -0.2657 (-0.3332, -0.1981) | -0.1930 (-0.3059, -0.0800) | -0.4571 (-0.6305, -0.2836) | -0.8269 (-1.0281, -0.6257) | 0.1092 (-0.1682, 0.3866) | -0.2116 (-0.4098, -0.0135) | -0.0011 (-0.0702, 0.0680) | -0.4590 (-1.0238, 0.1058) | 0.0075 (-0.1988, 0.2137) | 0.4981 (-0.2768, 1.2730) |
| **Mean Cell Hemoglobin: Median** | | **28.8966 (28.8110, 28.9822)** | 29.0569 (28.8843, 29.2296) | 28.9440 (28.7565, 29.1314) | 28.7904 (28.5322, 29.0486) | 28.9788 (28.7293, 29.2284) | 28.7625 (28.4748, 29.0502) | 28.4267 (28.1810, 28.6723) | 29.0867 (28.7066, 29.4668) | 28.9327 (28.5450, 29.3205) | 28.9443 (28.4079, 29.4807) | 29.2625 (28.2964, 30.2286) |
| **Mean Cell Hemoglobin: Range** | | **0.6438 (0.6201, 0.6675)** | 0.6100 (0.5746, 0.6453) | 0.5775 (0.5366, 0.6184) | 0.4022 (0.3620, 0.4423) | 0.4859 (0.4289, 0.5429) | 0.6517 (0.5888, 0.7146) | 0.5415 (0.4902, 0.5928) | 1.7841 (1.5648, 2.0033) | 0.5738 (0.4858, 0.6618) | 1.5343 (1.3041, 1.7645) | 1.2750 (0.5448, 2.0052) |

| Labs | Feature Selected | all | C 1 | C 2 | C 3 | C 4 | C 5 | C 6 | C 7 | C 8 | C 9 | C10 |
|---|---|---|---|---|---|---|---|---|---|---|---|---|
| **Mean Cell Hemoglobin: Slope** | | **-0.0443 (-0.0749, -0.0137)** | -0.0227 (-0.0672, 0.0218) | -0.1095 (-0.1736, -0.0453) | -0.1316 (-0.2525, -0.0107) | 0.0500 (-0.0590, 0.1590) | -0.0571 (-0.1437, 0.0294) | -0.1067 (-0.2372, 0.0238) | -0.0047 (-0.0500, 0.0407) | 0.1903 (-0.0055, 0.3861) | 0.0553 (-0.0181, 0.1286) | 0.2876 (0.0187, 0.5565) |
| **Mean Cell Volume: Median** | | **88.5363 (88.3022, 88.7704)** | 87.5968 (87.1634, 88.0302) | 88.7204 (88.2155, 89.2253) | 88.2699 (87.5611, 88.9787) | 88.8521 (88.1723, 89.5319) | 89.9188 (89.0589, 90.7786) | 86.6750 (86.0352, 87.3148) | 90.1465 (89.1517, 91.1412) | 90.5464 (89.3419, 91.7510) | 90.6286 (89.1366, 92.1206) | 91.0125 (86.9072, 95.1178) |
| **Mean Cell Volume: Range** | | **2.4429 (2.3514, 2.5344)** | 1.7929 (1.6875, 1.8983) | 1.7602 (1.6448, 1.8756) | 1.4590 (1.3090, 1.6089) | 2.8576 (2.5820, 3.1333) | 2.6621 (2.3883, 2.9358) | 1.8825 (1.6458, 2.1192) | 7.4637 (6.7902, 8.1372) | 3.4738 (2.9591, 3.9885) | 6.6629 (5.8164, 7.5093) | 5.1500 (2.1555, 8.1445) |
| **Mean Cell Volume: Slope** | | **0.6084 (0.4537, 0.7630)** | 0.6360 (0.4982, 0.7738) | 0.1677 (-0.0675, 0.4029) | 0.8658 (0.5166, 1.2149) | 2.4736 (1.9458, 3.0014) | -0.5635 (-1.5760, 0.4489) | 0.1961 (-0.1443, 0.5364) | 0.0004 (-0.1055, 0.1064) | 1.8436 (0.3334, 3.3538) | 0.2084 (-0.4426, 0.8595) | -0.7071 (-2.2587, 0.8445) |
| **Monocyte %: Slope** | yes | **-1.2176 (-1.4110, -1.0242)** | -0.7480 (-0.9833, -0.5128) | -1.4446 (-1.7969, -1.0923) | -1.4105 (-2.0725, -0.7485) | -2.2300 (-3.0058, -1.4542) | -0.9559 (-1.4173, -0.4946) | -1.3201 (-1.7928, -0.8475) | -0.2285 (-0.5262, 0.0692) | -3.5541 (-6.9352, -0.1729) | -0.2755 (-0.6190, 0.0680) | -1.6710 (-2.9983, -0.3438) |
| **Neutrophil #: Range** | yes | **2.7138 (2.5583, 2.8693)** | 2.3107 (2.0945, 2.5268) | 1.7398 (1.5130, 1.9665) | 1.2829 (1.0963, 1.4695) | 2.6615 (2.2546, 3.0684) | 2.4989 (2.1233, 2.8745) | 2.4215 (2.0613, 2.7816) | 8.1797 (6.9718, 9.3877) | 3.2157 (2.3156, 4.1158) | 10.2594 (8.2015, 12.3174) | 14.6000 (3.5084, 25.6916) |
| **Nucleated RBC: Median** | | **0.2478 (0.1783, 0.3174)** | 0.0361 (0.0184, 0.0537) | 0.0463 (0.0209, 0.0717) | 0.1635 (0.0899, 0.2371) | 0.3976 (0.2281, 0.5670) | 0.6031 (0.0630, 1.1432) | 0.3280 (0.1839, 0.4722) | 0.2870 (0.0613, 0.5127) | 0.8214 (0.4576, 1.1853) | 0.7672 (0.2910, 1.2435) | 2.5000 (0.2129, 4.7871) |
| **Nucleated RBC: Range** | | **0.7086 (0.2932, 1.1239)** | 0.0962 (0.0413, 0.1510) | 0.0556 (0.0326, 0.0785) | 0.1115 (0.0634, 0.1597) | 0.5122 (0.2497, 0.7747) | 1.6443 (-0.7552, 4.0438) | 0.1854 (0.0340, 0.3367) | 5.0556 (-0.0912, 10.2023) | 0.8214 (0.3218, 1.3211) | 1.7586 (1.0357, 2.4816) | 4.3333 (0.4953, 8.1713) |
| **Nucleated RBC: Slope** | | **0.1692 (-0.0062, 0.3446)** | -0.0260 (-0.0770, 0.0251) | -0.0234 (-0.0536, 0.0068) | -0.0153 (-0.1016, 0.0709) | 0.8665 (-0.4095, 2.1425) | 0.5179 (-0.3945, 1.4302) | -0.1327 (-0.3016, 0.0362) | 0.1217 (-0.1347, 0.3780) | 0.5051 (-0.0704, 1.0806) | 0.1797 (-0.3510, 0.7105) | 1.4014 (-0.4038, 3.2065) |
| **pCO2, Arterial: Median** | | **42.9202 (42.3648, 43.4756)** | 40.3588 (39.4726, 41.2450) | 40.1366 (39.0676, 41.2055) | 42.8696 (41.2721, 44.4671) | 48.8865 (47.3114, 50.4616) | 40.6192 (39.2658, 41.9725) | 41.3710 (39.5499, 43.1920) | 39.4856 (37.5836, 41.3875) | 51.6229 (48.3417, 54.9041) | 44.3871 (40.6835, 48.0907) | 42.6000 (33.1998, 52.0002) |
| **pCO2, Arterial: Range** | | **12.2632 (11.5692, 12.9571)** | 8.9302 (7.6688, 10.1917) | 6.0396 (4.7871, 7.2922) | 9.0978 (7.1274, 11.0683) | 18.6614 (16.8638, 20.4589) | 10.4477 (8.7963, 12.0990) | 11.0968 (8.6712, 13.5224) | 18.9423 (15.5558, 22.3289) | 20.3494 (17.2280, 23.4708) | 19.0484 (14.5894, 23.5073) | 17.6000 (8.7483, 26.4517) |
| **pCO2, Arterial: Slope** | | **-0.4789 (-3.0185, 2.0608)** | 8.3989 (2.4704, 14.3273) | -1.3030 (-8.9704, 6.3643) | -3.2630 (-12.7705, 6.2444) | 0.8518 (-4.5853, 6.2889) | -1.7411 (-9.4500, 5.9678) | -13.7537 (-26.9987, -0.5087) | 1.2517 (-0.6118, 3.1151) | -10.3153 (-20.3560, -0.2746) | 0.1898 (-10.0551, 10.4347) | -7.4459 (-15.3351, 0.4433) |

| Labs | Feature Selected | all | C 1 | C 2 | C 3 | C 4 | C 5 | C 6 | C 7 | C 8 | C 9 | C10 |
|---|---|---|---|---|---|---|---|---|---|---|---|---|
| **pCO2, Venous: Median** | yes | **44.8132 (44.2697, 45.3567)** | 42.6263 (41.7725, 43.4801) | 45.6172 (44.4372, 46.7972) | 43.6008 (41.6579, 45.5438) | 48.2148 (46.1746, 50.2551) | 45.4565 (43.3073, 47.6058) | 43.3989 (42.2858, 44.5119) | 46.8548 (43.9976, 49.7121) | 48.7627 (46.0405, 51.4849) | 43.8375 (40.2173, 47.4577) | 40.5000 (37.4438, 43.5562) |
| **pCO2, Venous: Range** | | **2.9076 (2.5482, 3.2670)** | 1.9439 (1.4253, 2.4624) | 1.3919 (0.9417, 1.8422) | 0.9664 (0.4270, 1.5058) | 3.9141 (2.4792, 5.3489) | 3.4522 (2.1722, 4.7321) | 2.7809 (1.9482, 3.6136) | 6.7742 (4.6810, 8.8674) | 8.9153 (5.3855, 12.4450) | 6.1000 (2.9943, 9.2057) | 7.0000 (1.7004, 12.2996) |
| **pCO2, Venous: Slope** | | **6.0258 (-1.7916, 13.8431)** | -4.0691 (-18.5098, 10.3715) | -1.6363 (-23.0702, 19.7977) | -27.3488 (-88.1647, 33.4672) | 45.4590 (13.9015, 77.0165) | -4.9622 (-42.4600, 32.5355) | 6.9963 (-1.0843, 15.0770) | -0.5461 (-11.0179, 9.9258) | 42.8984 (2.5237, 83.2730) | 8.9001 (-3.1139, 20.9141) | -0.6878 (-1.5953, 0.2196) |
| **pH, Arterial: Median** | yes | **7.3436 (7.3384, 7.3487)** | 7.3971 (7.3893, 7.4049) | 7.3923 (7.3833, 7.4012) | 7.3551 (7.3420, 7.3682) | 7.2709 (7.2586, 7.2831) | 7.3248 (7.3096, 7.3399) | 7.3617 (7.3451, 7.3784) | 7.3709 (7.3564, 7.3854) | 7.2024 (7.1747, 7.2301) | 7.3202 (7.2932, 7.3471) | 7.2600 (7.1705, 7.3495) |
| **pH, Arterial: Range** | | **0.1083 (0.0980, 0.1186)** | 0.0758 (0.0660, 0.0857) | 0.0526 (0.0431, 0.0621) | 0.0766 (0.0611, 0.0920) | 0.1536 (0.1395, 0.1676) | 0.0905 (0.0776, 0.1035) | 0.0857 (0.0686, 0.1027) | 0.1775 (0.1522, 0.2028) | 0.1415 (0.1193, 0.1636) | 0.2798 (0.0868, 0.4729) | 0.1820 (0.1406, 0.2234) |
| **pH, Arterial: Slope** | | **-0.0040 (-0.0260, 0.0179)** | -0.0530 (-0.0964, -0.0095) | -0.0029 (-0.0732, 0.0675) | -0.0177 (-0.1021, 0.0667) | 0.0008 (-0.0369, 0.0386) | 0.0186 (-0.0154, 0.0526) | 0.0831 (-0.0075, 0.1737) | -0.0109 (-0.0318, 0.0101) | 0.0995 (0.0277, 0.1714) | -0.1357 (-0.3763, 0.1050) | 0.1224 (-0.0382, 0.2831) |
| **pH, Venous: Median** | | **7.3508 (7.3462, 7.3554)** | 7.3972 (7.3914, 7.4031) | 7.3699 (7.3628, 7.3770) | 7.3661 (7.3536, 7.3787) | 7.2969 (7.2786, 7.3152) | 7.2962 (7.2773, 7.3150) | 7.3658 (7.3569, 7.3747) | 7.3196 (7.2981, 7.3411) | 7.2312 (7.1999, 7.2624) | 7.3373 (7.3130, 7.3615) | 7.3064 (7.2202, 7.3927) |
| **pH, Venous: Range** | | **0.0221 (0.0196, 0.0246)** | 0.0149 (0.0108, 0.0189) | 0.0101 (0.0069, 0.0133) | 0.0096 (0.0045, 0.0147) | 0.0323 (0.0217, 0.0429) | 0.0239 (0.0154, 0.0324) | 0.0205 (0.0152, 0.0258) | 0.0540 (0.0387, 0.0694) | 0.0610 (0.0390, 0.0830) | 0.0503 (0.0342, 0.0663) | 0.0529 (0.0179, 0.0878) |
| **pH, Venous: Slope** | yes | **-0.0125 (-0.0634, 0.0383)** | 0.0288 (-0.0623, 0.1200) | 0.0943 (0.0074, 0.1812) | 0.1934 (-0.3436, 0.7304) | -0.3653 (-0.5356, -0.1950) | 0.0411 (-0.1699, 0.2520) | 0.0246 (-0.0670, 0.1162) | 0.0803 (-0.0555, 0.2162) | -0.2889 (-0.4559, -0.1220) | -0.0829 (-0.1782, 0.0123) | -0.0057 (-0.0123, 0.0010) |
| **Phosphorus Level, Serum: Median** | | **3.9772 (3.9009, 4.0535)** | 3.1154 (3.0389, 3.1918) | 3.4255 (3.3135, 3.5375) | 3.6053 (3.4604, 3.7503) | 4.6788 (4.4461, 4.9116) | 5.6026 (5.3617, 5.8435) | 3.5079 (3.3298, 3.6860) | 3.8014 (3.5506, 4.0523) | 6.2000 (5.5069, 6.8931) | 4.0278 (3.6781, 4.3775) | 4.9125 (3.6794, 6.1456) |
| **Phosphorus Level: Range** | yes | **0.9164 (0.8474, 0.9855)** | 0.4194 (0.3563, 0.4825) | 0.4066 (0.3414, 0.4717) | 0.1647 (0.1131, 0.2163) | 0.8236 (0.6736, 0.9735) | 0.9394 (0.7589, 1.1199) | 0.9024 (0.7148, 1.0900) | 3.2712 (2.8138, 3.7285) | 2.0446 (1.5020, 2.5871) | 2.8889 (2.3423, 3.4355) | 2.0500 (1.0172, 3.0828) |
| **Platelet Count - Automated: Median** | | **221.4757 (218.3004, 224.6510)** | 217.2718 (211.3190, 223.2245) | 208.8625 (202.9914, 214.7336) | 226.6481 (217.3421, 235.9542) | 239.7328 (229.8845, 249.5812) | 200.9375 (190.8551, 211.0199) | 246.2759 (235.3785, 257.1734) | 205.9336 (192.2710, 219.5963) | 248.0952 (228.2798, 267.9107) | 234.6214 (208.5265, 260.7164) | 230.1250 (170.2277, 290.0223) |

| Labs | Feature Selected | all | C 1 | C 2 | C 3 | C 4 | C 5 | C 6 | C 7 | C 8 | C 9 | C10 |
|---|---|---|---|---|---|---|---|---|---|---|---|---|
| **Platelet Count - Automated: Range** | | **57.0728 (54.8121, 59.3336)** | 55.5027 (51.7677, 59.2377) | 42.0542 (38.7896, 45.3187) | 29.8765 (26.6324, 33.1207) | 52.0229 (46.8263, 57.2195) | 49.9483 (44.4107, 55.4859) | 55.8491 (48.2489, 63.4492) | 162.5487 (145.5019, 179.5954) | 63.5357 (53.3786, 73.6928) | 163.0857 (137.7285, 188.4430) | 89.2500 (54.8992, 123.6008) |
| **Platelet Count - Automated: Slope** | | **2.0464 (-0.5427, 4.6356)** | 16.9307 (14.2243, 19.6371) | 7.6853 (3.9093, 11.4614) | -2.6467 (-9.7748, 4.4815) | -9.7040 (-21.1805, 1.7725) | -14.0805 (-23.3964, -4.7646) | 9.4466 (-2.3184, 21.2116) | 4.0345 (1.2383, 6.8306) | -41.9105 (-69.3158, -14.5051) | -11.6665 (-21.0047, -2.3282) | -30.2518 (-86.7900, 26.2865) |
| **pO2, Arterial: Median** | | **109.0160 (106.6515, 111.3806)** | 101.0149 (96.4436, 105.5862) | 103.5639 (97.3333, 109.7945) | 102.4837 (95.1462, 109.8212) | 116.0219 (110.5693, 121.4745) | 113.9128 (106.4862, 121.3394) | 98.8495 (91.6267, 106.0722) | 121.0385 (111.5367, 130.5402) | 126.8651 (114.9929, 138.7373) | 115.5323 (105.0929, 125.9716) | 122.0000 (85.1648, 158.8352) |
| **pO2, Arterial: Range** | yes | **67.6062 (64.0156, 71.1968)** | 51.7053 (44.6031, 58.8075) | 35.9207 (29.4012, 42.4402) | 38.6304 (30.6311, 46.6297) | 102.4741 (92.9524, 111.9958) | 67.5523 (57.8150, 77.2896) | 44.2043 (34.7015, 53.7071) | 129.2404 (111.5312, 146.9496) | 97.9398 (81.8428, 114.0367) | 89.9032 (71.4594, 108.3471) | 157.6000 (93.4537, 221.7463) |
| **pO2, Arterial: Slope** | | **4.8191 (-12.4241, 22.0622)** | 37.5918 (8.3523, 66.8313) | 15.1740 (-31.2275, 61.5755) | 34.8126 (-17.1834, 86.8086) | 5.4602 (-50.2238, 61.1443) | 3.8330 (-39.8566, 47.5226) | -25.7835 (-48.9666, -2.6004) | -20.9505 (-45.7062, 3.8053) | -64.3706 (-144.480, 15.7383) | 0.9188 (-30.1945, 32.0321) | -113.1814 (-285.050, 58.6872) |
| **pO2, Venous: Median** | | **40.4512 (39.3222, 41.5802)** | 41.2430 (38.5496, 43.9364) | 35.3546 (33.7386, 36.9705) | 42.5932 (38.1875, 46.9989) | 41.4992 (38.9028, 44.0956) | 46.6783 (42.4099, 50.9467) | 37.1966 (34.8937, 39.4995) | 42.9194 (35.8541, 49.9846) | 48.7119 (41.0005, 56.4232) | 39.9625 (35.7379, 44.1871) | 43.3571 (36.9345, 49.7798) |
| **pO2, Venous: Range** | | **7.0837 (5.9849, 8.1826)** | 5.2378 (3.6064, 6.8691) | 3.6667 (2.1759, 5.1575) | 2.7797 (1.2692, 4.2901) | 5.4531 (3.1516, 7.7547) | 9.6000 (5.3363, 13.8637) | 9.6067 (6.3334, 12.8801) | 9.2903 (6.5553, 12.0254) | 17.9492 (4.7307, 31.1676) | 16.4250 (9.8919, 22.9581) | 48.1429 (4.1524, 92.1333) |
| **pO2, Venous: Slope** | | **56.2407 (37.6430, 74.8384)** | 94.2212 (47.0999, 141.3426) | 90.9070 (24.3918, 157.4223) | -49.5740 (-199.866, 100.7178) | 46.9183 (9.7950, 84.0417) | 43.0009 (-17.5541, 103.5559) | 57.2108 (27.2904, 87.1312) | 7.1160 (-6.8370, 21.0690) | 121.2634 (32.8474, 209.6794) | 14.1424 (-19.5167, 47.8016) | 4.1166 (1.4381, 6.7951) |
| **POCT Blood Glucose.: Median** | | **188.6887 (185.5229, 191.8544)** | 163.3088 (158.3277, 168.2900) | 193.0736 (186.1634, 199.9838) | 177.1145 (169.2269, 185.0021) | 197.4749 (189.0007, 205.9491) | 185.9461 (177.1488, 194.7434) | 228.0345 (216.2884, 239.7806) | 160.4902 (152.4080, 168.5724) | 278.0467 (255.5711, 300.5222) | 156.5726 (142.8023, 170.3428) | 148.6875 (118.4812, 178.8938) |
| **POCT Blood Glucose.: Range** | | **112.6139 (107.9310, 117.2969)** | 79.2255 (71.1404, 87.3106) | 106.5644 (97.1374, 115.9914) | 63.3464 (53.2536, 73.4391) | 96.2965 (84.6240, 107.9690) | 127.5784 (114.4846, 140.6723) | 118.2552 (104.4742, 132.0362) | 197.6176 (173.8120, 221.4232) | 213.1733 (183.8562, 242.4904) | 181.5806 (152.2593, 210.9020) | 99.6250 (37.8327, 161.4173) |
| **POCT Blood Glucose.: Slope** | | **-23.2502 (-30.9956, -15.5048)** | 10.2142 (0.6421, 19.7862) | -2.0422 (-11.9163, 7.8320) | -3.3504 (-22.7499, 16.0490) | -54.2100 (-96.1626, -12.2573) | -28.7527 (-48.2075, -9.2978) | -32.1646 (-51.9537, -12.3755) | 1.3818 (-2.9835, 5.7472) | -216.6847 (-275.110, -158.2594) | -5.4560 (-20.6214, 9.7094) | 2.6831 (-24.2776, 29.6438) |
| **Potassium, Serum: Median** | | **4.2541 (4.2335, 4.2746)** | 4.0195 (3.9905, 4.0486) | 4.2290 (4.1913, 4.2667) | 4.1531 (4.1035, 4.2026) | 4.3414 (4.2672, 4.4156) | 4.7086 (4.6269, 4.7903) | 4.2249 (4.1675, 4.2823) | 4.1774 (4.1043, 4.2506) | 4.8970 (4.7604, 5.0336) | 4.3550 (4.2439, 4.4661) | 4.5250 (4.1682, 4.8818) |

| Labs | Feature Selected | all | C 1 | C 2 | C 3 | C 4 | C 5 | C 6 | C 7 | C 8 | C 9 | C10 |
|---|---|---|---|---|---|---|---|---|---|---|---|---|
| **Potassium, Serum: Slope** | | **0.0400 (-0.0118, 0.0919)** | 0.0303 (-0.0098, 0.0704) | -0.0073 (-0.0659, 0.0513) | 0.1707 (0.0497, 0.2916) | 0.3212 (-0.0320, 0.6743) | -0.3420 (-0.4748, -0.2093) | 0.0135 (-0.1359, 0.1628) | 0.0120 (-0.0168, 0.0408) | 0.1618 (-0.2325, 0.5561) | 0.0944 (-0.0342, 0.2230) | -0.0035 (-0.2125, 0.2056) |
| **Potassium: Range** | yes | **0.8029 (0.7766, 0.8291)** | 0.6089 (0.5737, 0.6441) | 0.5948 (0.5548, 0.6347) | 0.3941 (0.3573, 0.4309) | 1.0031 (0.9122, 1.0939) | 1.0487 (0.9575, 1.1399) | 0.6578 (0.5947, 0.7209) | 1.9982 (1.8559, 2.1406) | 1.4369 (1.2470, 1.6269) | 1.8043 (1.6387, 1.9698) | 1.3000 (0.8637, 1.7363) |
| **Procalcitonin, Serum: Median** | | **3.6346 (2.9643, 4.3049)** | 1.5711 (0.6699, 2.4724) | 1.7475 (1.1376, 2.3574) | 3.3537 (1.8518, 4.8557) | 8.9553 (5.0948, 12.8159) | 6.3125 (4.2883, 8.3367) | 2.2643 (1.2147, 3.3138) | 3.7486 (1.7974, 5.6998) | 9.8450 (0.9668, 18.7232) | 4.2943 (1.6458, 6.9428) | 4.6860 (-0.7872, 10.1592) |
| **Procalcitonin: Range** | yes | **1.5058 (1.0776, 1.9340)** | 0.4199 (0.1778, 0.6620) | 0.8884 (0.3387, 1.4382) | 0.4063 (0.1486, 0.6641) | 2.5068 (1.2348, 3.7789) | 1.9427 (0.8505, 3.0349) | 1.2066 (0.2689, 2.1442) | 2.6250 (0.9772, 4.2727) | 8.0284 (-0.9507, 17.0075) | 5.0708 (2.3685, 7.7732) | 6.5280 (-4.2055, 17.2615) |
| **Protein Total, Serum: Median** | | **6.9204 (6.8928, 6.9480)** | 6.9617 (6.9159, 7.0075) | 6.9936 (6.9421, 7.0452) | 7.0150 (6.9373, 7.0927) | 6.8397 (6.7533, 6.9261) | 6.7413 (6.6402, 6.8425) | 7.4021 (7.3301, 7.4742) | 6.2066 (6.0675, 6.3458) | 7.0298 (6.8898, 7.1697) | 6.1900 (6.0074, 6.3726) | 6.2500 (5.6415, 6.8585) |
| **Protein Total, Serum: Range** | | **0.8391 (0.8129, 0.8652)** | 0.7673 (0.7287, 0.8059) | 0.6238 (0.5837, 0.6640) | 0.4585 (0.4167, 0.5003) | 0.9618 (0.8862, 1.0374) | 0.9524 (0.8606, 1.0442) | 0.6611 (0.5902, 0.7321) | 1.9611 (1.8218, 2.1003) | 1.3929 (1.1738, 1.6119) | 1.7829 (1.5881, 1.9776) | 1.5875 (0.7102, 2.4648) |
| **Protein Total, Serum: Slope** | | **-0.6731 (-0.7194, -0.6268)** | -0.4674 (-0.5174, -0.4174) | -0.4608 (-0.5282, -0.3934) | -0.7301 (-0.8413, -0.6189) | -1.2493 (-1.4590, -1.0397) | -0.9427 (-1.1618, -0.7236) | -0.4950 (-0.6028, -0.3872) | -0.1601 (-0.2010, -0.1191) | -1.9743 (-2.3914, -1.5572) | -0.3896 (-0.5870, -0.1922) | -0.4558 (-0.9136, 0.0020) |
| **Prothrombin Time, Plasma: Median** | | **15.0722 (14.8563, 15.2881)** | 13.9898 (13.7254, 14.2541) | 14.6684 (14.2244, 15.1124) | 16.0929 (15.2528, 16.9330) | 15.3971 (14.8250, 15.9693) | 15.4620 (14.7705, 16.1535) | 14.3506 (13.7273, 14.9739) | 16.8544 (15.4119, 18.2969) | 15.3480 (14.5609, 16.1351) | 15.8419 (14.9140, 16.7699) | 17.0167 (14.8783, 19.1550) |
| **Prothrombin Time, Plasma: Range** | | **1.7175 (1.3703, 2.0648)** | 0.3700 (0.2322, 0.5079) | 0.8322 (0.3543, 1.3101) | 1.4093 (0.2625, 2.5562) | 1.3118 (0.5722, 2.0515) | 2.1906 (1.1543, 3.2270) | 0.7354 (0.2836, 1.1872) | 9.2137 (5.4887, 12.9387) | 1.5760 (0.9596, 2.1924) | 5.2824 (2.4574, 8.1073) | 4.3667 (0.5230, 8.2103) |
| **RBC Count: Median** | | **4.3660 (4.3402, 4.3919)** | 4.5594 (4.5192, 4.5996) | 4.3866 (4.3335, 4.4396) | 4.4411 (4.3759, 4.5064) | 4.4252 (4.3553, 4.4950) | 3.9129 (3.8177, 4.0080) | 4.6379 (4.5589, 4.7168) | 3.8410 (3.7064, 3.9757) | 4.4897 (4.3253, 4.6541) | 3.5576 (3.4096, 3.7055) | 3.6812 (3.3239, 4.0386) |
| **RBC Count: Range** | | **0.4728 (0.4576, 0.4880)** | 0.4226 (0.4004, 0.4448) | 0.3592 (0.3374, 0.3810) | 0.2786 (0.2537, 0.3035) | 0.4876 (0.4424, 0.5328) | 0.5194 (0.4679, 0.5708) | 0.4014 (0.3602, 0.4426) | 1.3033 (1.2017, 1.4049) | 0.5524 (0.4529, 0.6519) | 1.0756 (0.9878, 1.1634) | 0.9138 (0.4977, 1.3298) |
| **RBC Count: Slope** | | **-0.2299 (-0.2581, -0.2016)** | -0.1720 (-0.1981, -0.1459) | -0.1679 (-0.2049, -0.1310) | -0.2288 (-0.2891, -0.1686) | -0.5107 (-0.5882, -0.4332) | -0.2169 (-0.3905, -0.0433) | -0.1177 (-0.2745, 0.0391) | -0.0608 (-0.0808, -0.0408) | -0.8031 (-0.9894, -0.6168) | -0.1207 (-0.1988, -0.0426) | -0.0012 (-0.2121, 0.2097) |

| Labs | Feature Selected | all | C 1 | C 2 | C 3 | C 4 | C 5 | C 6 | C 7 | C 8 | C 9 | C10 |
|---|---|---|---|---|---|---|---|---|---|---|---|---|
| **Red Cell Distrib Width: Median** | | **14.2799 (14.2144, 14.3453)** | 13.5874 (13.4803, 13.6945) | 14.0202 (13.9043, 14.1361) | 14.4603 (14.2701, 14.6506) | 14.3643 (14.1721, 14.5565) | 15.2274 (15.0034, 15.4513) | 14.0130 (13.8409, 14.1851) | 15.3429 (14.9479, 15.7380) | 14.3405 (14.0564, 14.6246) | 16.0807 (15.5480, 16.6134) | 16.6188 (15.1711, 18.0664) |
| **Red Cell Distrib Width: Range** | | **0.4993 (0.4661, 0.5325)** | 0.3461 (0.3167, 0.3755) | 0.2687 (0.2483, 0.2892) | 0.2386 (0.2068, 0.2703) | 0.5183 (0.4254, 0.6113) | 0.4832 (0.3825, 0.5839) | 0.3349 (0.2669, 0.4029) | 2.2673 (1.8942, 2.6403) | 0.5286 (0.3802, 0.6770) | 1.9271 (1.5523, 2.3020) | 2.0875 (0.8935, 3.2815) |
| **Red Cell Distrib. Width: Slope** | yes | **0.1597 (0.1357, 0.1836)** | 0.1227 (0.1000, 0.1453) | 0.0444 (0.0207, 0.0681) | 0.1686 (0.1247, 0.2126) | 0.6225 (0.4960, 0.7491) | -0.0609 (-0.1959, 0.0740) | 0.0974 (0.0527, 0.1422) | 0.1347 (0.0953, 0.1741) | 0.5164 (0.3566, 0.6762) | 0.0717 (0.0000, 0.1434) | -0.2547 (-0.9437, 0.4343) |
| **Sodium, Serum: Median** | | **136.6761 (136.4892, 136.8630)** | 135.8996 (135.6230, 136.1763) | 136.8167 (136.4783, 137.1550) | 136.7022 (136.2667, 137.1376) | 135.8989 (135.2456, 136.5521) | 138.6875 (137.8609, 139.5141) | 135.4245 (134.8087, 136.0403) | 138.2168 (137.3747, 139.0589) | 137.9226 (136.3467, 139.4986) | 137.7500 (136.5797, 138.9203) | 137.8750 (135.4385, 140.3115) |
| **Sodium, Serum: Range** | | **4.9374 (4.7766, 5.0981)** | 4.7549 (4.5123, 4.9975) | 3.9708 (3.7313, 4.2104) | 2.6914 (2.3854, 2.9973) | 4.4656 (4.0531, 4.8782) | 5.0560 (4.6428, 5.4693) | 4.3396 (3.9336, 4.7456) | 13.4425 (12.2407, 14.6442) | 6.4167 (5.5238, 7.3095) | 10.9143 (9.2808, 12.5478) | 6.6250 (3.9162, 9.3338) |
| **Sodium, Serum: Slope** | | **2.2569 (1.9584, 2.5553)** | 2.2135 (1.9245, 2.5026) | 1.8822 (1.5658, 2.1985) | 3.4858 (1.5950, 5.3765) | 3.8791 (2.9923, 4.7660) | 1.1064 (0.4278, 1.7851) | 1.2422 (0.4999, 1.9846) | 0.6517 (0.4345, 0.8689) | 5.1729 (3.5137, 6.8321) | 0.2705 (-0.1099, 0.6509) | 1.0246 (-0.2040, 2.2533) |
| **WBC Count: Median** | | **10.1664 (9.9406, 10.3921)** | 8.4187 (8.0951, 8.7422) | 9.0474 (8.6176, 9.4772) | 9.6958 (9.2252, 10.1665) | 13.3594 (12.6750, 14.0439) | 10.4266 (9.8018, 11.0513) | 10.3217 (9.7757, 10.8678) | 12.1947 (9.8441, 14.5452) | 15.3217 (14.1108, 16.5325) | 10.8241 (9.2935, 12.3548) | 14.5787 (7.6599, 21.4976) |
| **WBC Count: Range** | | **4.4361 (4.2237, 4.6485)** | 3.4547 (3.1749, 3.7345) | 3.1810 (2.9078, 3.4541) | 2.2929 (2.0357, 2.5500) | 4.6845 (4.1700, 5.1991) | 4.3006 (3.8055, 4.7958) | 3.5829 (3.1668, 3.9990) | 13.7400 (11.9867, 15.4933) | 6.2021 (5.0881, 7.3162) | 13.7234 (11.1964, 16.2504) | 22.7800 (2.2583, 43.3017) |
| **WBC Count: Slope** | | **1.4268 (1.2124, 1.6412)** | 0.8252 (0.6103, 1.0400) | 0.8183 (0.4222, 1.2144) | 1.0810 (0.5451, 1.6170) | 3.7471 (2.7959, 4.6984) | 1.3676 (0.6134, 2.1218) | 0.7741 (0.2303, 1.3178) | 0.6588 (0.3430, 0.9746) | 6.8981 (4.1731, 9.6231) | 0.5062 (-0.5508, 1.5631) | 0.7825 (-2.0028, 3.5677) |

### Feature Importance Model Details

*Table S6:Feature Importance Model Details*

| Cluster | Cluster Size (full train set) | Regularization Strength | Median AUC-ROC (validation folds) | Number of Selected Features in Median Fold | AUC-ROC (held-out test set) | Number of Selected Features after train on all folds |
|---|---|---|---|---|---|---|
| **C1** | 563 | 0.145 | 0.7206 | 15 | 0.6656 | 21 |
| **C2** | 481 | 0.1 | 0.7297 | 10 | 0.7181 | 12 |
| **C3** | 325 | 0.2 | 0.6520 | 17 | 0.6196 | 12 |
| **C4** | 262 | 0.08 | 0.7321 | 6 | 0.7127 | 7 |
| **C5** | 232 | 0.12 | 0.6244 | 8 | 0.5483 | 13 |
| **C6** | 212 | 0.075 | 0.6464 | 1 | 0.5308 | 2 |
| **C7** | 113 | 0.125 | 0.6353 | 10 | 0.5714 | 10 |
| **C8** | 84 | 0.1 | 0.7619 | 4 | 0.6818 | 6 |
| **C9** | 70 | 0.055 | 0.5657 | 1 | 0.5536 | 1 |
| **C10** | 10 | NA | NA | NA | NA | NA |
| **All data** | 2350 | 0.0275 | 0.7357 | 16 | 0.6981 | 19 |

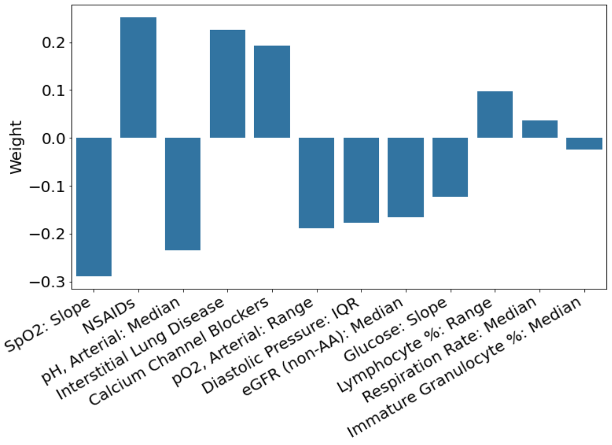


*Figure S4a: C3 Feature importance weights*

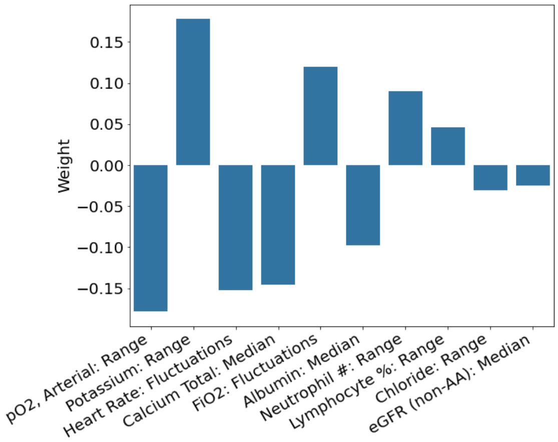


*Figure S4c: C7 Feature importance weights*

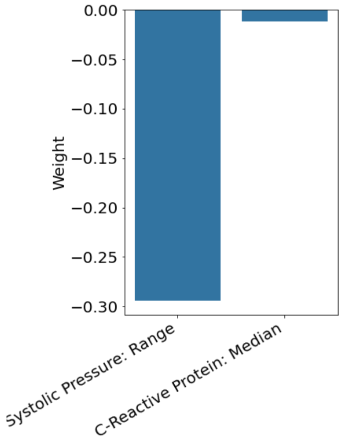


*Figure S4b: C6 Feature importance weights*

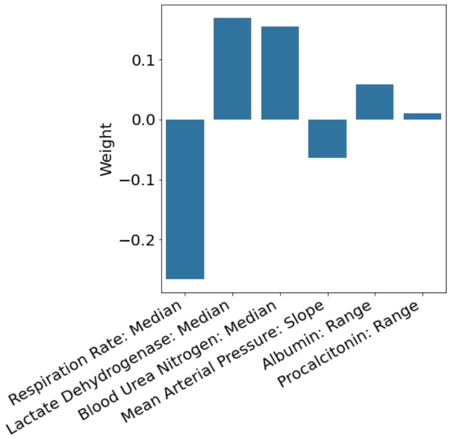


*Figure S4d: C8 Feature importance weights*

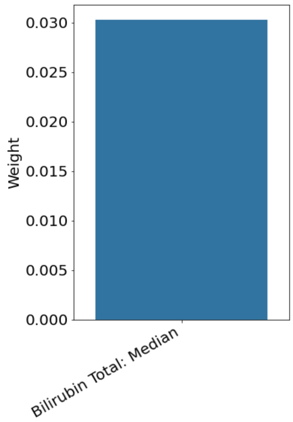


*Figure S4e: C9 Feature importance weights*

### Test data supplement

*Table S7: Cluster assignment distribution and mortality rate. Note that the overall mortality rate in the held-out test set is lower than in the train set. This is because after stratifying on demographics and mortality the initially censored patients were placed in the test set. This group had a lower mortality rate than the patients for whom complete data was initially available, causing the reduction in mortality in the held-out test set.*

| | **Train Set** | | | **Held-out Test Set** | | |
|---|---|---|---|---|---|---|
| **Cluster** | **Number in Cluster** | **Percent in Cluster** | **Mortality Rate** | **Number in Cluster** | **Percent in Cluster** | **Mortality Rate** |
| **C 1** | 563 | 24.0% | 62.0% | 121 | 23.0% | 61.2% |
| **C 2** | 481 | 20.5% | 82.5% | 99 | 18.8% | 73.7% |
| **C 3** | 325 | 13.8% | 77.2% | 79 | 15.0% | 72.2% |
| **C 4** | 262 | 11.1% | 63.0% | 51 | 9.7% | 54.9% |
| **C 5** | 232 | 9.9% | 89.7% | 70 | 13.3% | 87.1% |
| **C 6** | 212 | 9.0% | 81.6% | 43 | 8.2% | 69.8% |
| **C 7** | 113 | 4.8% | 77.0% | 24 | 4.6% | 70.8% |
| **C 8** | 84 | 3.6% | 82.1% | 17 | 3.2% | 64.7% |
| **C 9** | 70 | 3.0% | 70.0% | 20 | 3.8% | 70.0% |
| **C10** | 8 | 0.3% | 87.5% | 2 | 0.4% | 50.0% |
| **Overall** | 2350 | 100.0% | 74.7% | 526 | 100.0% | 69.6% |

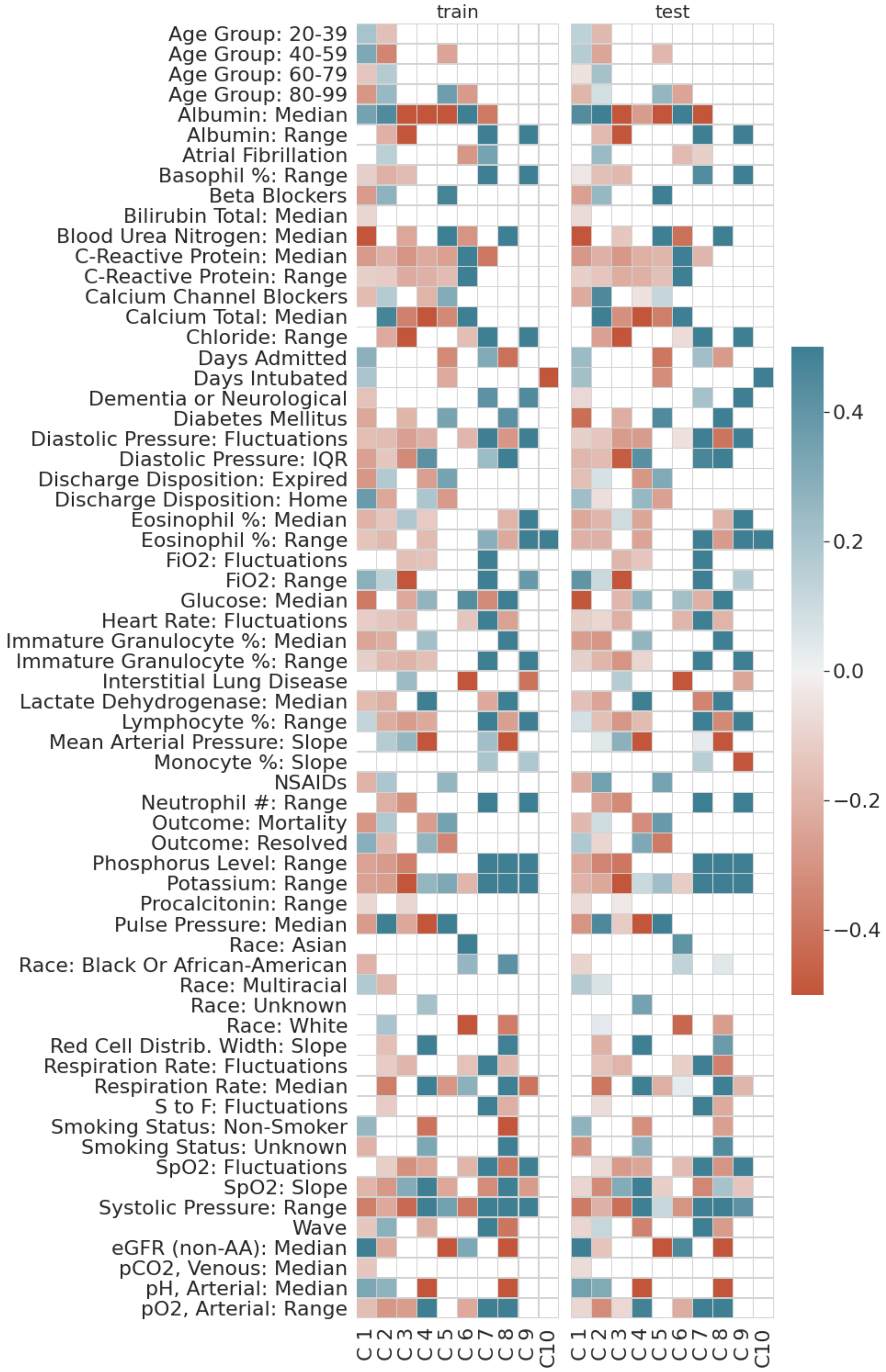


*Figure S5: Heatmap of enrichments. Only variable-phenocluster pairs that have a significant difference as compared to the rest of the training set are shown ($p < 0.05$). Colour intensity is defined by the number of training set standard deviations from the training set mean. Both the of the train set and held out test set are shown for comparison of the different levels. The same patterns emerge in both sets on all of these variables even though the phenoclustering was only performed on 5 principal components.*

#### Difference Between Higher Mortality and Lower Mortality Clusters

We examined features differentiating the lower and higher mortality phenoclusters from the rest of the patient dataset (**Figure 2**). Figure S provides data for all the clusters and includes a comparison of training and held-out test dataset. As expected, there are notable differences in average values for several variables (compared to the rest of the patients in the COVID19 ARDS training cohort dataset at the 95% significance level) between the lower and higher mortality groups (C1 or C4 vs. C2 or C5), and within the mortality groups between clusters (C1 vs. C4 and C2 vs. C5). These descriptive differences are explained with a reminder that they do not imply causal link to increased or decreased mortality.

**Lower mortality group (C1 and C4)**

Demographic features: Both phenoclusters in the lower mortality group (C1 and C4) were comprised of fewer people from wave 2, and fewer people who were prescribed calcium channel blockers as home medication. In cluster C1 there were more people who were younger, and who identified as multiracial, and fewer who identified as Black or African-American. Additionally, C1 had fewer people with diabetes or neurologic diseases including dementia, and fewer who were prescribed home beta blockers or NSAIDs (see **Supplemental Table 2** for all means with confidence intervals). In C4 there were more people with race unknown (meaning that this field was not filled in or other was selected).

Laboratory value features: We considered changes over time in laboratory values from hospital admission to 24 hours after T0, and compared these for each phenocluster to the rest of the training cohort. This included volatility (fluctuations), variation (range), level (median), and trend (slope). Laboratory features that defined both C1 and C4 included lower median eosinophils, lower range in immature granulocytes and lower CRP.

C1 was characterized by more people with high eGFR, high albumin, lymphocyte range and arterial PH (7.40 90%CI 7.40, 7.40); and lower low basophil range, bilirubin, BUN, phosphorous range, procalcitonin range, venous PCO2, glucose, LDH, Potassium range and less immature granulocytes. For C4 there were more people with high RDW slope (meaning more quickly increasing RDW over the course of the hospital stay) higher glucose, higher LDH and higher potassium range, and more immature granulocytes (median values). There were lower median values for calcium, albumin, and arterial PH (7.2709 90%CI 7.2586, 7.2831)), and lower lymphocyte range when compared to the rest of the cohort.

Vital signs and supplemental oxygen features: We also considered changes over time in vital signs from hospital admission to 24 hours after T0, and compared these for each phenocluster to the rest of the training cohort. Vital signs that defined both C1 and C4 included lower median pulse pressure and lower diastolic fluctuations. Those in C1 had a higher FiO2 range, and lower heart rate fluctuations, diastolic pressure range (Interquartile range, IQR), systolic blood pressure and arterial pO2 range. They also had lower SPO2 slope indicating less rapidly increasing SpO2 over the hospital admission. Those in C4 had higher respiration rates, diastolic blood pressure range (IQR), systolic blood pressure and arterial PO2 range. They also had a higher SpO2 slope indicating more rapidly increasing SpO2 over the hospital admission, lower FiO2, less SpO2 fluctuations, and lower MAP slope indicating more rapidly decreasing MAP over time.

**Higher mortality group (C2 and C5)**

Demographic features: Both phenoclusters in the higher mortality group (C2 and C5) were comprised of more people from the oldest age group 80-99, and more people who were prescribed NSAIDSs, Beta Blockers or calcium channel blockers as home medication. In cluster C2 there were more people who were identified as white, and more from wave 1, as well as more people who had atrial fibrillation as a comorbidity. There were less very young people in C2 (age 20-39), and less people who identified as multiracial. In C4 there were more people with diabetes as a comorbidity.

Laboratory value features:
Both C2 and C5 had lower levels of CRP and eGFR when compared to the rest of the training cohort. The comparatively lower level of CRP was also seen in the lower mortality group (C1 and C4). For the whole dataset, the overall mean for CRP was 41 units mg/L. C1, C2, C4 and C5 had average values in the 20mg/L range. Cluster 6 on the other hand had an average value of 181 mg/L. Of course, all these values are highly elevated compared to normal/healthy people. People in C2 had higher median arterial PH of 7.39 (90%CI 7.38,7.4) compared to 7.34 (90%CI 7.34,7.35) for the rest of the training cohort, higher albumin and higher total calcium. In addition, they had less eosinophils, immature granulocytes and LDH and lower RDW slope indicating less rapidly increasing RDW over time in the hospital. Interestingly, the ranges for several hematologic values were also lower than the rest of the cohort. People in C5 had on average lower median albumin and lower calcium median levels.

Vital signs and supplemental oxygen features:
Both C2 and C5 had higher median pulse pressure (a known risk factor for cardiovascular mortality especially in older people[2]). People in phenocluster C2 had higher MAP slope (indicating less rapid decrease in MAP), and greater range of FiO2 supplementation. They had lower diastolic blood pressure (IQR) with less fluctuations. Similarly, there were less fluctuations in heart rate, respiratory rate, SpO2, and S:F, and lower systolic pressure range. These variables suggest on average fixed lower levels of vital sign parameters. People in C5 had higher systolic pressure compared to the rest of the dataset.

#### Glossary

Phenoclusters – clusters of people with COVID-19 ARDS defined by their phenotypes up to 24 hours after the onset of hypoxemia

Lower mortality group - C1 and C4, these are the phenoclusters with significantly lower mortality than the rest of the cohort (p<0.05), that is they have enriched mortality and the it is lower than in the rest of the cohort

Higher mortality group – C2 and C5, these are the phenoclusters with significantly higher mortality than the rest of the cohort (p<0.05) ), that is they have enriched mortality and the it is higher than in the rest of the cohort

Variable – any variable available in the dataset. This includes labels, and all selected and not selected features.

Labels – features that are of interest regarding the patients, they may or may not be considered for feature selection and are of interest whether or not they were selected by feature selection. These are examined for enrichment across phenoclusters.

Possible Features – Features that could be selected by feature selection. This included only measurements available from admission to 24 hours after the onset of hypoxemia.

Selected Features – features that were selected by feature selection to be clustered on. These are examined for enrichment across phenoclusters and feature importance based on mortality within the phenoclusters.
Train set – all of the training data, five sixths of the data that was not censored

Train fold – four of 5 subsets of the training set that includes 80% of the train set

Validation fold – one of 5 subsets of the training set that includes 20% of the train set and is the compliment to a train fold

Held-out test set – the remaining one sixth of the uncensored data, plus the initially censored data.

Enrichment – a phenocluster is said to be enriched for a variable if the mean of that variable within that phenocluster is different at the 95% confidence level as compared to the mean of that variable in the rest of the dataset; in other words, the mean of that variable in the full dataset with the relevant phenocluster's patients excluded.
